# Defining and Reducing Variant Classification Disparities

**DOI:** 10.1101/2024.04.11.24305690

**Authors:** Moez Dawood, Shawn Fayer, Sriram Pendyala, Mason Post, Divya Kalra, Karynne Patterson, Eric Venner, Lara A. Muffley, Douglas M. Fowler, Alan F. Rubin, Jennifer E. Posey, Sharon E. Plon, James R. Lupski, Richard A. Gibbs, Lea M. Starita, Carla Daniela Robles-Espinoza, Willow Coyote-Maestas, Irene Gallego Romero

## Abstract

**Background:** Multiplexed Assays of Variant Effects (MAVEs) can test all possible single variants in a gene of interest. The resulting saturation-style data may help resolve variant classification disparities between populations, especially for variants of uncertain significance (VUS).

**Methods:** We analyzed clinical significance classifications in 213,663 individuals of European-like genetic ancestry versus 206,975 individuals of non-European-like genetic ancestry from *All of Us* and the Genome Aggregation Database. Then, we incorporated clinically calibrated MAVE data into the Clinical Genome Resource’s Variant Curation Expert Panel rules to automate VUS reclassification for *BRCA1, TP53, and PTEN*.

**Results:** Using two orthogonal statistical approaches, we show a higher prevalence (*p*≤5.95e-06) of VUS in individuals of non-European-like genetic ancestry across all medical specialties assessed in all three databases. Further, in the non-European-like genetic ancestry group, higher rates of Benign or Likely Benign and variants with no clinical designation (*p*≤2.5e-05) were found across many medical specialties, whereas Pathogenic or Likely Pathogenic assignments were higher in individuals of European-like genetic ancestry (*p*≤2.5e-05).

Using MAVE data, we reclassified VUS in individuals of non-European-like genetic ancestry at a significantly higher rate in comparison to reclassified VUS from European-like genetic ancestry (*p*=9.1e-03) effectively compensating for the VUS disparity. Further, essential code analysis showed equitable impact of MAVE evidence codes but inequitable impact of allele frequency (*p*=7.47e-06) and computational predictor (*p*=6.92e-05) evidence codes for individuals of non-European-like genetic ancestry.

**Conclusions:** Generation of saturation-style MAVE data should be a priority to reduce VUS disparities and produce equitable training data for future computational predictors.

## Introduction

Medicine faces challenges of unequal access and representation, particularly for individuals with non-European-like genetic ancestries, which results in a disproportionate number of inconclusive diagnostic outcomes for these populations^1–3^. This inequity is exacerbated in genomic medicine since the vast majority of research and clinical genomic sequencing to date has prioritized individuals of European-like genetic ancestries resulting in a comparative deficiency in knowledge about disease risk associated with genetic variants for individuals of non-European-like genetic ancestry^4–6^. This lack of diversity in control population data has repeatedly led to incorrect diagnoses^7^, missed diagnoses^8–11^ and inappropriate medical management^7^ for individuals of non-European-like genetic ancestry^7–9^. Compounding these challenges, genomic medicine can struggle to determine if identified genetic variants are harmful (Pathogenic or Likely Pathogenic; P/LP) or harmless (Benign or Likely Benign; B/LB), resulting in an exponentially increasing number of variants of uncertain significance (VUS) which are most commonly reported in individuals of non-European-like ancestry^8,10–14^. Thus, medicine urgently requires a systematic, population-scale understanding of variant classification inequities across genetic ancestries and a solution for large-scale reclassification of VUS, especially for individuals of non-European-like genetic ancestry.

Recent advances in functional genomics are enabling systematic, high-throughput experimental testing via Multiplexed Assays of Variant Effect (MAVEs), a family of methods able to characterize every possible SNV (single nucleotide variant) or indel (insertion or deletion) in a target gene and are being used for reclassification of VUS at scale^13,15–17^ (**Fig.1**). Thus, we hypothesized that the saturation nature of MAVEs would produce variant effects for VUS in individuals of non-European-like genetic ancestry leading to a higher rate of VUS reclassification compared to individuals of European-like genetic ancestry by compensating for the original VUS disparity. Further, we posited there would be an inequitable impact of different evidence towards VUS reclassification, but MAVE data would be used equitably. MAVEs mark a pivotal experimental advance in rectifying variant classification disparities and contributing to more equitable health outcomes for diverse populations worldwide.

**Figure 1:**
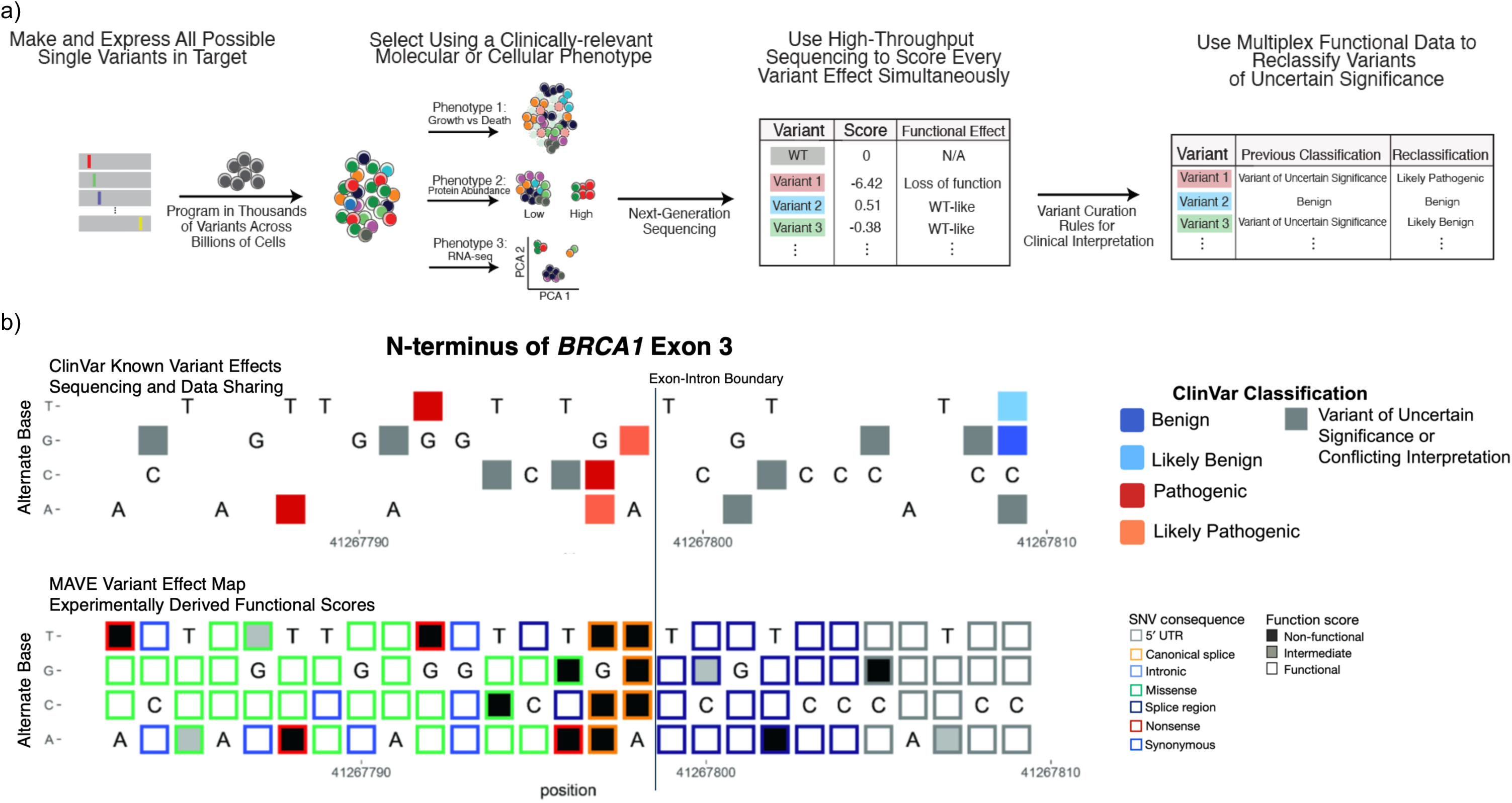
Multiplexed Assays of Variant Effects (MAVEs) Produce Saturation-level Variant Effect Maps Containing Functional Scores For Every Variant in a Target Locus. **a)** General scheme depicting the workflow of a MAVE starting with the design and construction of potentially every possible SNV or indel in a target locus. Next, the constructed variants are introduced into cells *in vitro*. MAVEs by their nature are able to test thousands of variant simultaneously across millions or potentially billions of cells ensuring each variant is programmed across thousands of cells for functional interrogation. After engineering the variants into the cells, a multiplexable phenotype such as cellular viability over time or fluorescence of an expressed protein is measured. Changes in the measured molecular phenotype for each variant are then read out via next generation sequencing. Functional scores are then calculated from the sequencing data for each variant. When used within the standard ACMG/AMP clinical interpretation framework, potential PS3/BS3 evidence codes of varying strengths dependent on clinical calibration of the functional scores can reclassify VUS. **b)** Both the top and bottom maps show the N-terminus of *BRCA1* Exon 3 for comparison purposes. The top map represents all the known ClinVar classifications for this particular locus as of November 2023 in the style of a MAVE variant effect map. The bottom map is an excerpt and adaptation of the *BRCA1* MAVE variant effect map from Findlay et al. (2018)^22^ where the experimental functional scores are depicted by shading and mutational consequences by the outline color of each SNV box. For both maps, reference nucleotides are indicated by the letters based on their position in the GRCh37 reference genome (position numbers of x-axis), the alternate nucleotides are indicated by the row labels (y-axis) and missing data is represented by no boxes. Notably, the MAVE variant effect map exhibits significantly higher information content with no missing SNV functional effects, while the map of clinical significance data contains much sparser information with VUS and missing data dominating the map. Of note, *BRCA1* is one of if not the most well studied gene in medical genetics. Thus, for most other genes the difference in information content would be even more pronounced as there would be even sparser clinical information, but the MAVE map would still be saturated. Further, because of the saturation nature of the MAVE map, there is no bias in variant selection to include in the assay – all variants in the target locus receive a functional score.

## Methods

### Data Curation and Allele Prevalence

Variants for each gene are available on the *All of Us* Public Data Browser (v7; https://databrowser.researchallofus.org/variants) or the Genome Aggregation Database (gnomAD) (https://gnomad.broadinstitute.org/). All databases assign a single genetic ancestry to each individual based on projection to principal components built using reference populations^9,18,19^. We have appended ‘-like’ to these descriptors to better reflect similarity to a particular set of reference populations^20^. We acknowledge their imperfect and incomplete nature as descriptors of continuous human diversity. The clinical variant classifications in this study found in *All of Us* or gnomAD were originally sourced from ClinVar. Variant calls, allele counts, population descriptors and variant classifications were used as prescribed by *All of Us* or gnomAD. Allele prevalence was calculated by summing allele counts for variants of each clinical classification for examined genetic ancestries and dividing this sum by the number of individuals in the genetic ancestry group(s).

### Two Orthogonal Statistical Methods

Two orthogonal statistical methods were used to assess variant classification disparities. First, at the gene-level using a matched pairs, Wilcoxon signed-rank test with Bonferroni correction resulting in a p-value, estimate of statistical power, and rank biserial coefficient with 95% confidence interval to quantify the magnitude of the differences using pre-established thresholds^21^. The matched pairs were the same gene’s allele prevalence between ancestry groups. Second, unique variants (not allele counts) for each clinical classification were counted across a gene list that were exclusive to each superpopulation group. If alleles for a unique variant were found in both superpopulation groups that unique variant was excluded from the counts. Then a chi-square test for independence with Bonferroni correction was conducted.

### Variant Reclassification and Essential Code Analysis

We developed an automated pipeline to reclassify VUS in *BRCA1*, *TP53*, and *PTEN* found in gnomAD and *All of Us*. These three genes were selected because all three have clinically calibrated MAVE data and Clinical Genome Resource’s (ClinGen) Variant Curation Expert Panel (VCEP) guidelines^13,22–29^. Our pipeline follows the gene-specific criteria of the corresponding VCEP as closely as possible except for the functional data evidence code (PS3/BS3) where MAVE data was used. To assess evidence code essentiality, we sequentially removed each code from the final set of codes for a reclassified VUS and observed if removal led to reversion of the reclassified variant back to VUS. Further details for any above methods can be found in the Supplementary Methods.

## Results

### Rationale for Selecting Databases

We analyzed genomes of 245,394 Americans in *All of Us* v7 and orthogonally validated our findings in two independent versions of the Genome Aggregation Database (123,709 exomes of gnomAD v2.1.1 and 51,535 genomes of gnomAD v3.1.2 (non v2)). We formed two superpopulation groupings: European-like and non-European-like genetic ancestry. Individual assignment was based on genetic ancestry labels reported by the respective database (Supplementary Methods)^9,18,19^. We chose these three population-scale databases, because the number of individuals sequenced in each was similar for both superpopulation groupings (**Fig.S1**) (non-European-like vs. European-like: 122,322 vs. 123,072 *All of Us* v7; 59,106 vs. 64,603 gnomAD v2.1.1; 25,547 vs. 25,988 gnomAD v3.1.2 (non v2)).

### Higher VUS Prevalence in Non-European-Like Genetic Ancestry

First, using the gene by gene statistical approach, we investigated allele prevalence differences of each clinical variant classification category between individuals of non-European-like versus European-like genetic ancestry at population scale. Individuals of non-European-like genetic ancestry exhibited significantly higher VUS prevalence across all medical specialties and gene groupings assessed in all three databases (*p*-values ranging 1.52e-211 to 1.4e-07; effect sizes ranging 0.35 to 0.76; **Fig.2, Fig.S2,** Tables S2-4). In contrast, P/LP classifications were significantly increased in individuals of European-like genetic ancestry (*p*-values ranging 2.3e-63 to 1.2e-04; effect sizes ranging −0.57 to −0.18; **Fig.S3**, Tables S2-4). Further, a significantly higher prevalence of B/LB and variants with no clinical designation (ND) was found in individuals of non-European-like genetic ancestry across several of the medical specialties (*p*-values ranging 2.9e-303 to 1.98e-05; effect sizes ranging 0.09 to 0.94; **Fig.S4-5**, Tables S2-4), while only isolated significant differences that did not validate across all three databases were seen for conflicting interpretation (CI) or noncoding variants (**Fig.S6-11**, Tables S2-6).

**Figure 2:**
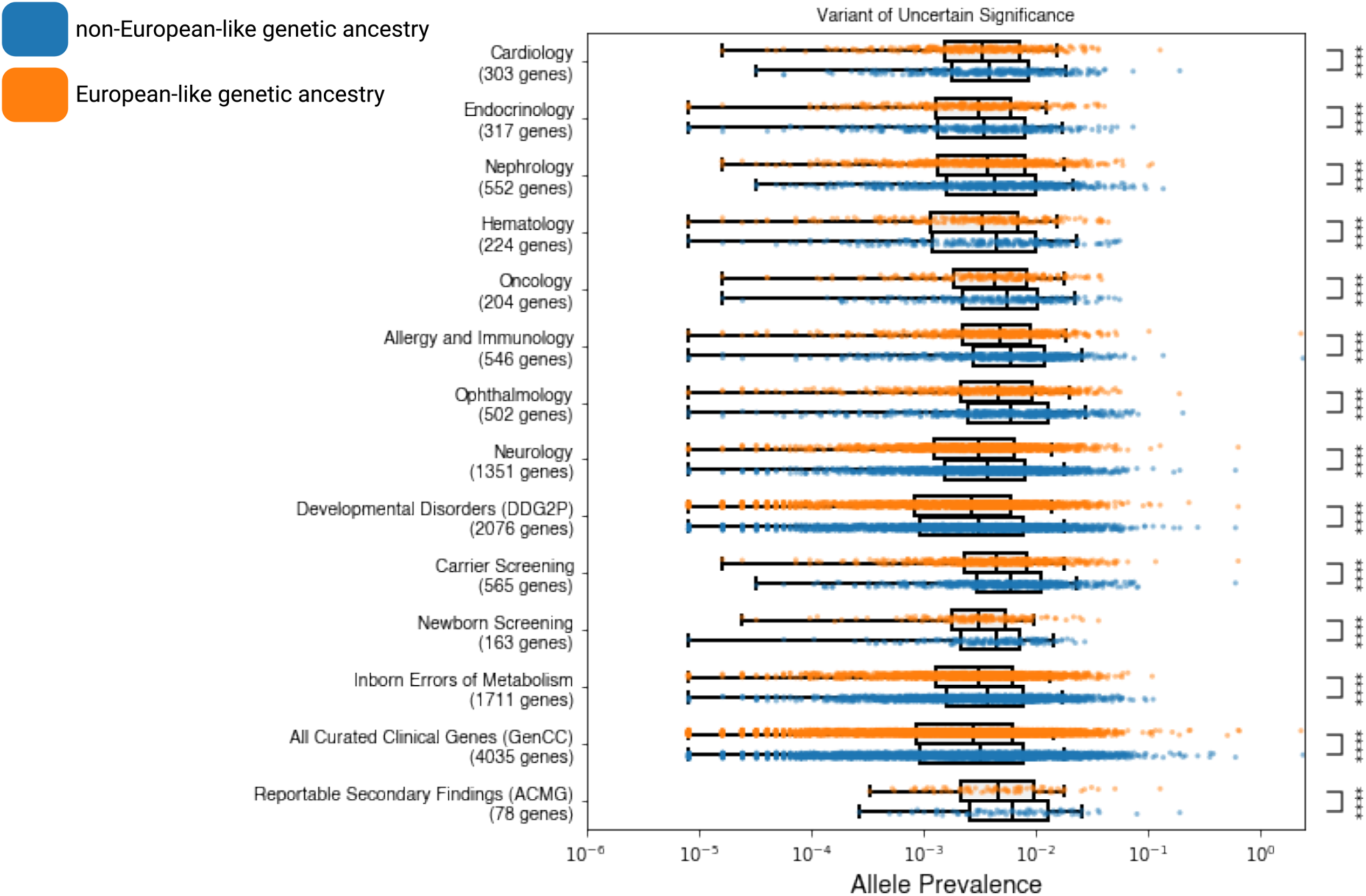
Higher VUS Prevalence Found in Individuals of Non-European-like Genetic Ancestry Across Medical Specialties. Box plots corresponding to VUS allele prevalence (x-axis) in each gene (dot) for individuals of non-European-like (blue) versus European-like (orange) genetic ancestry for the corresponding medical specialty (y-axis) as best visualized in *All of Us* v7 for all coding variants. Genes with zero alleles for allele prevalence for either individuals of European-like or non-European-like genetic ancestry are omitted from the above visualization to maintain a reasonable scale for data visualization. However, genes with zero alleles for only one category of either individuals of European-like or non-European-like genetic ancestry are included in the Bonferroni-corrected, signed rank, matched pairs Wilcoxon statistical test. The Bonferroni corrected p-values associated with these comparisons are annotated as follows with "ns" indicating not significant, * for 1.19e-04 < p ≤ 2.38e-04, ** for 5.95e-05 < p ≤ 1.19e-04, *** for 5.95e-06 < p ≤ 5.95e-05, and **** for p ≤ 5.95e-06. Across all medical specialties and categories shown, VUS are observed to be statistically significantly increased in individuals of non-European-like genetic ancestry compared to individuals of European-like genetic ancestry.

Next, to understand the magnitude and potential causes of VUS disparity, we ranked all curated clinical genes based on their difference in VUS allele prevalence and examined which genes were amenable to current MAVE techniques (**Fig.SF12-14** Tables S7-9). Over 83% of VUS across each medical specialty for all three databases were missense variants (**Fig.S15**). However, when missense VUS were excluded, the significant difference in VUS prevalence persisted (*p*-values ranging 2.78e-70 to 1.2e-05; effect sizes ranging 0.21 to 0.60; **Fig.3, Fig.S16**, Tables S10-12), emphasizing the VUS disparity is not driven solely by missense variants. In-frame indels, splice region, and synonymous variants also drove the VUS disparity (*p*-values ranging 1.63e-194 to 1.63e-04; effect sizes ranging 0.11 to 0.51; **Fig.S19**, Tables S13-15). All four of these variant types, missense, in-frame indels, splice region, and synonymous variants can be systematically catalogued via MAVEs.

**Figure 3:**
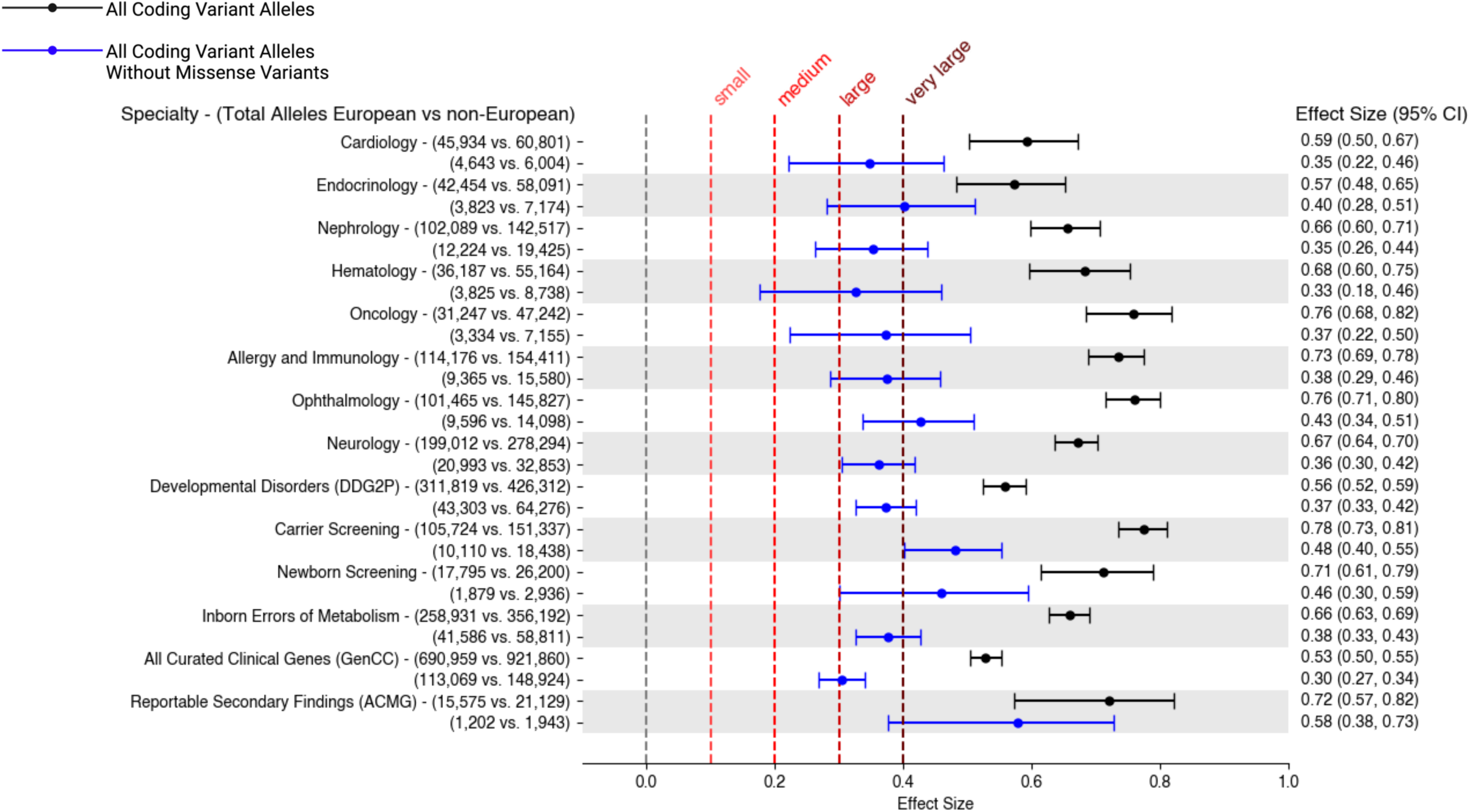
The Disparity in VUS Prevalence is Present Even in the Absence of Missense Variants. Effect size with 95% confidence interval (plotted and denoted on the right) shown for the differences between VUS prevalence in individuals of non-European-like versus European-like genetic ancestry as measured by the rank biserial coefficient from the signed rank, matched pairs, Wilcoxon test with a Bonferroni correction as best visualized in gnomAD v3.1.2 (non v2). The total number of alleles from individuals of non-European-like versus European-like genetic ancestry are indicated on the left. Effect sizes in black were calculated from all coding variants while effect sizes in blue were calculated from all coding variants excluding missense variants corresponding to the medical specialty (y-axis). Thresholds as determined by Funder and Ozer (2019)^21^ for quantifying the magnitude of the effect size difference are plotted as vertical dashed lines. Across medical specialties and categories, the disparity in VUS prevalence between individuals of non-European-like versus European-like genetic ancestry is not just statistically significant but very large. Further, the statistically significant disparity in VUS prevalence is still intact and medium to large even with the exclusion of missense VUS (∼85-90% of all VUS) across the medical specialties.

### Increased P/LP Classifications for European-like Genetic Ancestry

Using a second orthogonal statistical approach based on unique variants exclusive to only one superpopulation group, we show similar patterns for each clinical variant classification category. Across all medical specialties and all three databases, the non-European-like genetic ancestry group exhibited significantly higher counts of unique VUS, B/LB, CI, and ND variants (*p*-values ranging 7.97e-156 to 6.215e-18, **Fig.S25**), while pathogenic variants were the sole clinical classification where the European-like genetic ancestry group showed significantly higher counts (*p*=1.05e-05, **Fig.4a**). Further, the overlap of unique variants shared between superpopulation groups for VUS, P/LP, B/LB, and especially for CI variants is significantly greater relative to ND variants across every medical specialty in all three databases (*p*-values ranging 7.97e-156 to 6.215e-18, **Fig.4b**, **Fig.S26**). Thus, our current understanding of clinical variation especially pathogenic variation for individuals of non-European-like genetic ancestry is heavily shaped and limited by our existing knowledge of clinical variation in individuals of European-like genetic ancestry.

**Figure 4:**
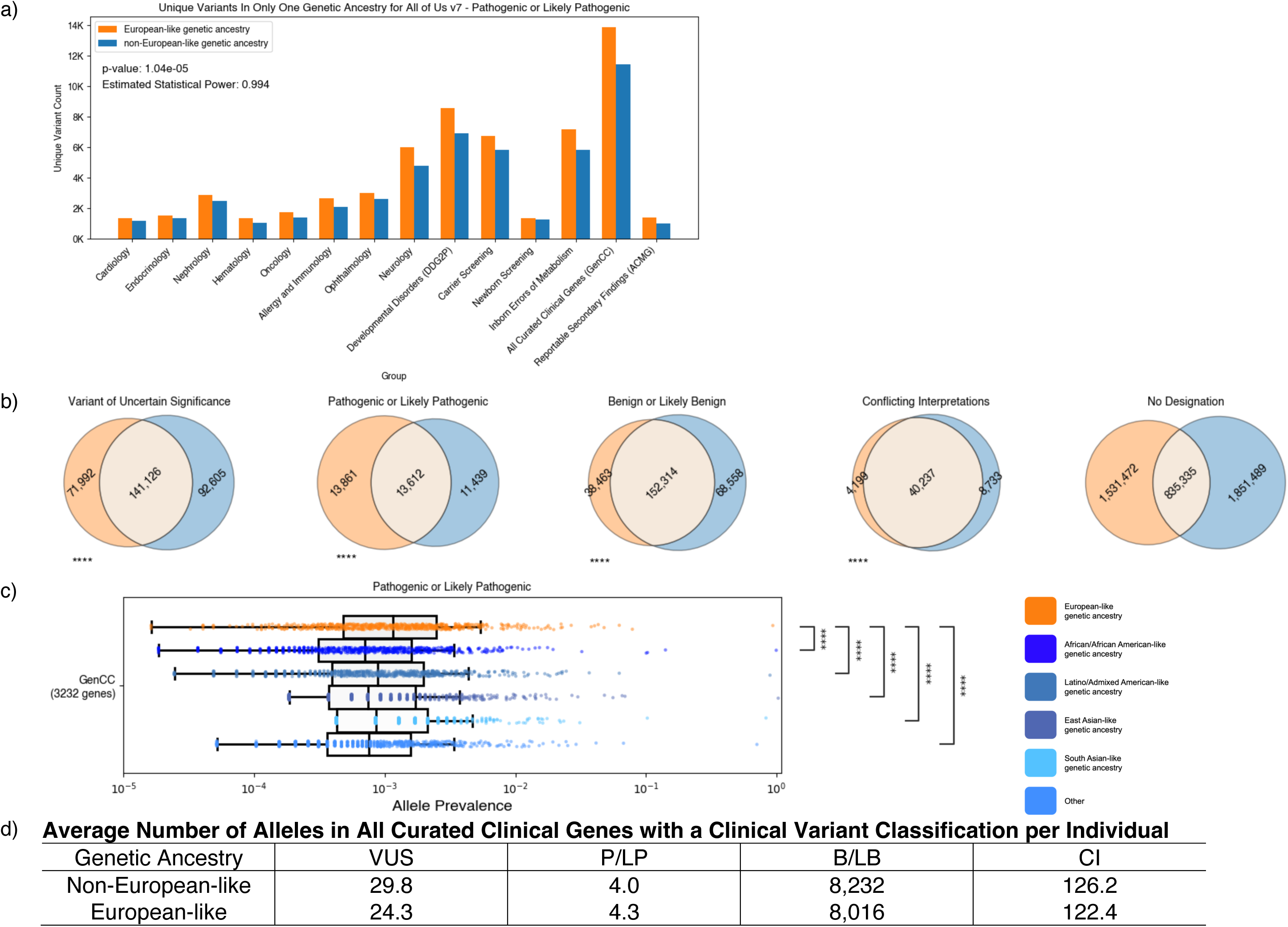
Lower Prevalence of Pathogenic or Likely Pathogenic Variants In Non-European-like Genetic Ancestry Group. a) Grouped bar graphs corresponding to unique coding variant counts (y-axis) for P/LP variants found either only in individuals of European-like (orange) genetic ancestry or only in individuals of non-European-like (blue) genetic ancestry across the medical specialties (x-axis) in *All of Us* v7. The Bonferroni corrected p-values from the chi-square test of independence associated with these comparisons are annotated along with the estimated statistical power. b) Venn Diagrams stratified by clinical variant classification (columns) for unique variants found either only in individuals of European-like (orange) genetic ancestry or only in individuals of non-European-like (blue) genetic ancestry across the medical specialties (y-axis) for all unique coding variant counts in *All of Us* v7 for the list of all curated clinical genes (GenCC). Across all medical specialties and categories shown, the overlap of unique variants for VUS, P/LP, B/LB, and CI variants is statistically significantly greater (chi-square test for independence) than the overlap between ND variants in all 3 databases across all medical specialties in both the presence and absence of missense variants. c) Box plots corresponding to Pathogenic or Likely Pathogenic allele prevalence (x-axis) in each gene (dot) for each non-European-like (shades of blue) genetic ancestry versus European-like (orange) genetic ancestry for all curated clinical genes (GenCC list; y-axis) across all three population databases (*All of Us* v7, gnomAD v2.1.1 and gnomAD v3.1.2 (non v2)) for all coding variants. Genes with zero alleles for allele prevalence for either individuals of European-like or non-European-like genetic ancestry are omitted from the above visualization to maintain a reasonable scale for data visualization. However, genes with zero alleles for only one category of either individuals of European-like or non-European-like genetic ancestry are included in the Bonferroni-corrected, signed rank, matched pairs Wilcoxon statistical test. The Bonferroni corrected p-values associated with these comparisons are annotated as follows with "ns" indicating not significant, * for 3.33e-04 < p ≤ 6.67e-04, ** for 1.67e-04 < p ≤ 3.33e-04, *** for 1.67e-05 < p ≤ 1.67e-04, and **** for p ≤ 1.67e-05. There is a statistically significantly increased prevalence of Pathogenic or Likely pathogenic variants found in the European-like genetic ancestry relative to each non-European-like group. d) Average number of alleles per individual based on weighted average of all three databases in all curated clinical genes (GenCC) for the differing clinical variant classifications.

Among all curated clinical genes (GenCC), all five genetic ancestries included in the non-European-like superpopulation group, African/African-American, Latino/Admixed American, South Asian, East Asian, and Other, demonstrated significantly decreased P/LP prevalence when compared to European-like genetic ancestry across all three databases (*p*≤1.67e-05; **Fig.4c**, **Fig.S27).** Overall, across all curated clinical genes and all three databases, there are an average of 29.8 VUS per individual of non-European-like genetic ancestry versus 24.3 VUS per individual of European-like genetic ancestry (**Fig.4d**).

### Integration of MAVE Data Reduces VUS Disparity

Next, we tested our hypothesis that the saturation nature of MAVE data would produce functional scores for VUS from individuals of non-European-like genetic ancestry and reduce VUS disparity. We built an automated VUS reclassification pipeline based on ClinGen VCEP rules for *BRCA1, TP53, and PTEN* with the amendment that we incorporated clinically calibrated MAVE data for the functional evidence codes. Given both the *All of Us* Public Data Browser and gnomAD are public genomic resources with deidentified variant data, we did not possess requisite individual-specific clinical histories to assess the clinically oriented evidence codes of the ClinGen VCEP criteria for gene-specific variant interpretation (**Fig.S28**). Thus, to validate the accuracy of our variant reclassifications, we benchmarked our pipeline against the Fayer et al. (2021)^13^ dataset where MAVE data was used for VUS reclassification. Our automated pipeline produced variant reclassifications that were 100% concordant for the 168 reclassified VUS in Fayer et al. (2021) (Table S18).

We found a significantly increased VUS prevalence (*p*=8.7e-06; one-tail z proportions test) for *BRCA1, TP53,* and *PTEN* across the three databases: 604 VUS across 206,975 non-European-like individuals assessed, compared to 480 VUS across 213,663 European-like individuals assessed (**Fig.5a**, Table S18). In individuals of European-like genetic ancestry, we reclassified 480 VUS as 315/480 (65.6%) Likely Benign, 4/480 (0.8%) as Benign, 16/480 (3.3%) as Likely Pathogenic, and 145/480 (30.2%) remained VUS (**Fig.5b**, Table S18, **Fig.SF29-30**). In individuals of non-European-like genetic ancestry, we reclassified the 604 VUS as 405/604 (67.1%) Likely Benign, 54/604 (8.9%) as Benign, 5/604 (0.8%) as Likely Pathogenic, and 140/604 (23.2%) remained VUS. MAVE evidence codes were used by most reclassified VUS alleles at 97.0% (775/799) compared to 75.8% (606/799) for computational predictors and 47.9% (383/799) for allele frequency (**Fig.5c**, Table S18). The statistically significant difference in reclassification rates (*p*=9.06e-03; one-tail z proportions test; **Fig.5b**, Table S18) between the two superpopulation groups resulted in nearly the same number of VUS remaining after reclassification in the non-European-like (140) and European-like (145) groups with no significant discernible disparity remaining (**Fig.5a**).

**Figure 5:**
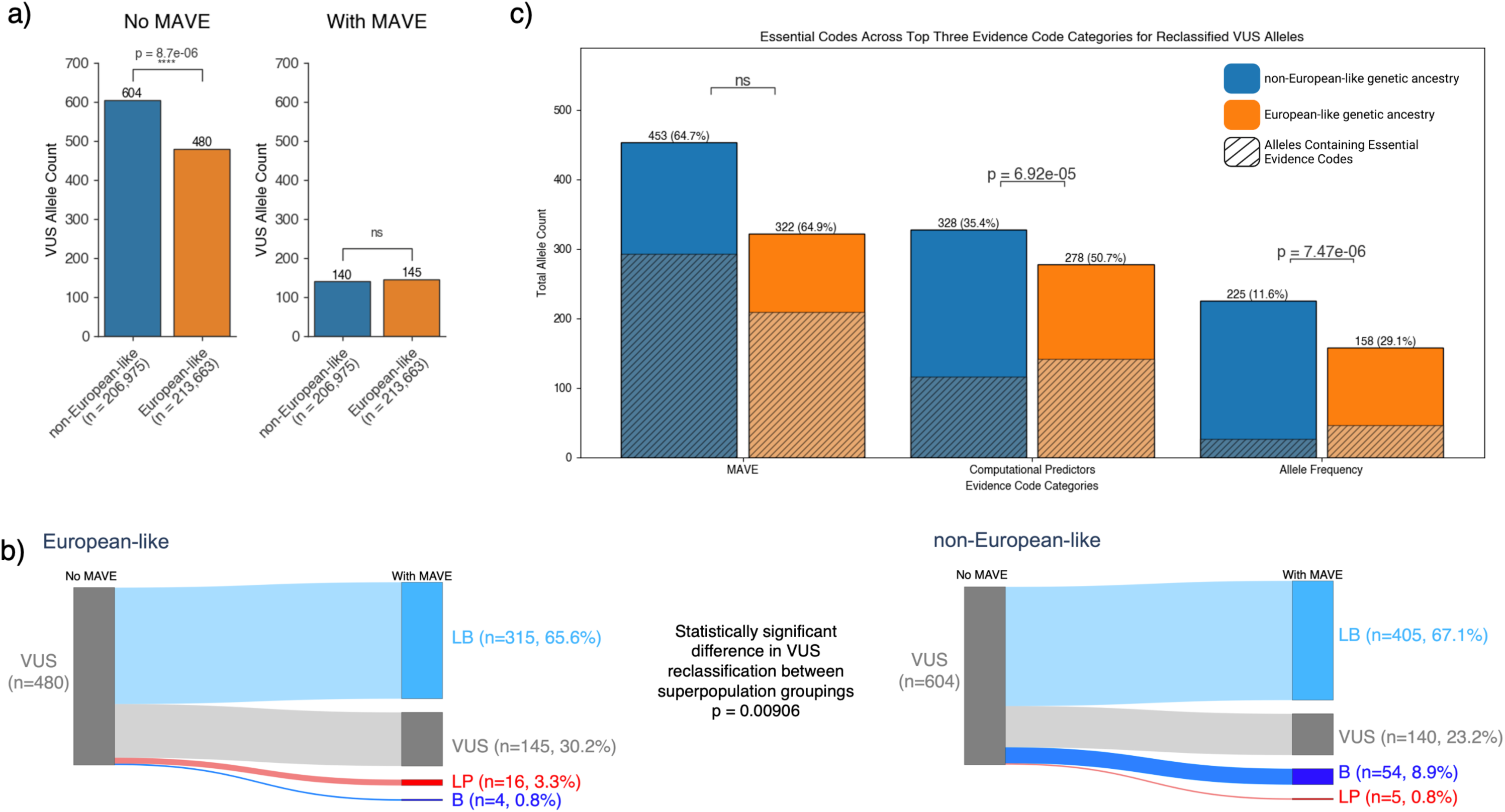
MAVE Data can Reclassify non-European-like VUS at a Statistically Significant Higher Rate Compared to European-like VUS. **a)** Sankey flow diagrams depicting VUS reclassification (read from left to right) for individuals of European-like (left) versus non-European-like (right) genetic ancestry before reclassification (No MAVE) and after reclassification (With MAVE). The examined VUS for *BRCA1*, *TP53* and *PTEN* are the total VUS alleles summed from all three databases *All of Us* v7, gnomAD v2.1.1, and gnomAD v3.1.2 (non v2) corresponding to the coding region saturated by the MAVE. The VUS were reclassified as either Likely Benign (LB; light blue), Benign (B; dark blue), Likely Pathogenic (LP; red), or remained as Variants of Uncertain Significance (VUS; gray). Reclassification was conducted using an automated pipeline based on the ClinGen Variant Curation Expert Panel gene specific variant interpretation guidelines for each gene with the amendment of using clinically-calibrated MAVE data for the functional evidence codes. **b)** The presence of VUS in individuals of non-European-like versus European-like genetic ancestry was statistically significantly higher in non-European-like superpopulation group. However, after using MAVE data for reclassification in the ClinGen VCEP frameworks, there was no statistically significant VUS disparity detected. **c)** Bar graphs for each evidence code category (x-axis) used in VUS reclassification across *BRCA1*, *TP53*, and *PTEN* for all three databases, *All of Us* v7, gnomAD v2.1.1, and gnomAD v3.1.2 (non v2). Blue bars represent alleles from individuals of non-European-like genetic ancestry, whereas orange bars represent alleles from individuals of European-like genetic ancestry. Shading represents essential codes, codes which if removed from the set of evidence codes used to reclassify the VUS would cause the variant to regress back to VUS. MAVE evidence codes were used the most based on total allele count for both individuals of non-European-like and European-like genetic ancestry. However, computational predictor and allele frequency codes were more essential for individuals of European-like genetic ancestry. PP3, PP3_Moderate, and BP4 correspond to the computational predictor codes. PS3, PS3_Moderate, BS3, BS3_Moderate, BS3_Supporting corresponded to the MAVE evidence codes. BA1, BS1, BS1_Supporting correspond to the allele frequency codes.

### Inequitable Impact of Computational Predictor and Allele Frequency Evidence Codes

We quantified "essential codes." For each variant, we deemed an evidence code as essential if removal would revert the reclassified variant back to VUS. We did not observe any significant difference in essentiality of MAVE codes between individuals of European-like (64.9%) versus non-European-like genetic ancestry (63.9%) (**Fig.5c**, **Fig.S31**, Table S18). Surprisingly, we did observe a significant difference in essentiality of computational predictor (37.3% non-European-like versus 49.8% European-like; *p*=1.65e-03, one-tail z proportions test) and allele frequency codes (7.0% non-European-like versus 21.6% European-like; *p*=1.13e-05, one-tail z proportions test, **Fig.5c**, Table S18). We validated this finding in the Fayer et al. (2021)^13^ dataset and observed no significant difference in essentiality of MAVE codes but a significant difference for computational predictor codes (77.9% non-European-like versus 84.1% European-like; *p*=5.73e-04, one-tail z proportions test; **Fig.S32**, Table S19).

This suggests the impact of computational predictor and allele frequency evidence codes towards VUS reclassification is not equitable for the two superpopulation groupings and, at least in part, describes the gap contributing to VUS disparity for which MAVE evidence compensates.

## Discussion

Our findings have important implications for ascertaining molecular diagnoses across medical specialties in patients of non-European-like genetic ancestry. Clinicians and genetic counselors should be aware when ordering next-generation sequencing (NGS) tests for non-European patients, there is a significantly higher pre-test probability of finding VUS or B/LB variants and significantly lower pre-test probability of finding P/LP variants relative to patients of European-like genetic ancestry. We show MAVE data reclassifies VUS at a significantly higher rate in individuals of non-European-like genetic ancestry compared to European-like compensating for the initial VUS disparity. Two prior studies reported VUS reclassification rates of 15.3%^30^ and 7.3%^12^ with clinical evidence codes being most important for VUS resolution^12^. Our study incorporated MAVE data and achieved a cumulative VUS reclassification rate of 73.7% without clinical evidence codes. Our VUS reclassification rate is similar to other single gene MAVE studies: 50% in *BRCA1*^13^, 69% in *TP53*, 75% in *MSH2*^31^ and 93% in *DDX3X*^32^. These findings underscore the necessity of proactive engagement in saturation-style MAVE data production for VUS reclassification at scale to advance our understanding of clinical variation in a more inclusive manner.

Importantly, the genetic ancestry groupings dictating our sample classifications are artificially bounded and not reflective of continuous human genetic variation^33^. We grouped individuals classified as non-European to improve statistical power due to limited sample sizes for each ancestry group. The non-European-like genetic ancestry group will contain a large number of admixed individuals, including many who are significantly admixed with individuals of European-like genetic ancestry. We hypothesize admixed individuals likely benefit from reduced VUS rates relative to more distantly related individuals, or those with reduced admixture proportions. Further, the population seen by clinical testing labs is significantly enriched in potential P/LP and VUS relative to population databases. Yet, we still identify consistent and significant trends across all three population databases independent of differences in genetic ancestry calculations, reference genome or NGS assay (**Fig.S1**).

Mechanistically, our findings suggest the increased VUS prevalence in individuals from non-European genetic ancestries is primarily due to the inability to interpret their genetic diversity. Non-Europeans had a significantly greater number of unique variants with no clinical designation and B/LB variants compared to Europeans, leading to a higher baseline prevalence of VUS in non-Europeans, which effectively remains uninterpreted due to the lack of sufficient evidence to classify these variants as pathogenic or benign. In contrast, Europeans had a higher P/LP prevalence. This discrepancy is attributable to historical disparities in access to genetic testing for individuals of non-European-like genetic backgrounds. As shown here, this has resulted in clinical variant databases enriched in clinically-relevant and pathogenic variation from individuals of European-like genetic ancestry giving a biased representation of global human genetic variation that hinders the interpretation of non-European-like genetic diversity.

This hinderance can be directly observed by the inequitable impact of the allele frequency and computational predictor evidence codes towards VUS reclassification. Allele frequency is directly impacted by the quantity and disproportionate levels of sequencing across populations. When computational predictors are trained and tested against excerpts of current sequencing and clinical variant databases^34,35^, there is a risk of overfitting on the distinctions between pathogenic and benign variations primarily within the European genetic ancestry group which may not always be translatable to other ancestry groups. Even though computational predictors produce saturation-style variant effects, we posit the lack of diverse training and testing data has perpetuated forward as AI bias preventing equitable impact for VUS reclassification and contributing to VUS disparity as seen in this study. Likely thousands of individuals of non-European-like genetic ancestry have had inequitable variant interpretations due to this bias in computational predictors. Thus, a systematic analysis should be undertaken to understand the potential bias of a variety of commonly used computational predictors. MAVEs could mitigate AI bias by producing saturation-style training data for future computational predictors.

The forthcoming new standards, ACMG/AMP/CAP/ClinGen Sequence Variant Guidelines v4, for variant interpretation suggest returning VUS with a high likelihood of pathogenicity to providers for clinical follow-up. We suggest availability of saturation-style MAVE data may help to ensure equitable benefit of this VUS gradation across populations and mitigate any unintentional exacerbation of the current VUS disparity.

Calls for diversifying genomics have yielded a pangenome reference^36^, H3Africa to equip Africa with genomics infrastructure^37^, and diverse participant recruitment in *All of Us*^38^. Diversifying genomics via recruitment, engagement, and retention is just one approach to pursuing equity^39^. MAVEs provide an orthogonal, experimental approach that can complement current sequencing efforts and benefit *All of Us* participants and millions from non-European-like genetic ancestry in global biobanks. MAVEs can scale to the size of the VUS reclassification problem. The saturation-style of MAVE data can also produce equitable training and testing data for future computational predictors. Expansion of MAVE data can spearhead an equitable revolution in genomic medicine for populations previously left on the margins of genetic research.

## Supporting information

All supplementary figures

All supplementary tables

## Data Availability

All input data derived from both gnomAD v2.1.1 and gnomAD v3.1.2 (non v2) is linked at the GitHub above and publicly available at https://gnomad.broadinstitute.org/. All of Us data used in this manuscript is publicly available through the All of Us Data v7 browser (https://databrowser.researchallofus.org/variants) but was accessed via the All of Us Researcher Workbench as Baylor College of Medicine is an approved institution. The project was declared on the All of Us Research Projects Directory in accordance with the All of Us data access model. This declaration can be publicly viewed at https://allofus.nih.gov/protecting-data-and-privacy/research-projects-all-us-data/ ("MAVEComparison"). Both the input data and associated code are accessible through the All of Us workbench and will be promptly shared with requesters with approved workbench access. The code used for analysis of the All of Us data is the same as in the above GitHub with minor modifications made that are specific to the All of Us Researcher Workbench. All the input data used from All of Us is publicly available at the All of Us Public Data Browser (v7; https://databrowser.researchallofus.org/variants). Complete rankings across all three databases of all clinically curated genes by VUS/PorLP/BorLB/CI/ND allele prevalence difference are also linked to the GitHub. All variant reclassifications will be submitted to ClinVar and made publicly available following peer review.

https://databrowser.researchallofus.org/variants

https://github.com/MoezDawood/ReducingVariantClassificationInequities.git

## Acknowledgements

This study originated within the Atlas of Variant Effects (AVE) and was further supported as a cross-consortia project via the Trans-Variant working group of the Impact of Genomic Variation on Function (IGVF) consortia of the United States National Human Genome Research Institute (NHGRI). Additional funding was provided in part by the NHGRI Genomics Research Elucidates Genetics of Rare Disease (BCM GREGoR Center, U01HG011758 for MD, JEP, JRL, RAG), NHGRI IGVF (University of Washington (UW) Center for Actionable Variant Analysis; UM1HG011969 for MD, SF, SP, MP, LAM, DMF, AFR, LMS), NHGRI Centers of Excellence in Genomic Sciences (UW Center for Multiplexed Assessment of Phenotypes; RM1HG010461 for MD, SF, SP, MP, LAM, DMF, AFR, LMS), NHGRI Clinical Genome (ClinGen) Resource (BCM/Stanford ClinGen Resource; U24HG009649 for SEP) and the NIH *All of Us* Program (The Baylor-Hopkins Clinical Genomics Center for *All of Us*; OT2OD002751 for MD, DK, KP, EV, RAG). MD was also supported by the Baylor College of Medicine Comprehensive Cancer Training Program of the Cancer Prevention Research Institute of Texas (CPRITRP210027). CDRE was supported by the Wellcome Trust through a Career Development Award (227228/Z/23/Z), the Melanoma Research Alliance (825924) and the Chan-Zuckerberg Initiative through the Ancestry Networks for the Human Cell Atlas grant program (CZI007). IGR was supported in part by Australian Research Council Discovery Project DP200101552, National Health and Medical Research Council Ideas Grant 2020501 and the European Union through the Horizon 2020 Research and Innovation Program under Grant No. 810645 and the European Union through the European Regional Development Fund Project No. MOBEC008. St Vincent’s Institute acknowledges the infrastructure support it receives from the National Health and Medical Research Council Independent Research Institutes Infrastructure Support Program and from the Victorian Government through its Operational Infrastructure Support Program. We thank Ryan D. Hernandez and Pradiptajati Kusuma for comments on the manuscript.

The *All of Us* Research Program is supported by the National Institutes of Health, Office of the Director: Regional Medical Centers: 1 OT2 OD026549; 1 OT2 OD026554; 1 OT2 OD026557; 1 OT2 OD026556; 1 OT2 OD026550; 1 OT2 OD 026552; 1 OT2 OD026553; 1 OT2 OD026548; 1 OT2 OD026551; 1 OT2 OD026555; IAA #: AOD 16037; Federally Qualified Health Centers: HHSN 263201600085U; Data and Research Center: 5 U2C OD023196; Biobank: 1 U24 OD023121; The Participant Center: U24 OD023176; Participant Technology Systems Center: 1 U24 OD023163; Communications and Engagement: 3 OT2 OD023205; 3 OT2 OD023206; and Community Partners: 1 OT2 OD025277; 3 OT2 OD025315; 1 OT2 OD025337; 1 OT2 OD025276. In addition, the *All of Us* Research Program would not be possible without the partnership of its participants.

## Code and Data Availability

All the code for replicating all the figures and tables in the main text and supplement is readily accessible at https://github.com/MoezDawood/ReducingVariantClassificationInequities.git.

Additionally, all input data derived from both gnomAD v2.1.1 and gnomAD v3.1.2 (non v2) is linked at the GitHub above, and publicly available at https://gnomad.broadinstitute.org/. All of Us data used in this manuscript is publicly available through the All of Us Data v7 browser (https://databrowser.researchallofus.org/variants) but was accessed via the All of Us Researcher Workbench as Baylor College of Medicine is an approved institution. The project was declared on the All of Us Research Projects Directory in accordance with the All of Us data access model. This declaration can be publicly viewed at https://allofus.nih.gov/protecting-data-and-privacy/research-projects-all-us-data/ ("MAVEComparison"). Both the input data and associated code are accessible through the All of Us workbench and will be promptly shared with requesters with approved workbench access. The code used for analysis of the All of Us data is the same as in the above GitHub with minor modifications made that are specific to the All of Us Researcher Workbench. Complete rankings across all three databases of all clinically curated genes by VUS/PorLP/BorLB/CI/ND allele prevalence difference are also linked to the GitHub. All variant reclassifications will be submitted to ClinVar and made publicly available following peer review.

