## Supplementary material for "Defining and Reducing Variant Classification Disparities": All supplementary figures

**Supplementary Appendix 1 for**

**Defining and Reducing Variant Classification Disparities**

Moez Dawood^1-3^*, B.A., Shawn Fayer^4,5^, M.S., Sriram Pendyala^5,6^, B.A., Mason Post^4^, M.S., Divya Kalra^1^, M.S., Karynne Patterson^5^, B.S., Eric Venner^1,2^, Ph.D., Lara A. Muffley^4,5^, B.S., Douglas M. Fowler^4,5,7^, Ph.D., Alan F. Rubin^8,9^, Ph.D., Jennifer E. Posey^2^, M.D., Ph.D., Sharon E. Plon^1,2^, M.D., Ph.D., James R. Lupski^1,2,10,11^, M.D., Ph.D., Richard A. Gibbs^1,2^, Ph.D., Lea M. Starita^4,5^, Ph.D., Carla Daniela Robles-Espinoza^12,13^, Ph.D., Willow Coyote-Maestas^14,15^*, Ph.D., Irene Gallego Romero^16-18^*, Ph.D.

1. Human Genome Sequencing Center, Baylor College of Medicine, Houston, TX, USA.

2. Department of Molecular and Human Genetics, Baylor College of Medicine, Houston, TX, USA.

3. Medical Scientist Training Program, Baylor College of Medicine, Houston, TX, USA.

4. Brotman Baty Institute for Precision Medicine, Seattle, WA, USA.

5. Department of Genome Sciences, University of Washington, Seattle, WA, USA.

6. Medical Scientist Training Program, University of Washington, Seattle, WA, USA.

7. Department of Bioengineering, University of Washington, Seattle, WA, USA.

8. Bioinformatics Division, The Walter and Eliza Hall Institute of Medical Research, Parkville, VIC, Australia.

9. Department of Medical Biology, University of Melbourne, Melbourne, VIC, Australia.

10. Texas Children's Hospital, Houston, TX, USA.

11. Department of Pediatrics, Baylor College of Medicine, Houston, TX, USA.

12. Laboratorio Internacional de Investigación sobre el Genoma Humano, Universidad Nacional Autónoma de México, Campus Juriquilla, Querétaro, Qro, Mexico

13. CASM, Wellcome Sanger Institute, Hinxton, UK

14. Department of Bioengineering and Therapeutic Sciences, University of California, San Francisco, USA

15. Quantitative Biosciences Institute, University of California, San Francisco, USA

16. Human Genomics and Evolution, St Vincent's Institute of Medical Research, Fitzroy, 3065, Australia

17. School of BioSciences and Melbourne Integrative Genomics, The University of Melbourne, Royal Parade, Parkville, 3010, Australia

18. Center for Genomics, Evolution and Medicine, Institute of Genomics, University of Tartu, Riia 23b, 51010, Tartu, Estonia

***Corresponding Authors:**

| **Moez Dawood** | **Willow Coyote-Maestas** | **Irene Gallego Romero** |
| --- | --- | --- |
| | | |
| MD/PhD Student | Assistant Professor | Laboratory Head |
| Human Genome Sequencing Center | Department of Bioengineering | Human Genomics and Evolution |
| Baylor College of Medicine | University of California, San Francisco | St. Vincent’s Institute of Medical Research |

### Supplementary Appendix Table of Contents:

### Supplementary Methods:

Pre-processing of Data from All of Us and gnomAD:

The non-European-like group encompassed individuals with genetic ancestries from the 'African/African American,' 'Latino/Admixed American,' 'East Asian,' 'South Asian,' and ‘Other’ groups as prescribed by the genetic ancestry calculation done by *All of Us* or gnomAD, which we briefly describe here. Full details are available in the respective website documentation and publications of gnomAD (https://gnomad.broadinstitute.org/) and *All of Us* (https://support.researchallofus.org/)^9,18,19^. We use descriptors from *All of Us* and gnomAD for consistency, as we cannot reclassify individuals due to the public, deidentified nature of the databases. In all cases, individuals are assigned to a single genetic ancestry first by projection into a principal component space built from established population genetics resources. Principal component loadings for each individual are then input into a random forest classifier, and the genetic ancestry label is assigned on the basis of the output from the classifier. Given the nature of random forest classifiers, this approach will struggle to assign a label to admixed individuals and to individuals whose genetic ancestry is poorly represented in the reference samples. These individuals, therefore, make up a significant fraction of the "Other" group, which is openly acknowledged by both databases. We use the genetic ancestry labels as prescribed by *All of Us* or gnomAD throughout while acknowledging their imperfect nature as descriptors of continuous human genetic diversity, given our inability to independently reclassify the ancestry of individual samples, but have elected to append the word "like" to the labels to explicitly reflect that they primarily capture genetic similarity to reference groups used by the original publications to train their classifiers.

From gnomAD, allele prevalence for the individuals of European-like genetic ancestry was calculated from the ‘European-like (non-Finnish)’ group. Due to the high degrees of consanguinity in the Finnish and Ashkenazi Jewish populations, these two populations were not included in our analysis. Allele counts, frequencies, population descriptors, ClinVar clinical significance calls, and number of individuals sequenced in each population were used as prescribed by *All of Us* or gnomAD as of June 2023. All clinical significance calls were mapped to one of six categories: ‘Pathogenic or Likely Pathogenic,’ ‘Benign or Likely Benign,’ ‘Variant of Uncertain Significance,’ ‘Conflicting Interpretations,’ ‘Not Included,’ or ‘No Designation’ based on their current ClinVar Clinical Significance designation as specified in *All of Us* or gnomAD. Trends were pinpointed if shown to be consistent across all three databases. Due to differences in extraction of ClinVar data between gnomAD and *All of Us*, there are systematic database level differences that potentially are unaccounted for. In these instances, the GenCC list of all curated clinical genes being the biggest and most comprehensive list is used as the main indicator of a trend. gnomAD version 2.1.1 and version 3.1.2, non-v2 (removes individuals overlapping between v2 and v3) were treated as two independent population databases^18,19^.

Gene Lists*:*

Gene lists for medical specialties that commonly use genetic testing were compiled from genes known to be tested on next-generation sequencing tests of Invitae, Ambry Genetics, and Baylor Genetics. The ACMG78 gene list represents the 78 genes from the secondary findings list curated per the American College of Medical Genetics Secondary Findings v3.2 standard. The GenCC gene list represents all 4640 curated known clinical disease genes (<https://thegencc.org/>; as of June 2023). The ‘Cancer’ gene list represents 209 genes implicated in hereditary cancers and cancer syndromes across every major organ system. The ‘Cardiac’ gene list represents 306 genes implicated in arrhythmias, cardiomyopathies, RASopathies, congenital heart diseases, lipidemias, and aortopathies. The ‘Hematology’ gene list represents 240 genes implicated in benign and malignant blood disorders such as inherited platelet disorders and thrombocytopenias, anemias, enzymopathies, red blood cell membrane disorders, telomere disorders, bone marrow failure, and more. The ‘Newborn screening’ gene list represents 1755 genes implicated in inherited metabolic disorders. The ‘Carrier Screening’ gene list represents 568 genes commonly examined to understand if there is an increased risk of having a child affected with a genetic condition. The ‘Endocrinology’ gene list represents 321 genes implicated in disorders of sex development, obesity, thyroid and parathyroid conditions, bone mineralization disorders, and glucose metabolism. The ‘Immunology’ gene list represents 572 genes implicated in primary immunodeficiency, telomere biology disorders, antibody deficiencies, autoinflammatory syndromes, B and T cell deficiencies, phagocytic defects, hereditary angioedema, complement deficiencies, and congenital diarrhea. The ‘Nephrology’ gene list represents 565 genes implicated in ciliopathies, nephrolithiasis, progressive renal disease, rare clinical syndromes with renal manifestations, atypical hemolytic uremic syndrome, and thrombotic microangiopathies. The ‘Neurology’ gene list represents 1374 genes implicated in neuropathies, movement disorders, neurodegenerative disorders, neurovascular disorders, epilepsy disorders, seizure disorders, neurodevelopmental disorders, and neuromuscular disorders. The ‘Ophthalmology’ gene list represents 514 implicated in blindness are rare disorders affecting vision, the eye, and/or the retina. The DDG2P gene list representing the curated list of 2307 genes reported to be associated with developmental disorders from the DECIPHER project was accessed in June 2023. The ‘SGE’ gene list represents the 694 genes that are both essential in HAP1 cells and found in the GenCC gene list. The ‘VAMPseq’ gene list represents the 394 genes that are both high priority for VAMPseq and found in the GenCC gene list. The high priority VAMPseq genes were selected because their proteins are not secreted extracellularly, thermostable, have previously been shown to be GFP tagged, and are monomeric. The ‘MAVERegistry’ list was determined based on the 110 genes as of August 2023 that are either ‘Under Investigation’ or in the ‘MAVE Data Collection’ phases on the MAVERegistry (<https://registry.varianteffect.org>). When appropriate, the same gene may be found in more than one gene list (for example, *BRCA2* would be found in the Oncology, GenCC, ACMG78, SGE, and MAVERegistry lists). Overall, all gene lists used in this study are available in Table S1.

Wilcoxon matched-pairs signed-rank test*:*

We employed a matched pairs signed rank Wilcoxon test, with the matched pairs based on the gene itself and its allele prevalence between individuals of non-European-like versus European-like genetic ancestry. This gene-by-gene comparison mitigates any other confounders such as gene length, coverage during sequencing, and other gene-specific intricacies that are canceled out by comparing the allele prevalence within the non-European-like group to the allele prevalence in the European-like group within each gene. The ranking aspect of the test is crucial, as it does not presuppose a uniform trend of larger allele prevalence in the non-European-like group compared to the European-like group across all genes for every clinical significance allele type. By ranking the genes prior to the statistical test, we incorporate genes that have a higher number of alleles in Europeans into our analysis, ensuring a complete survey of the allele prevalence in all genes in the statistical test. The difference in allele prevalence and difference in unique VUS between the non-European-like group and European-like group was also used to rank the genes with the greatest VUS disparity between non-Europeans vs. Europeans.

While the p-value informs us whether or not there is a difference, we then calculated the rank biserial coefficient (r) with a 95% confidence interval to quantify the magnitude of the statistically significant differences. This calculation was performed using Python-wrapped R code, employing the ggwithinstats function from the ggstatsplot library and the effectsize library, with settings based on thresholds outlined by Funder and Ozer (2019)^21^. The resultant coefficient categories are based on the magnitude of r < 0.05 – Tiny; 0.05 ≤ r < 0.1 - Very small; 0.1 ≤ r < 0.2 – Small; 0.2 ≤ r < 0.3 – Medium; 0.3 ≤ r < 0.4 – Large; r ≥ 0.4 - Very large. Additionally, we evaluated the statistical power of each Wilcoxon matched-pairs signed-rank using a simulation-based approach. The simulation iterates 50,000 times to generate matched pair samples under a normal distribution, with the first sample being the control and the second sample being offset by the defined effect size. Each iteration performs the Wilcoxon signed-rank test to assess the significance of the observed effect based on the Bonferroni-corrected alpha. The proportion of 50,000 iterations yielding significant results was the estimate of statistical power, reflecting the test’s ability to correctly reject the null hypothesis for a specified effect size and sample size.

We also assessed the overlap in variants between the Non-European-like and European-like groups. This involved calculating the number of variants present in both groups, as well as the number of variants unique to each group, and expressing these as percentage contributions. In contrast to the below orthogonal statistical method, all variants, including those shared between groups, were retained for the Wilcoxon matched-pairs signed-rank test to ensure that any unique variant’s prevalence in both populations was duly considered in assessing potential differences. All code has been made freely available and is linked in the Code Availability section.

Chi-square test for independence:

Furthermore, we employed a chi-square test for independence to investigate the presence of unique variants in each population group. In contrast to the above orthogonal statistical method, variants found in both groups were removed for the Chi-square test for independence to examine prevalence differences of variants found exclusively in the European-like versus non-European-like genetic ancestry groups with an accompanying power estimate. Instead of the gene-by-gene approach, this approach allowed us to systematically assess three population databases, seeking to determine whether there is a consistent higher count of unique variants (not allele count) across different medical specialties and gene groups. Further, it helps to satisfy the requirement of independence of observations for the chi-square test as there are no relationships between the counts in the individual medical specialty groups and no pairing of the data between the super populations.

It is worth noting that neither the Wilcoxon test nor the chi-square test for independence necessitates an underlying distribution that approximates normality. Visual inspection of variant prevalence in the GenCC data revealed that the distributions of variants best resembles a chi-square distribution. Thus, the chi-square test, based on the chi-square data distribution, is particularly suitable for modeling our data. All code has been made freely available and is linked in the Code Availability section.

#### Bonferroni Corrections:

To counteract the potential for type I errors due to multiple comparisons, we apply a stringent Bonferroni correction to each statistical test. For testing the difference in allele prevalence of all coding variants of a particular clinical significance type across specialties, there are 14 specialties x 3 databases x 5 clinical significance groups = 210 total tests. For testing the difference in allele prevalence of all coding variants without missense variants of a particular clinical significance type across specialties, there are also 14 specialties x 3 databases x 3 clinical significance groups = 126 total tests. For testing the difference in allele prevalence of variant types for different clinical classifications for the GenCC curated genes list, there are 11 variant types x 3 databases x 5 clinical significance categories = 165 statistical tests. For testing the difference in allele prevalence of coding variants of a particular clinical significance type across population distributions for the GenCC curated genes list, there are 1 specialty x 3 databases x 5 clinical significance groups x 5 pairwise population comparisons = 75 total tests. For testing the difference in allele prevalence of noncoding variants of a particular clinical significance type across specialties, there are 14 specialties x 2 databases x 5 clinical significance groups = 140 total tests. For testing the difference in unique variants of a particular clinical significance found only in one population group via chi-square testing, there are 3 databases x 5 clinical significance groups = 15 total tests. Of particular note, because the three research questions are independent of each other (e.g. no nested hypotheses, no repeated measures, no sequential testing) and the underlying data distributions for each statistical test are very different for the three questions each group of tests received its own Bonferroni correction.

Variant Reclassification:

Initially, each variant was annotated using the 2015 ACMG (American College of Medical Genetics and Genomics) evidence codes through the Intervar API. During this process, we ensured that the correct reference genomes were used for the different databases (*All of Us* and gnomAD v3.1.2 utilized GRCh38; whereas gnomAD 2.1.1 utilized GRCh37). Following this initial annotation, each variant was further annotated with functional scores from MAVE data. The clinical curation and clinical strength assignment as per the ClinGen recommendations in Brnich et al. (2020)^40^ for or against pathogenicity or benignity of each of these MAVE datasets utilized in this study were previously published in Fayer et al. (2021)^13^. In brief, for *BRCA1* variants, if a variant was categorized as FUNC (functional), it was assigned BS3 evidence and no PS3 evidence, whereas if it was categorized as LOF (loss of function), the variant was assigned PS3 evidence and no BS3 evidence. Variants categorized as INT (intermediate) were left unannotated. For the BRCA1 combining criteria, ≥ 1 criteria of strong benign evidence was enough to reclassify the VUS as Likely Benign. For *TP53*, we used the output of the Naïve Bayes classifier that synthesized data from four different *TP53* MAVEs in Fayer et al. (2021). If the classifier predicted a variant to be "Functionally abnormal," the variant was assigned PS3 evidence and no BS3 evidence. If a variant was predicted to be "Functionally normal," BS3_moderate evidence was used with no PS3 evidence. For PTEN, two assays measuring activity and abundance were used. If the abundance was categorized as "wt-like" or "possibly wt-like," BS3_Supporting evidence was used. Furthermore, if the cumulative score was greater than -5, BS3_moderate evidence was used. All other evidence codes and combining criteria were adhered to as closely as possible based on the ClinGen VCEP (Variant Curation Expert Panel) gene-specific recommendations for *BRCA1*, *TP53*, and *PTEN*, respectively. The ClinGen VCEPs are highly regarded as the gold-standard for gene-specific variant curation and are developed after extensive evaluation of the evidence by clinical and scientific experts for the particular gene to classify genomic variants on a spectrum from pathogenic to benign using the 2015 ACMG/AMP Variant Interpretation Guidelines as a backbone^29^. Reclassification of variants from gnomAD or *All of Us* focused only on variants originally prescribed as VUS to ensure that we were only reclassifying variants that had been previously seen in the context of an individual with a suspected clinical phenotype relevant to the gene. To ensure reproducibility, transparency, and increased throughput, all the procedures for annotating variants and assigning evidence codes were codified using Python. All code has been made freely available and is linked in the Code Availability section and all reclassified variants with evidence codes used can be found in Tables S18-19.

### Supplementary Results

#### Coding Variants

Current variant interpretation standards, focused on coding variants, still require expansion and refinement. For well-understood variant types such as stop gains, frameshifts, and canonical splice variants, our existing knowledge base is substantial enough that we do not observe a significant disparity in VUS classification between the non-Europeans-like and European-like groups. However, when classifying challenging synonymous, inframe indels, splice region and missense variants, our current interpretation of coding variants falls short in preventing VUS disparities between non-Europeans and Europeans. This gap in knowledge could be potentially addressed by MAVEs which are able to systematically ascertain a functional effect for each of these coding variant types.

#### Noncoding Variants

We were unable to assess the *All of Us* database for noncoding variants at this time due to cost restrictions, but we did evaluate noncoding variants in gnomAD v2.1.1 and gnomAD v3.1.2 (non v2) for any potential disparities in VUS (**Fig.SF7**) or P/LP (**Fig.SF8**) assignment. However, there are so few genes with known pathogenic noncoding variants (**Fig.SF8**) that this analysis is severely underpowered and premature at this time. This limitation can be attributed to our relatively nascent and limited but evolving understanding of how noncoding variants contribute to pathogenicity. While there may be initial hints of a VUS disparity based on the GenCC list in gnomAD v2 (**Fig.SF7**), further research is needed to even establish enough of a data distribution for analysis. While our current study did not reveal widespread disparities in noncoding variant classifications, as more noncoding variants need clinical interpretation, it is important to avert a genome-scale variant classification bias between different populations. Further mechanistic understanding of noncoding and more challenging coding variants where VUS disparity was found is necessary to ensure consistent and accurate interpretation across all populations.

#### Effect Sizes

After identifying significant differences in allele prevalence among the different clinical variant classifications, we examined the effect sizes of the Wilcoxon matched pairs signed rank tests to quantify these differences. Effect size thresholds, set by Funder and Ozer (2019)^21^, were used to describe the magnitude of differences from ‘small’ to ‘very large.’ In contrast to VUS, pathogenic and benign variants (**Fig.SF17****-18**) showed medium to large effect sizes (>0.2). Removing missense variants had minimal effect on the effect sizes of either P/LP or B/LB variants, suggesting that the disparity in these categories is largely driven by other variant types, not missense variants.

#### Greater Diversity of Unique Coding Variants in Individuals of non-European-like Genetic Ancestry At Baseline

Our findings align with previous research, underscoring the greater diversity of unique coding variants in non-European-like populations compared to individuals of European-like genetic ancestry. This observation is supported on a gene-by-gene basis by the significant increased allele prevalence in both B/LB variants and ND variants among non-European-like individuals when compared to Europeans for both coding and noncoding variants (**Fig.S4-5****,** **Fig.S9****,****11**). Moreover, using the orthogonal statistical method that focuses on comparing unique variants between individuals of non-European-like versus European-like genetic ancestry, our study consistently reveals a significantly greater count of B/LB and ND unique variants in non-Europeans (**Fig.S26**). Examining each of the five genetic ancestries (African/African-American, Latino/Admixed American, South Asian, East Asian, and Other) in pairwise comparisons with the European-like genetic ancestry group, each of these genetic ancestries displays a significant increased prevalence of variants with no designation, and several also show elevated prevalence of B/LB variants (**Fig.S27**). This trend is reinforced when examining the data by variant types. For non-designated (ND) variants, all coding and noncoding variant types exhibit significant increases in allele prevalence among non-European-like genetic ancestries, while several variant types also demonstrate heightened prevalence in non-European-like populations for benign variants (**Fig.S21****,****23-24**). These findings collectively establish a baseline depiction of the greater diversity of unique coding variants among the non-European-like superpopulation compared to European-like.

#### Comparison to Fayer et al. (2021):

We comprehensively reanalyzed the set of *BRCA1*, *PTEN*, and *TP53* VUS previously reclassified by Fayer et al. (2021)^13^ (Supplemental Tables 7, 10, 11 in Fayer et al. (2021)). The automated pipeline uses VCEP recommendations as of Fall 2023; however, the Fayer et al. (2021) VUS dataset was analyzed by hand with a mix of VCEP and ACMG/AMP 2015 recommendations prior to 2021. Using this dataset, we sought to establish a robust benchmark for the automated variant classification pipeline built for this project to ensure clinical variant classifications ascertained by the automated pipeline were concordant with the Fayer et al. (2021) reclassifications where MAVE data was also used for variant classification. We defined a concordant classification as a final clinical classification on the same side of pathogenicity as was found in the Fayer et al. (2021) dataset (the groups being Benign or Likely Benign versus Pathogenic or Likely Pathogenic versus remaining a VUS). Further, we used this dataset to follow-up on the essential code analysis. We annotated all possible variants in this dataset with allele frequency data from gnomAD v4 (not using v3 or v2 to prevent accidental double-dipping). gnomAD v4 is not an ideal dataset for this analysis as >80% of the individuals sequenced in gnomAD v4 are of European-like genetic ancestry (**Fig.S1**). However, even still we were able to confirm the significant inequitable impact of the allele frequency and computational predictor evidence codes in the Fayer et. al (2021) dataset (**Fig.S32**, Table S19).

### Supplemental Figure 1: Comparison of Genetic Ancestry Group Sizes in *All of Us* and gnomAD

(a) The pie charts show the percentage breakdown of individuals from each genetic ancestry group relative to the total number of individuals in that particular database. (b) Bar graph comparing number of individuals across various groups based on precomputed genetic ancestry in the three major sequencing databases: *All of Us* v7 (blue), gnomAD v2.1.1 (green), and gnomAD v3.1.2 (non v2) (red). Each bar represents a group, and the height of the bar indicates the number of individuals from that group present in the database. (c) Number of individuals used for each comparison throughout this study. Individuals of non-European-like genetic ancestry include individuals attributed as ‘African/African American,’ ‘Latino/Admixed American,’ ‘East Asian,’ ‘South Asian,’ and ‘Middle Eastern’ as attributed by either *All of Us* or gnomAD. For the three population databases chosen for this study, *All of Us* v7, gnomAD v2.1.1, and gnomAD v3.1.2 (non v2), all three have rough an equal number of individuals of European-like and non-European-like genetic ancestry. Although on average there are still ~2% more individuals of European-like genetic ancestry in each database than non-European-like. For this reason, this analysis was not pursued in gnomAD v4 nor the UK Biobank, because both of these databases have many more individuals of European-like genetic ancestry than non-European-like. The reference genome used for alignment and variant calling was GRCh38 for *All of Us* v7 and gnomAD v3.1.2 (non v2) versus GRCh37 for gnomAD v2.1.1. The NGS assay used was whole genome sequencing for *All of Us* v7 and gnomAD v3.1.2 versus whole exome sequencing for gnomAD v2.1.1 (although a few whole genomes are also present in gnomAD v2.1.1).

**Supplemental Figure 1a: Pie Charts of Distribution of Individuals in Each Population Database Based On Genetic Ancestry**

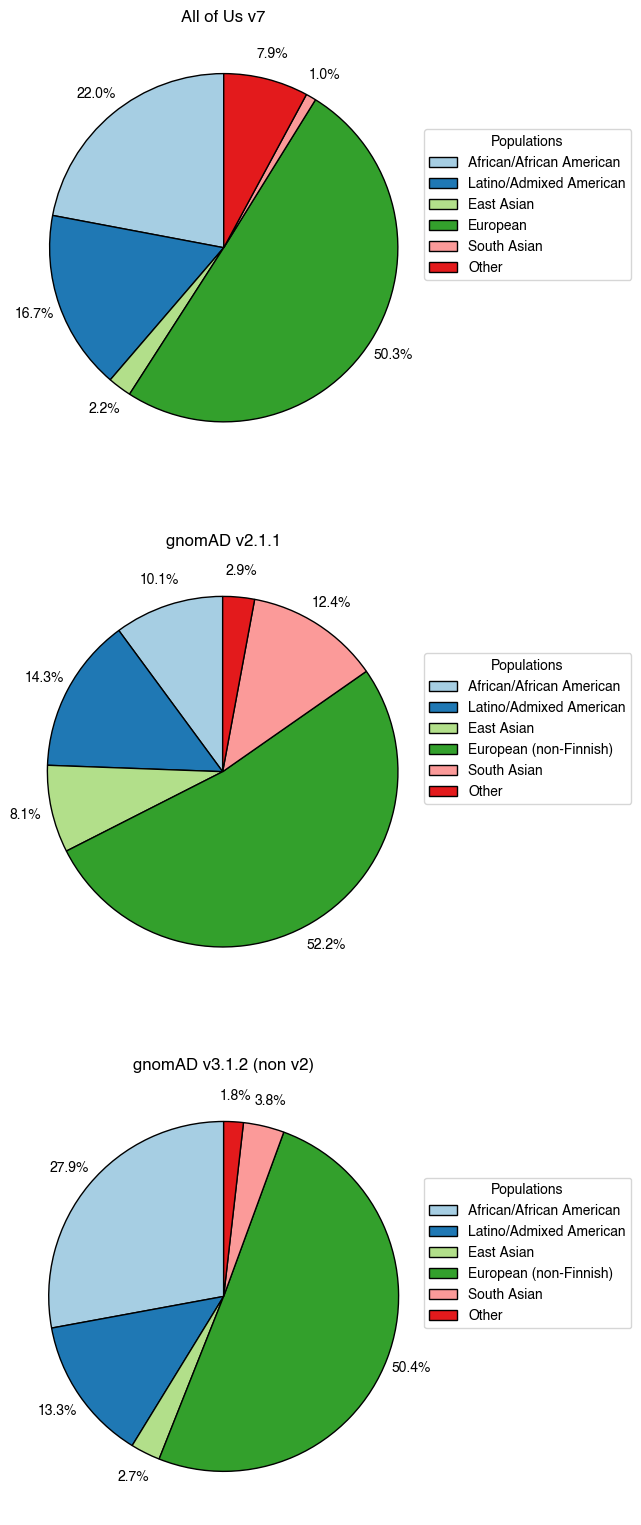

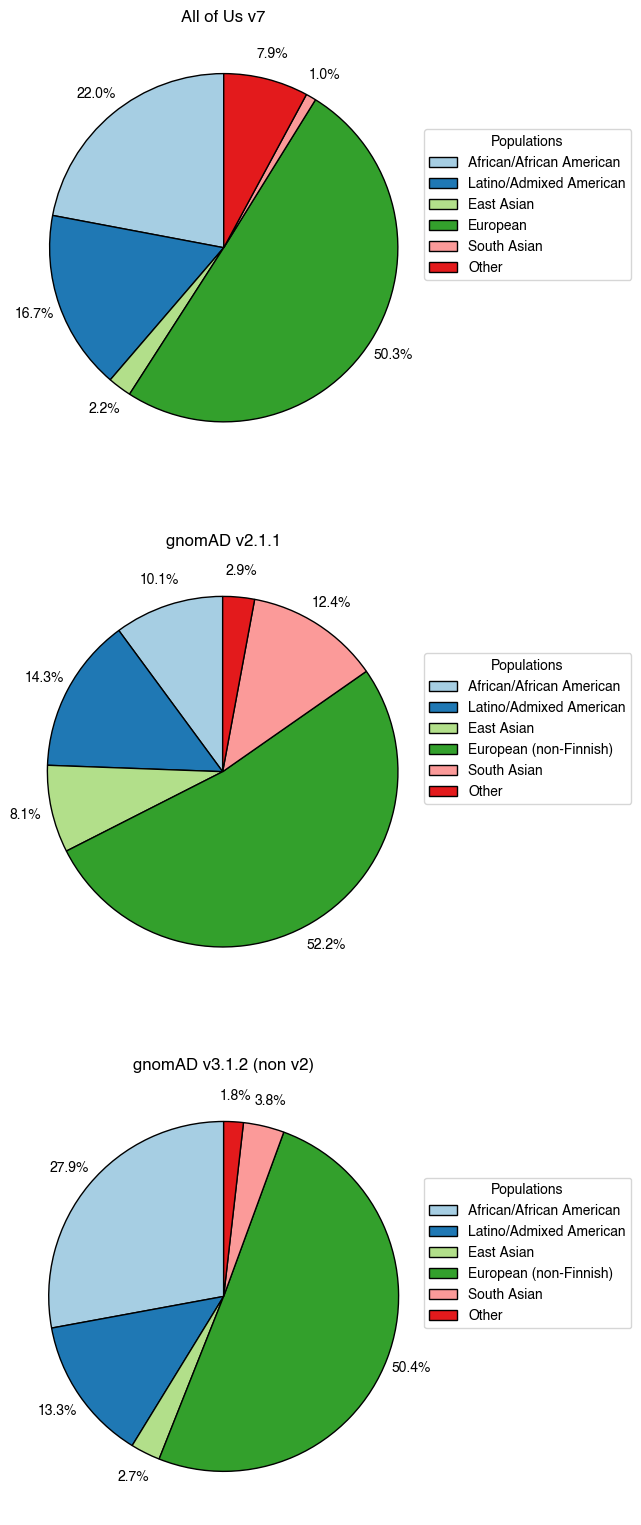

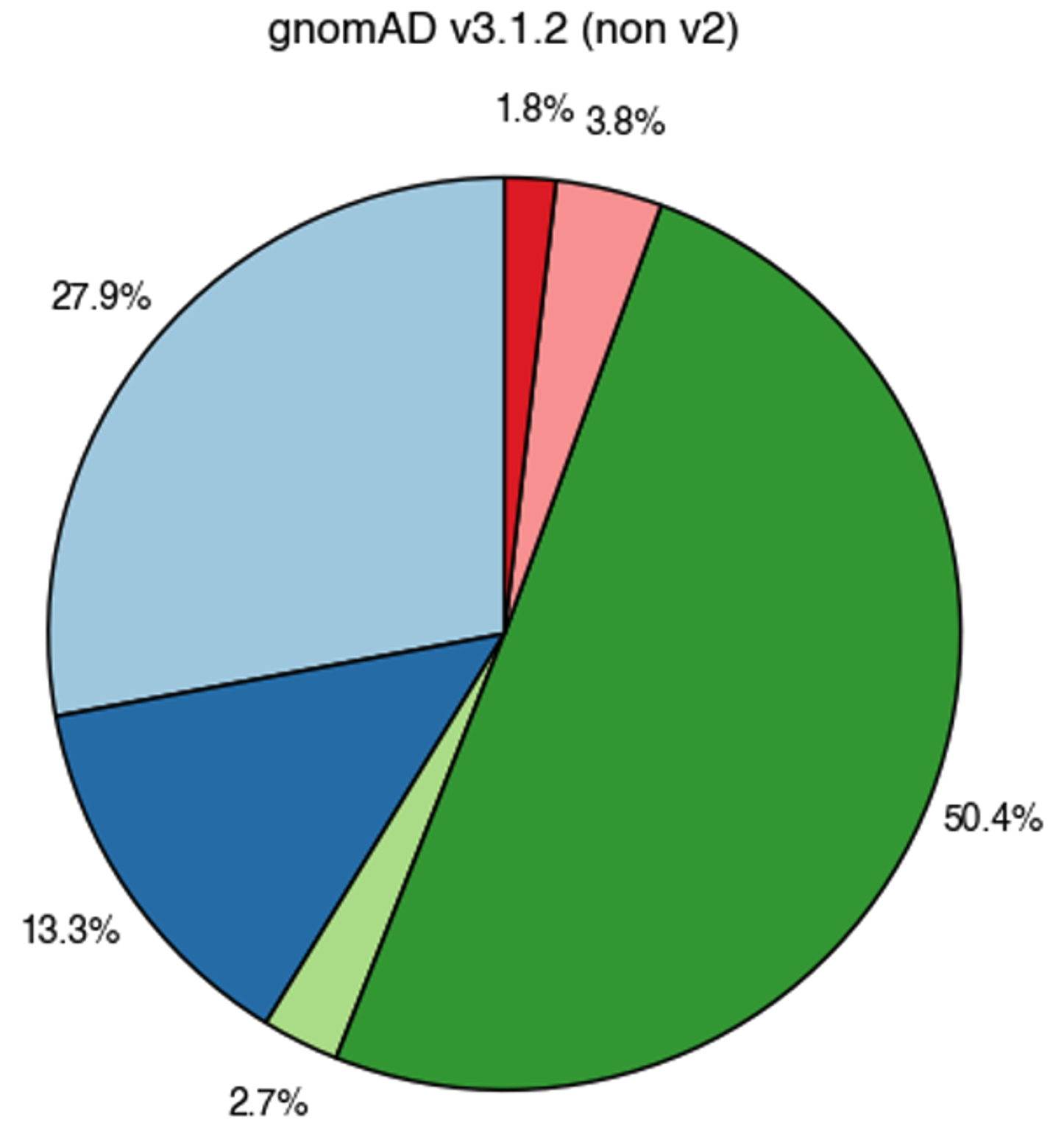

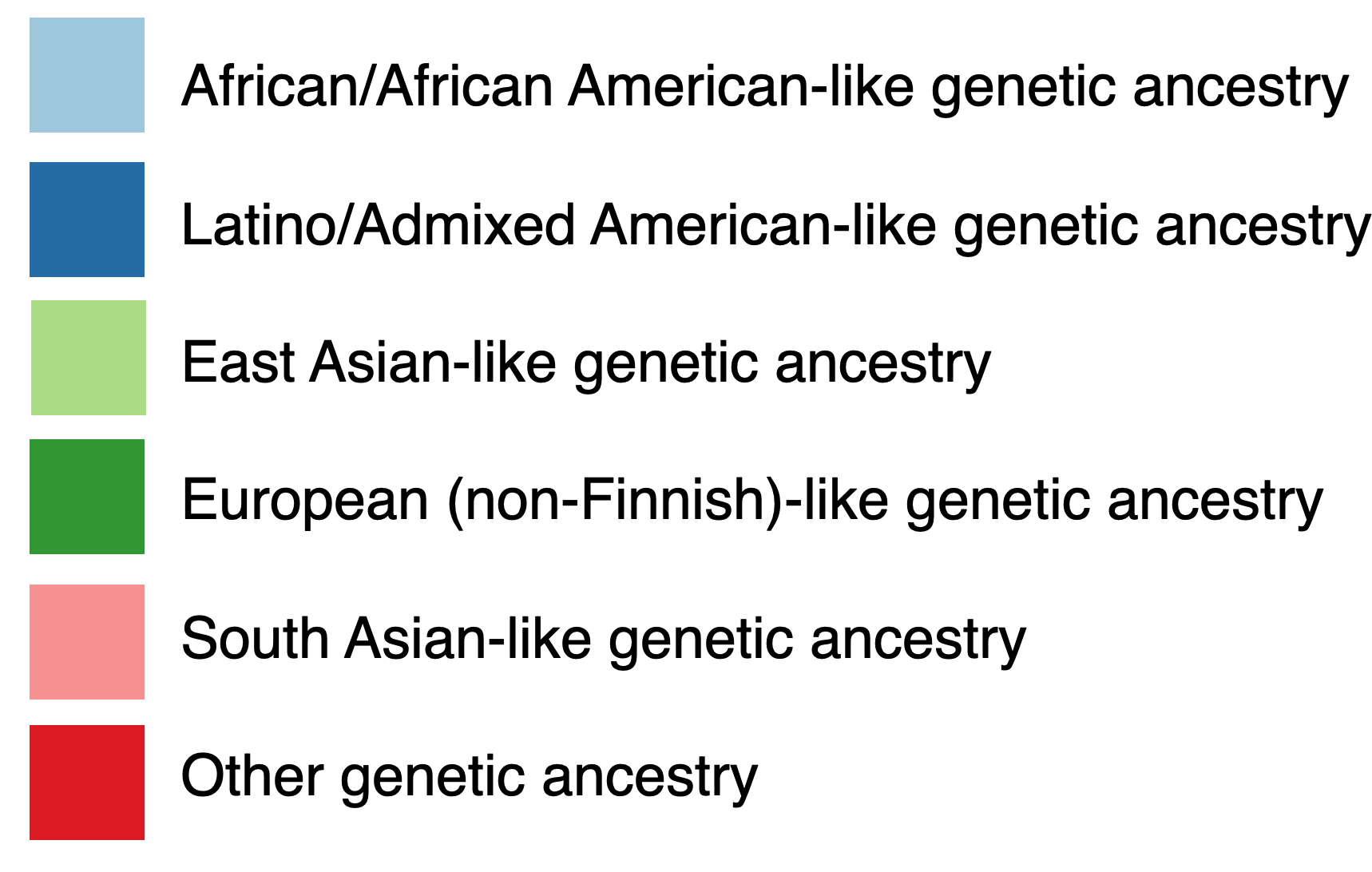

**Supplemental Figure 1b: Comparison of Individuals Across Populations Based on Genetic Ancestry**

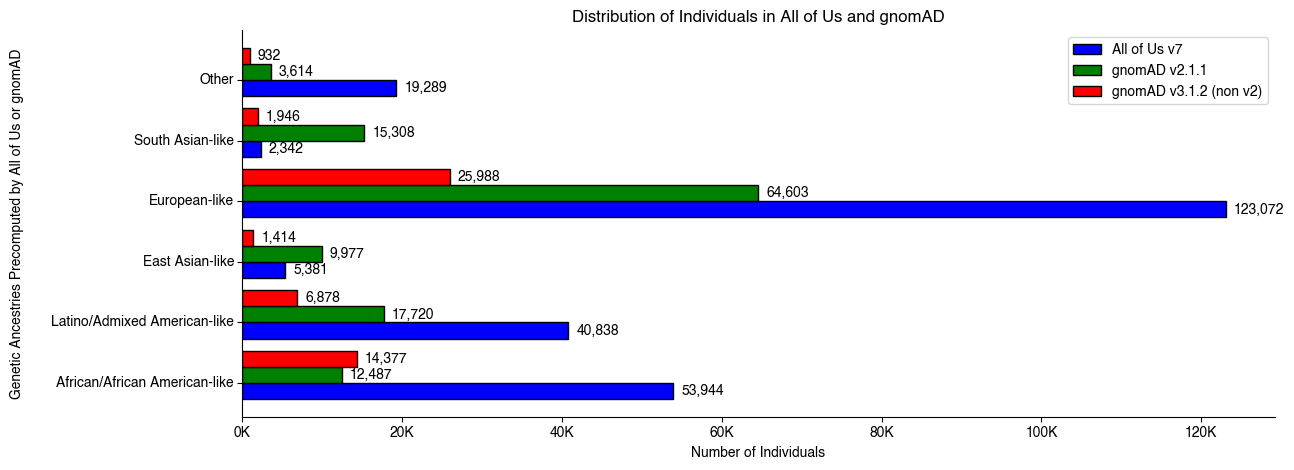

**Supplemental Figure 1c: Cumulative Number of Individuals of European-like Versus Non-European-like Genetic Ancestry**

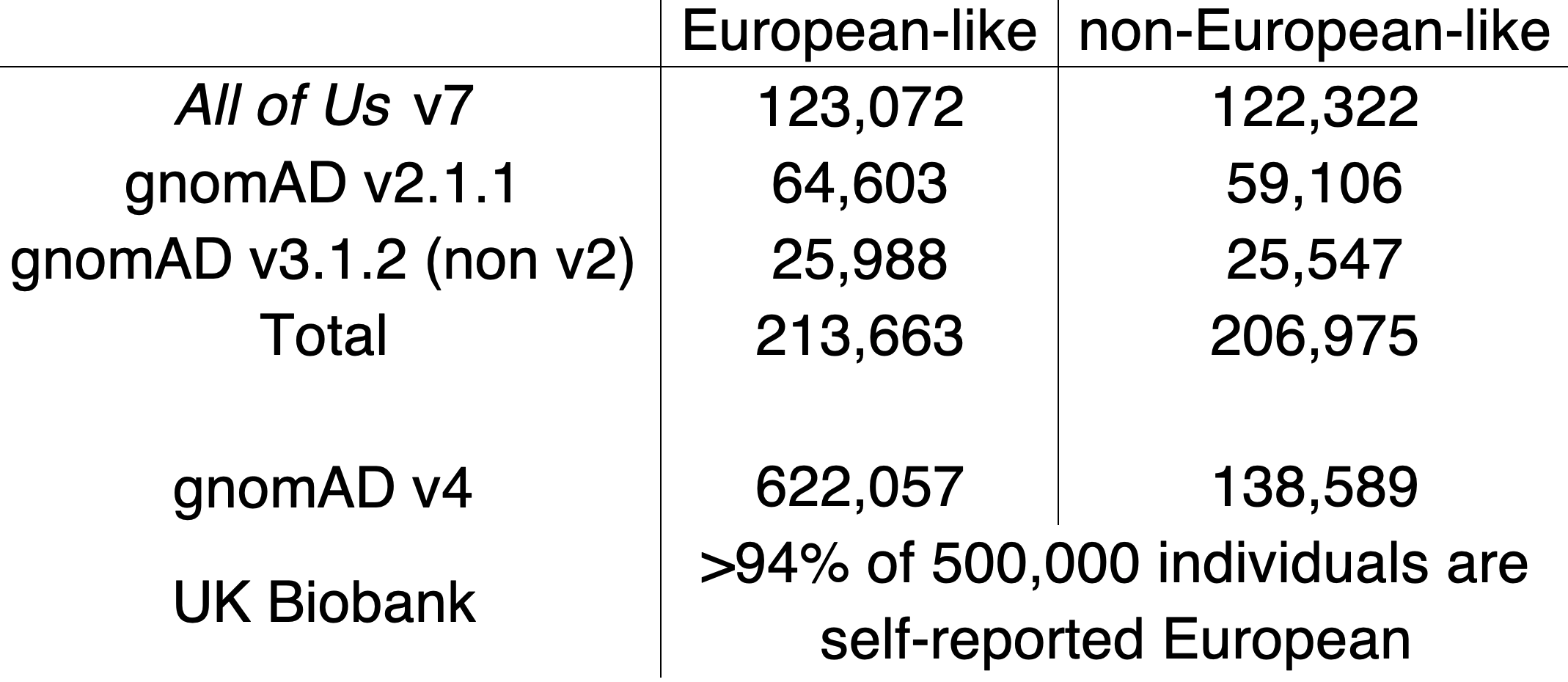

### Supplemental Figure 2: Allele Prevalence Comparison of Variants of Uncertain Significance (VUS)

Box plots corresponding to VUS allele prevalence (x-axis) in each gene (dot) for individuals of non-European-like (blue) versus European-like (orange) genetic ancestry for the corresponding medical specialty (y-axis) across a) gnomAD v2.1.1 and b) gnomAD v3.1.2 (non v2) for all coding variants. Genes with zero alleles for allele prevalence for either individuals of European-like or non-European-like genetic ancestry are omitted from the above visualization to maintain a reasonable scale for data visualization. However, genes with zero alleles for only one category of either individuals of European-like or non-European-like genetic ancestry are included in the Bonferroni-corrected, signed rank, matched pairs Wilcoxon statistical test. The Bonferroni corrected p-values associated with these comparisons are annotated as follows with "ns" indicating not significant, * for 1.19e-04 < p ≤ 2.38e-04, ** for 5.95e-05 < p ≤ 1.19e-04, *** for 5.95e-06 < p ≤ 5.95e-05, and **** for p ≤ 5.95e-06. Also refer to Tables S2-4. Across all medical specialties and categories shown, VUS are observed to be significantly increased in individuals of non-European-like genetic ancestry compared to individuals of European-like genetic ancestry.

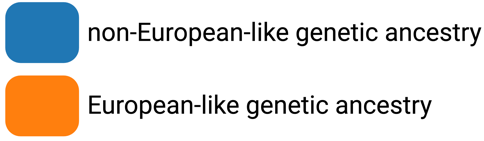
**Supplemental Figure 2a: Allele Prevalence Comparison of VUS in gnomAD v2.1.1**

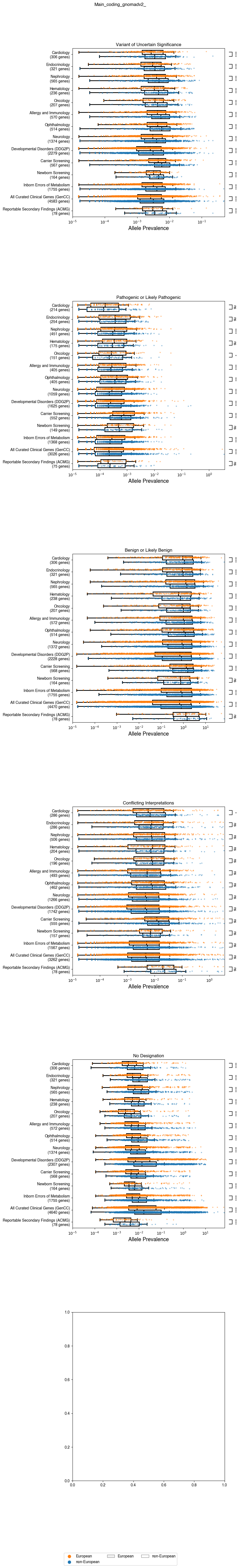

**Supplemental Figure 2b: Allele Prevalence Comparison of VUS in gnomAD v3.1.2 (non v2)**

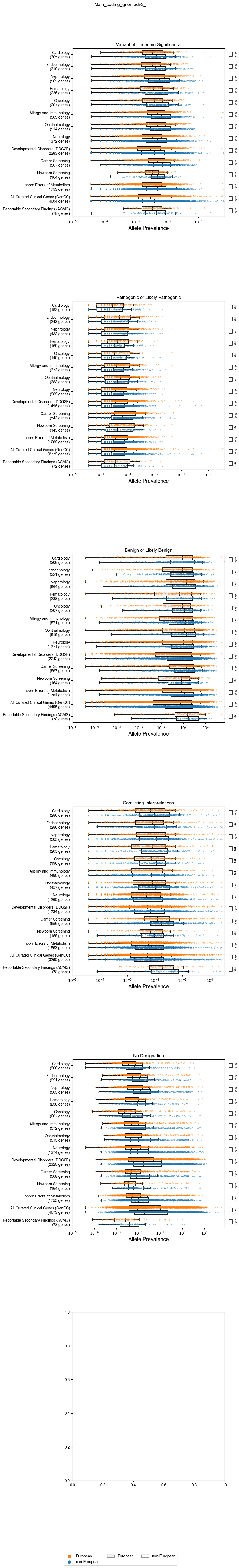

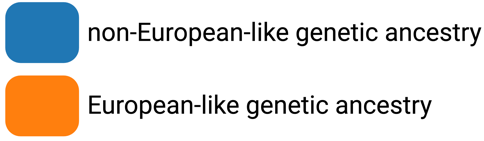

### Supplemental Figure 3: Allele Prevalence Comparison of Pathogenic or Likely Pathogenic (P/LP) Variants

Box plots corresponding to P/LP allele prevalence (x-axis) in genes (dots) for individuals of non-European-like (blue) versus European-like (orange) genetic ancestry for the corresponding medical specialty (y-axis) across a) *All of Us* v7, b) gnomAD v2.1.1, and c) gnomAD v3.1.2 (non v2) for all coding variants. Genes with zero alleles for allele prevalence for either individuals of European-like or non-European-like genetic ancestry are omitted from the above visualization to maintain a reasonable scale for data visualization. However, genes with zero alleles for only one category of either individuals of European-like or non-European-like genetic ancestry are included in the Bonferroni-corrected, signed rank, matched pairs Wilcoxon statistical test. The Bonferroni corrected p-values associated with these comparisons are annotated as follows with "ns" indicating not significant, * for 1.19e-04 < p ≤ 2.38e-04, ** for 5.95e-05 < p ≤ 1.19e-04, *** for 5.95e-06 < p ≤ 5.95e-05, and **** for p ≤ 5.95e-06. Also refer to Tables S2-4. Overall, in the set of curated clinical genes as a whole (GenCC), we observe a significant increase in P/LP variants in individuals of European-like genetic ancestry compared to individuals of non-European-like genetic ancestry. However, this trend varies across specialties and gene lists across the three databases.

**Supplemental Figure 3a: Allele Prevalence Comparison of P/LP Variants in *All of Us* v7**

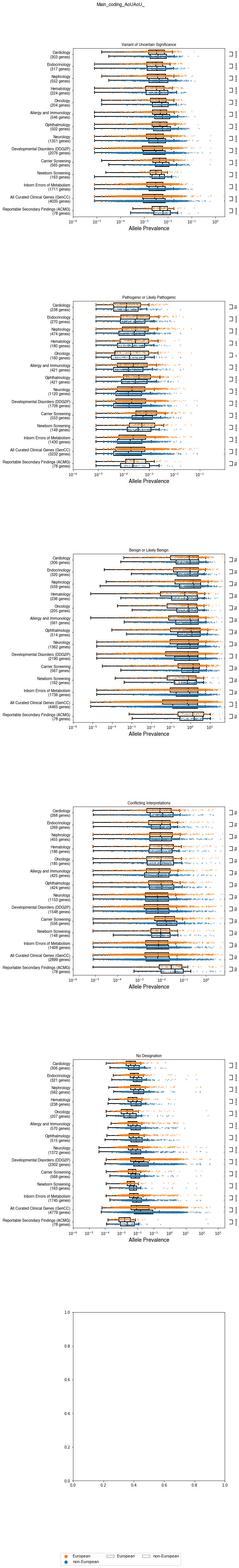

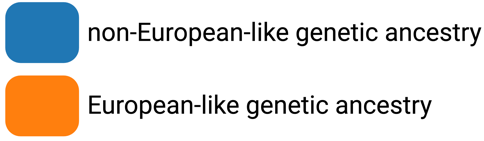

**Supplemental Figure 3b: Allele Prevalence Comparison of P/LP Variants in gnomAD v2.1.1**

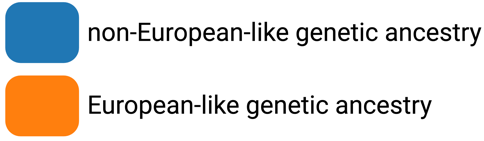

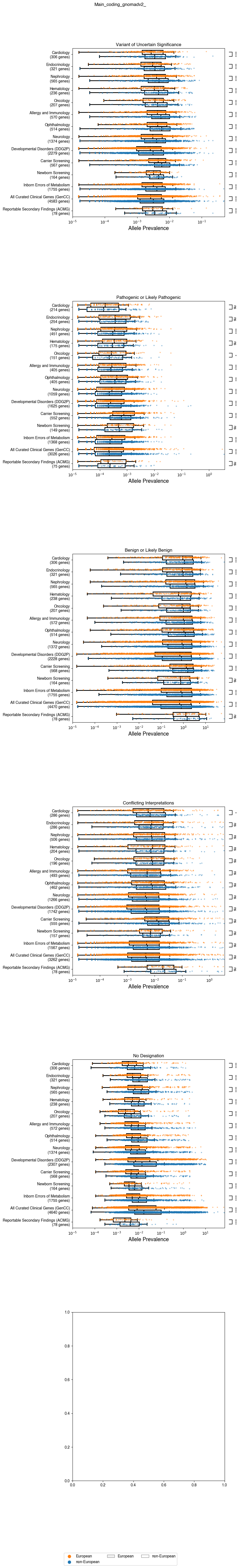

**Supplemental Figure 3c: Allele Prevalence Comparison of P/LP Variants in gnomAD 3.1.2 (non v2)**

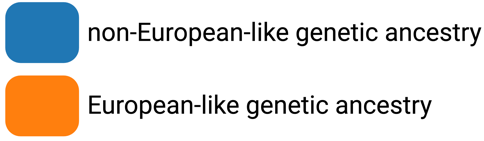

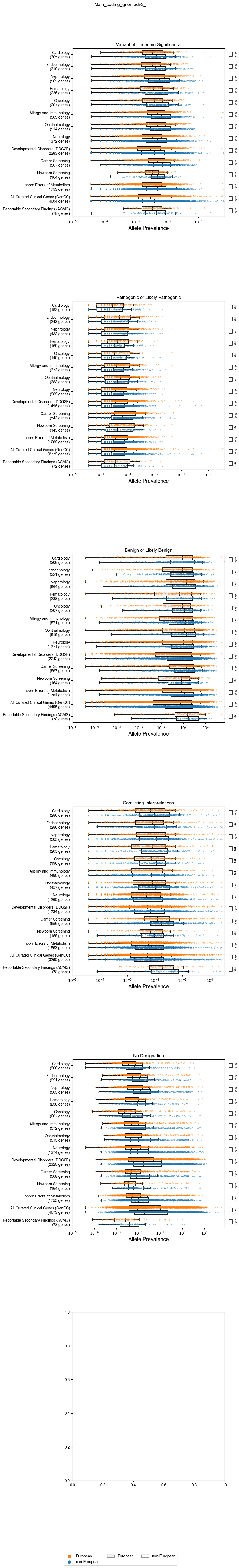

### Supplemental Figure 4: Allele Prevalence Comparison of Benign or Likely Benign (B/LB) Variants

Box plots corresponding to B/LB allele prevalence (x-axis) in genes (dots) for individuals of non-European-like (blue) versus European-like (orange) genetic ancestry for the corresponding medical specialty (y-axis) across a) *All of Us* v7, b) gnomAD v2.1.1, and c) gnomAD v3.1.2 (non v2) for all coding variants. Genes with zero alleles for allele prevalence for either individuals of European-like or non-European-like genetic ancestry are omitted from the above visualization to maintain a reasonable scale for data visualization. However, genes with zero alleles for only one category of either individuals of European-like or non-European-like genetic ancestry are included in the Bonferroni-corrected, signed rank, matched pairs Wilcoxon statistical test. The Bonferroni corrected p-values associated with these comparisons are annotated as follows with "ns" indicating not significant, * for 1.19e-04 < p ≤ 2.38e-04, ** for 5.95e-05 < p ≤ 1.19e-04, *** for 5.95e-06 < p ≤ 5.95e-05, and **** for p ≤ 5.95e-06. Also refer to Tables S2-4. Overall, there seems to be a systematic difference in B/LB allele prevalence between the *All of Us* database versus the gnomAD database as only the GenCC list is significant in the *All of Us* data versus the majority of the medical specialties and gene lists are significant in both gnomAD databases. However based on the GenCC list being the set of all curated clinical genes, we do observe a significant increase in B/LB variants in individuals of non-European-like genetic ancestry compared to individuals of non-European-like genetic ancestry across all three databases.

**Supplemental Figure 4a: Allele Prevalence Comparison of B/LB Variants in *All of Us* v7**

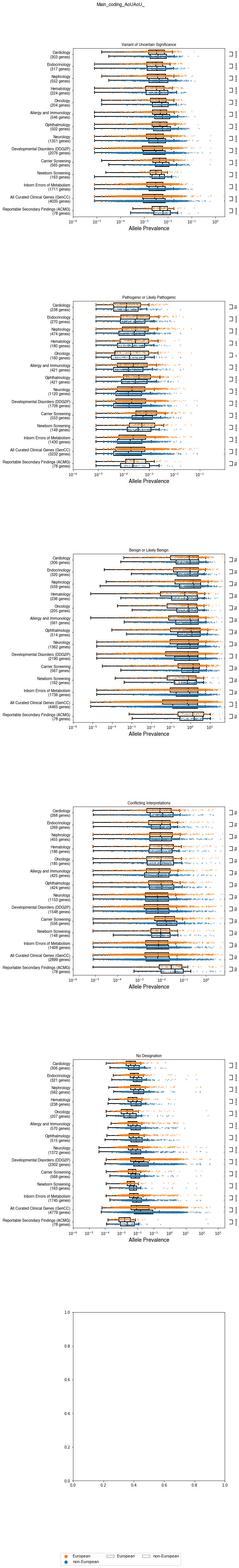

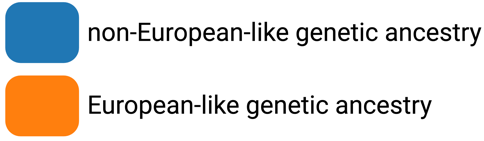

**Supplemental Figure 4b: Allele Prevalence Comparison of B/LB Variants in gnomAD v2.1.1**

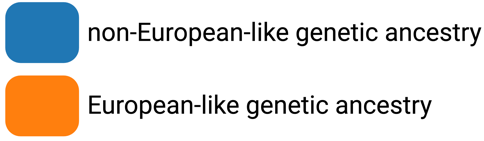

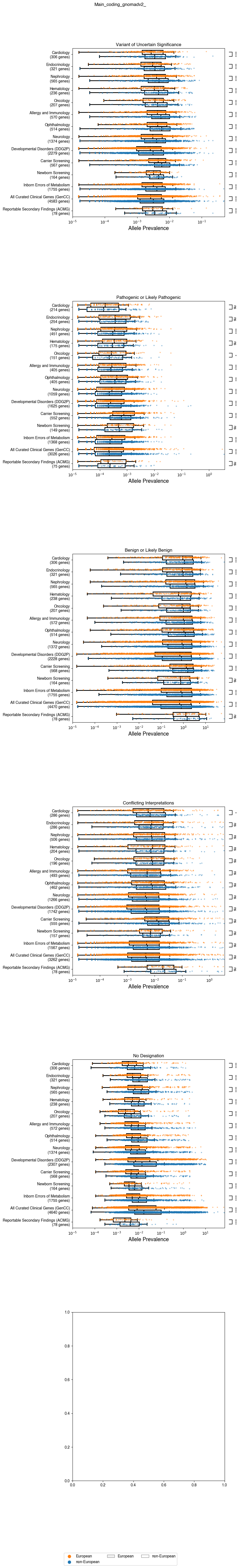

**Supplemental Figure 4c: Allele Prevalence Comparison of B/LB Variants in gnomAD v3.1.2 (non v2)**

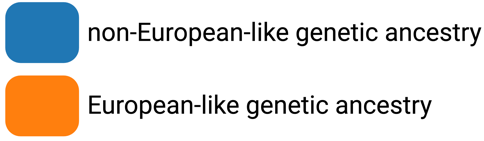

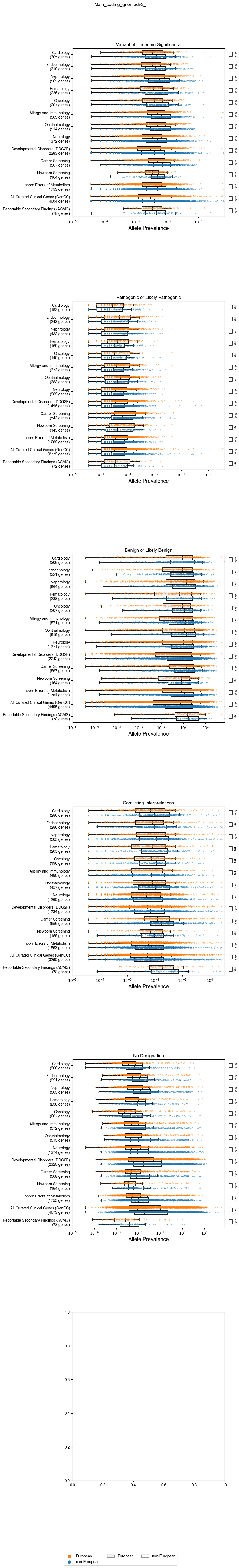

### Supplemental Figure 5: Allele Prevalence of Comparison of No Designation (ND) Variants

Box plots corresponding to ND allele prevalence (x-axis) in genes (dots) for individuals of non-European-like (blue) versus European-like (orange) genetic ancestry for the corresponding medical specialty (y-axis) in a) *All of Us* v7, b) gnomAD v2.1.1, and c) gnomAD v3.1.2 (non v2) for all coding variants. Genes with zero alleles for allele prevalence for either individuals of European-like or non-European-like genetic ancestry are omitted from the above visualization to maintain a reasonable scale for data visualization. However, genes with zero alleles for only one category of either individuals of European-like or non-European-like genetic ancestry are included in the Bonferroni-corrected, signed rank, matched pairs Wilcoxon statistical test. The Bonferroni corrected p-values associated with these comparisons are annotated as follows with "ns" indicating not significant, * for 1.19e-04 < p ≤ 2.38e-04, ** for 5.95e-05 < p ≤ 1.19e-04, *** for 5.95e-06 < p ≤ 5.95e-05, and **** for p ≤ 5.95e-06. Also refer to Tables S2-4. Overall, in the set of curated clinical genes as a whole (GenCC), we observe a significant increase in ND variants in individuals of non-European-like genetic ancestry compared to individuals of European-like genetic ancestry. However, this trend varies across specialties and gene lists.

**Supplemental Figure 5a: Allele Prevalence Comparison of ND Variants in *All of Us* v7**

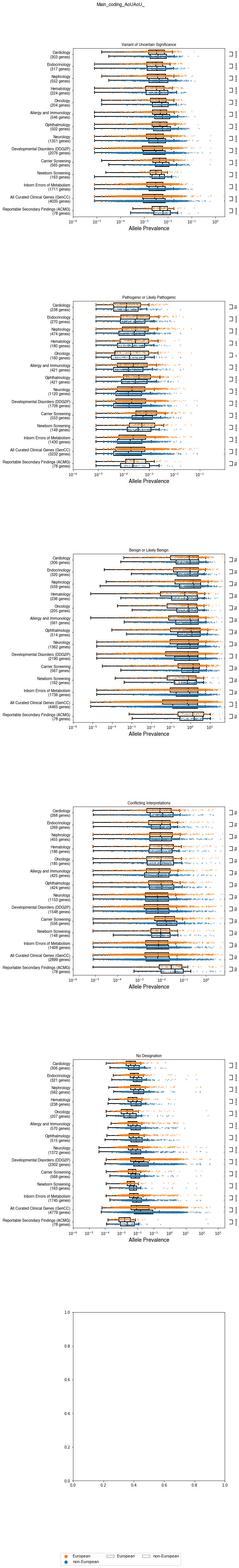

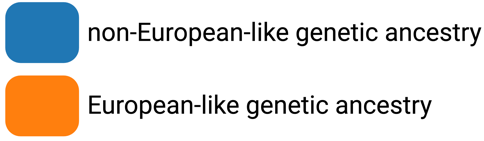

**Supplemental Figure 5b: Allele Prevalence Comparison of ND Variants in gnomAD v2.1.1**

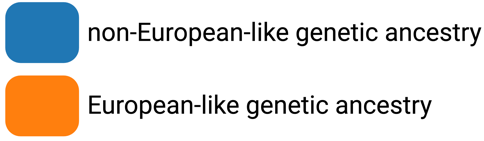

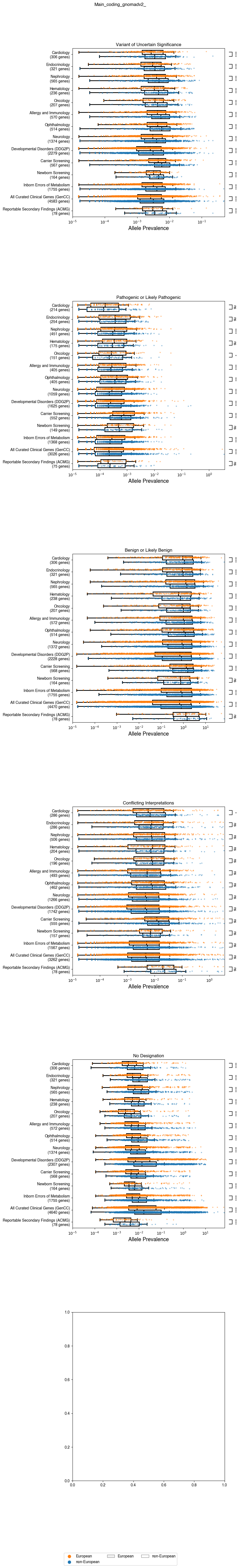

**Supplemental Figure 5c: Allele Prevalence Comparison of ND Variants in gnomAD v3.1.2 (non v2)**

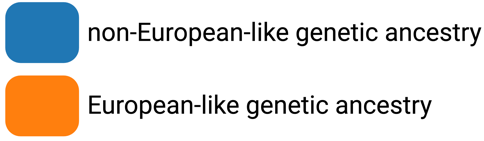

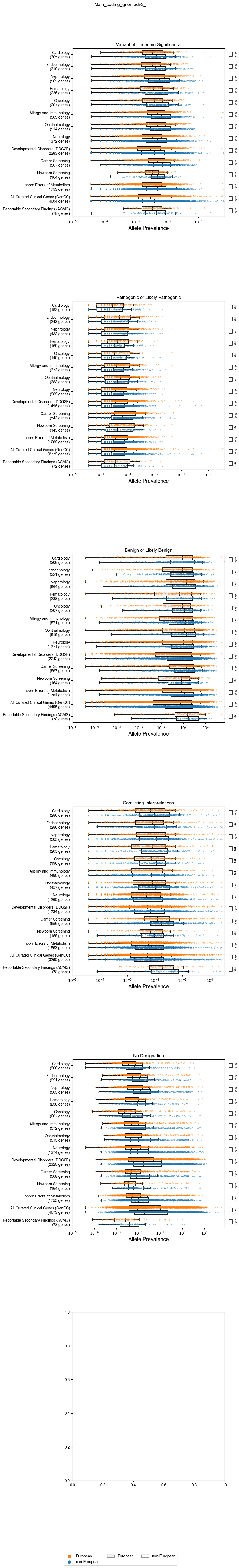

### Supplemental Figure 6: Allele Prevalence Comparison of Conflicting Interpretation (CI) Variants

Box plots corresponding to CI allele prevalence (x-axis) in genes (dots) for individuals of non-European-like (blue) versus European-like (orange) genetic ancestry for the corresponding medical specialty (y-axis) a) *All of Us* v7, b) gnomAD v2.1.1, and c) gnomAD v3.1.2 (non v2) for all coding variants. Genes with zero alleles for allele prevalence for either individuals of European-like or non-European-like genetic ancestry are omitted from the above visualization to maintain a reasonable scale for data visualization. However, genes with zero alleles for only one category of either individuals of European-like or non-European-like genetic ancestry are included in the Bonferroni-corrected, signed rank, matched pairs Wilcoxon statistical test. The Bonferroni corrected p-values associated with these comparisons are annotated as follows with "ns" indicating not significant, * for 1.19e-04 < p ≤ 2.38e-04, ** for 5.95e-05 < p ≤ 1.19e-04, *** for 5.95e-06 < p ≤ 5.95e-05, and **** for p ≤ 5.95e-06. Also refer to Tables S2-4. Overall, in the set of curated clinical genes as a whole (GenCC), we observe a significant increase in CI variants in individuals of non-European-like genetic ancestry compared to individuals of European-like genetic ancestry. However, this trend varies across specialties and gene lists.

**Supplemental Figure 6a: Allele Prevalence Comparison of CI Variants in *All of Us* v7**

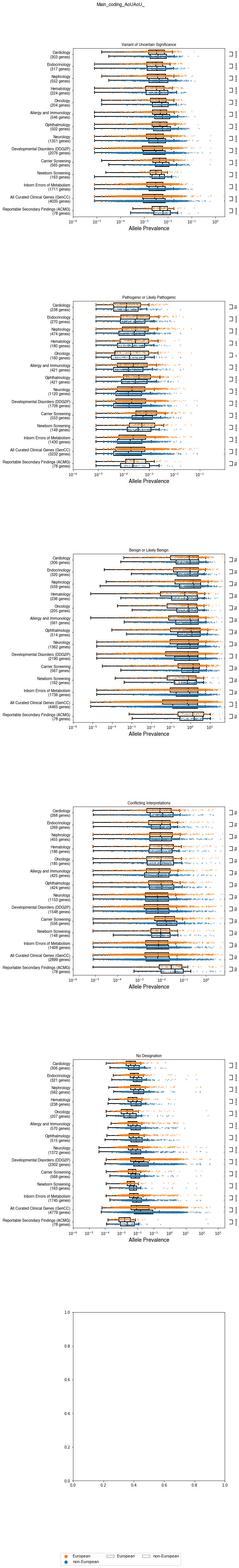

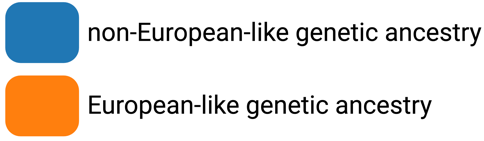

**Supplemental Figure 6b: Allele Prevalence Comparison of CI Variants in gnomAD v2.1.1**

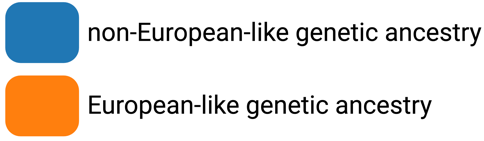

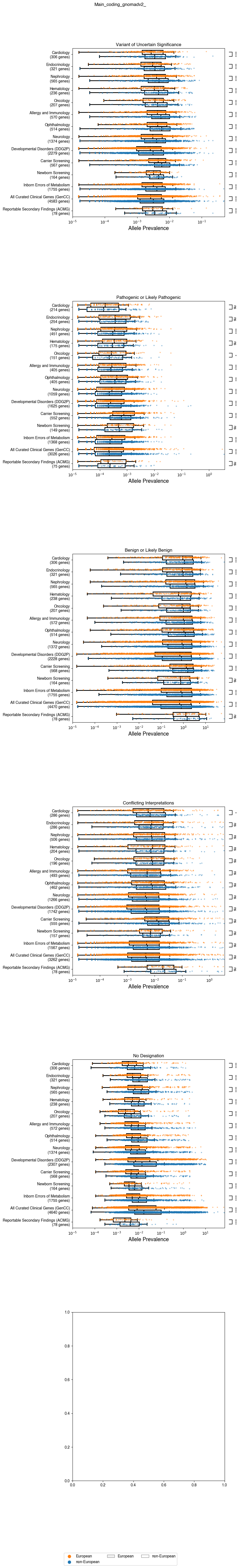

**Supplemental Figure 6c: Allele Prevalence Comparison of CI Variants in gnomAD v3.1.2 (non v2)**

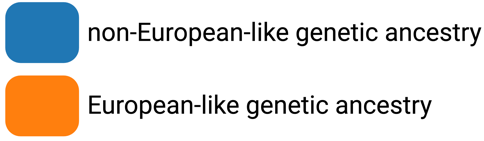

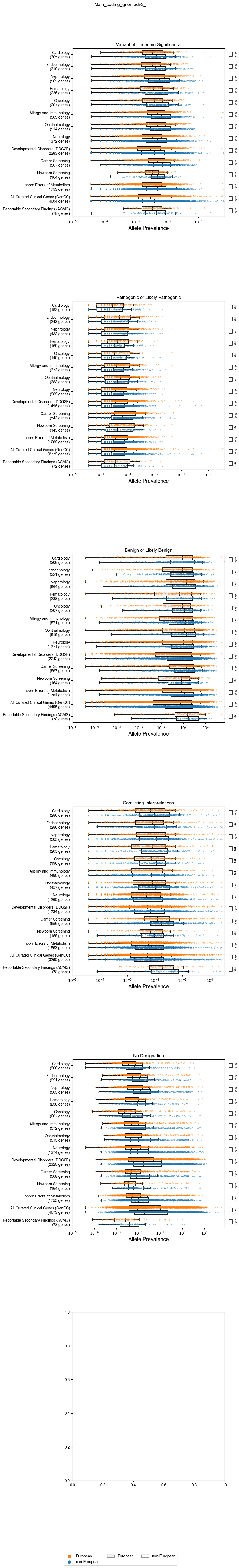

### Supplemental Figure 7: Allele Prevalence Comparison of Noncoding Variants of Uncertain Significance (VUS)

Box plots corresponding to VUS allele prevalence (x-axis) in genes (dot) for individuals of non-European-like (blue) versus European-like (orange) genetic ancestry for the corresponding medical specialty (y-axis) across a) gnomAD v2.1.1 and b) gnomAD v3.1.2 (non v2) for all noncoding variants. Genes with zero alleles for allele prevalence for either individuals of European-like or non-European-like genetic ancestry are omitted from the above visualization to maintain a reasonable scale for data visualization. However, genes with zero alleles for only one category of either individuals of European-like or non-European-like genetic ancestry are included in the Bonferroni-corrected, signed rank, matched pairs Wilcoxon statistical test. The Bonferroni corrected p-values associated with these comparisons are annotated according to the provided legend, with "ns" indicating not significant, * for 1.79e-04 < p ≤ 3.57e-04, ** for 8.93e-05 < p ≤ 1.79e-04, *** for 8.93e-06 < p ≤ 8.93e-05, and **** for p ≤ 8.93e-06. Also refer to Tables S5-6. Across all medical specialties and categories shown, VUS are not observed to be statistically significantly increased in individuals of non-European-like genetic ancestry compared to individuals of European-like genetic ancestry.

**Supplemental Figure 7a: Allele Prevalence Comparison of Noncoding VUS in gnomAD v2.1.1**

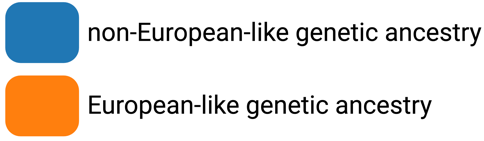

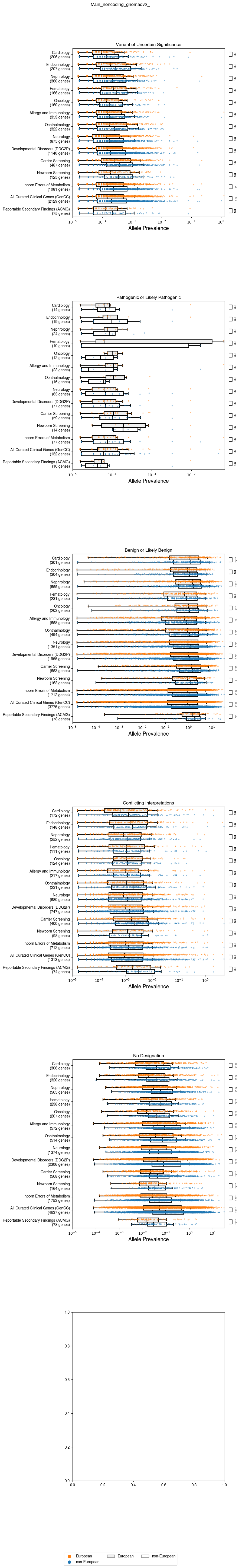

**Supplemental Figure 7b: Allele Prevalence Comparison of Noncoding VUS in gnomAD v3.1.2 (non v2)**

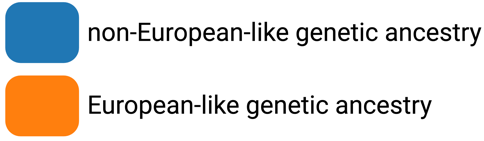

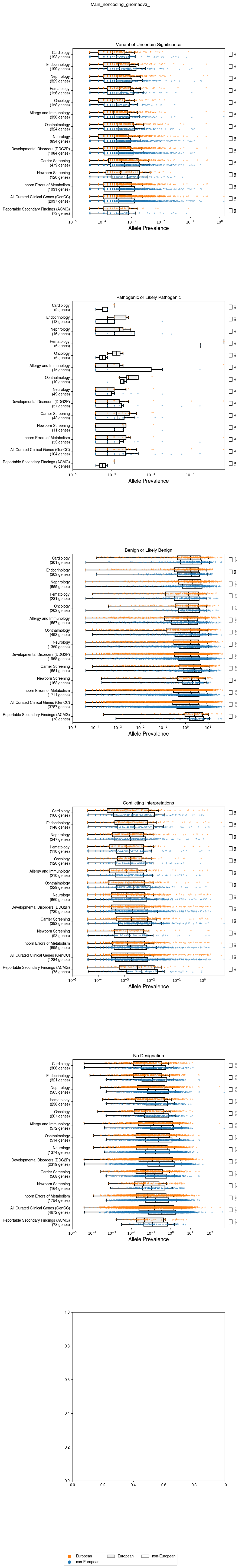

### Supplemental Figure 8: Allele Prevalence Comparison of Noncoding Pathogenic or Likely Pathogenic (P/LP) Variants

Box plots corresponding to P/LP allele prevalence (x-axis) in genes (dot) for individuals of non-European-like (blue) versus European-like (orange) genetic ancestry for the corresponding medical specialty (y-axis) across a) gnomAD v2.1.1 and b) gnomAD v3.1.2 (non v2) for all noncoding variants. Genes with zero alleles for allele prevalence for either individuals of European-like or non-European-like genetic ancestry are omitted from the above visualization to maintain a reasonable scale for data visualization. However, genes with zero alleles for only one category of either individuals of European-like or non-European-like genetic ancestry are included in the Bonferroni-corrected, signed rank, matched pairs Wilcoxon statistical test. The Bonferroni corrected p-values associated with these comparisons are annotated according to the provided legend, with "ns" indicating not significant, * for 1.79e-04 < p ≤ 3.57e-04, ** for 8.93e-05 < p ≤ 1.79e-04, *** for 8.93e-06 < p ≤ 8.93e-05, and **** for p ≤ 8.93e-06. Also refer to Tables S5-6. Across all medical specialties and categories shown, P/LP variants are not observed to be statistically significantly increased in individuals of non-European-like genetic ancestry compared to individuals of European-like genetic ancestry.

**Supplemental Figure 8a: Allele Prevalence Comparison of Noncoding P/LP variants in gnomAD v2.1.1**

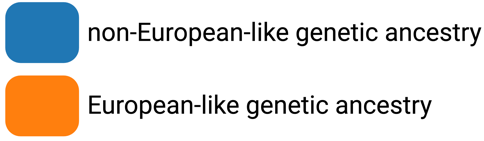

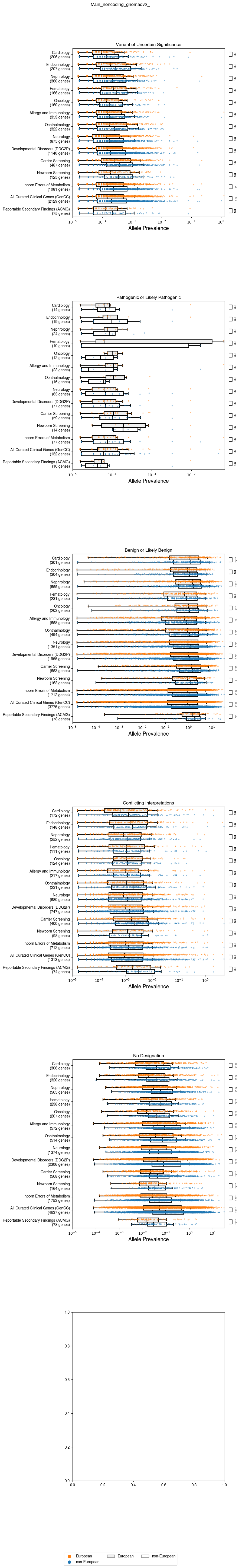

**Supplemental Figure 8b: Allele Prevalence Comparison of Noncoding P/LP variants in gnomAD v3.1.2 (non v2)**

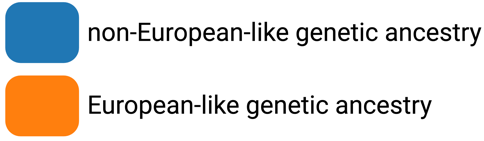

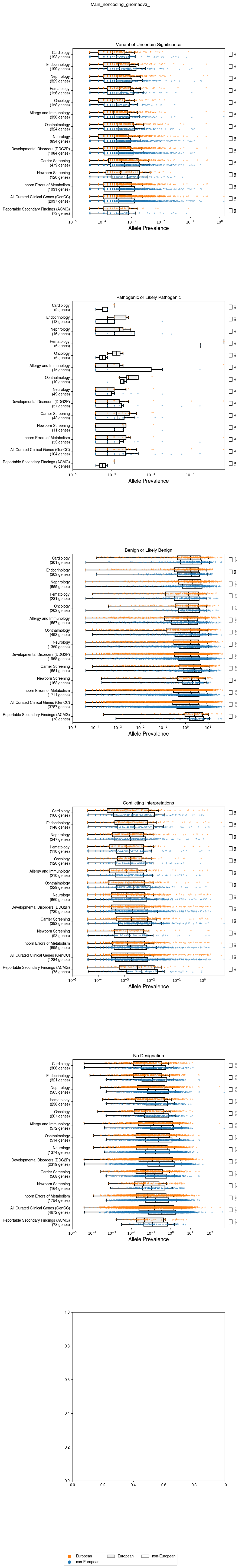

### Supplemental Figure 9: Allele Prevalence Comparison of Noncoding Benign or Likely Benign (B/LB) Variants

Box plots corresponding to B/LB allele prevalence (x-axis) in genes (dot) for individuals of non-European-like (blue) versus European-like (orange) genetic ancestry for the corresponding medical specialty (y-axis) across a) gnomAD v2.1.1 and b) gnomAD v3.1.2 (non v2) for all noncoding variants. Genes with zero alleles for allele prevalence for either individuals of European-like or non-European-like genetic ancestry are omitted from the above visualization to maintain a reasonable scale for data visualization. However, genes with zero alleles for only one category of either individuals of European-like or non-European-like genetic ancestry are included in the Bonferroni-corrected, signed rank, matched pairs Wilcoxon statistical test. The Bonferroni corrected p-values associated with these comparisons are annotated according to the provided legend, with "ns" indicating not significant, * for 1.79e-04 < p ≤ 3.57e-04, ** for 8.93e-05 < p ≤ 1.79e-04, *** for 8.93e-06 < p ≤ 8.93e-05, and **** for p ≤ 8.93e-06. Also refer to Tables S5-6. Across all medical specialties and categories shown, the prevalence of noncoding B/LB variants are observed to be statistically significantly increased in individuals of non-European-like genetic ancestry compared to individuals of European-like genetic ancestry across almost all the medical specialties assessed.

**Supplemental Figure 9a: Allele Prevalence Comparison of Noncoding B/LB gnomAD v2.1.1**

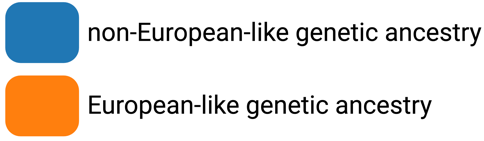

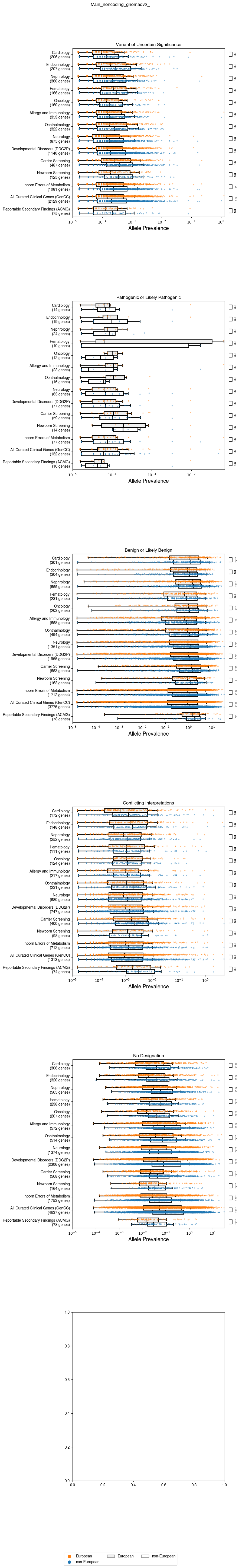

**Supplemental Figure 9b: Allele Prevalence Comparison of Noncoding B/LB gnomAD v3.1.2 (non v2)**

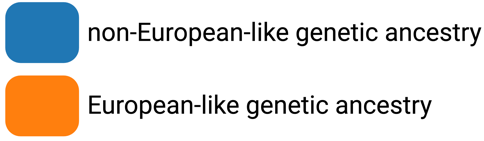

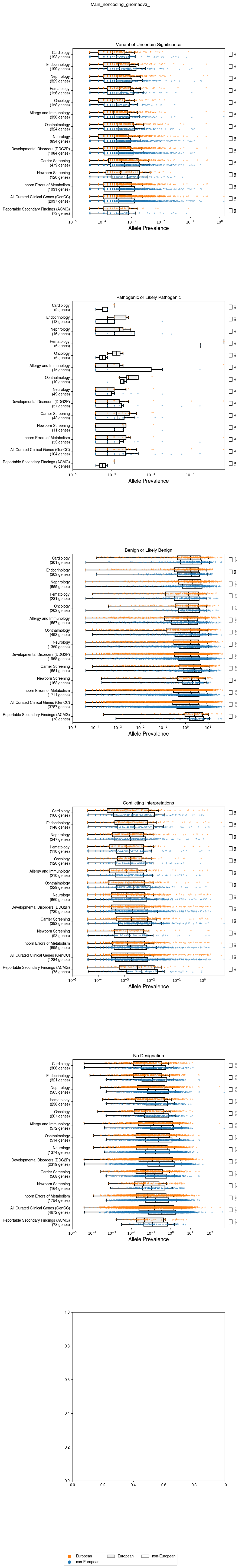

### Supplemental Figure 10: Allele Prevalence Comparison of Noncoding Conflicting Interpretation (CI) Variants

Box plots corresponding to CI allele prevalence (x-axis) in genes (dot) for individuals of non-European-like (blue) versus European-like (orange) genetic ancestry for the corresponding medical specialty (y-axis) across a) gnomAD v2.1.1 and b) gnomAD v3.1.2 (non v2) for all noncoding variants. Genes with zero alleles for allele prevalence for either individuals of European-like or non-European-like genetic ancestry are omitted from the above visualization to maintain a reasonable scale for data visualization. However, genes with zero alleles for only one category of either individuals of European-like or non-European-like genetic ancestry are included in the Bonferroni-corrected, signed rank, matched pairs Wilcoxon statistical test. The Bonferroni corrected p-values associated with these comparisons are annotated according to the provided legend, with "ns" indicating not significant, * for 1.79e-04 < p ≤ 3.57e-04, ** for 8.93e-05 < p ≤ 1.79e-04, *** for 8.93e-06 < p ≤ 8.93e-05, and **** for p ≤ 8.93e-06. Also refer to Tables S5-6. Across all medical specialties and categories shown, the prevalence of noncoding CI variants are not observed to be statistically significantly increased in individuals of non-European-like genetic ancestry compared to individuals of European-like genetic ancestry.

**Supplemental Figure 10a: Allele Prevalence Comparison of Noncoding CI variants in gnomAD v2.1.1**

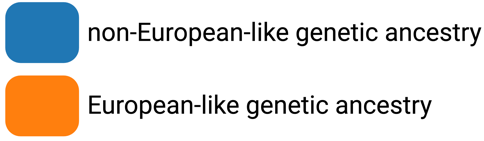

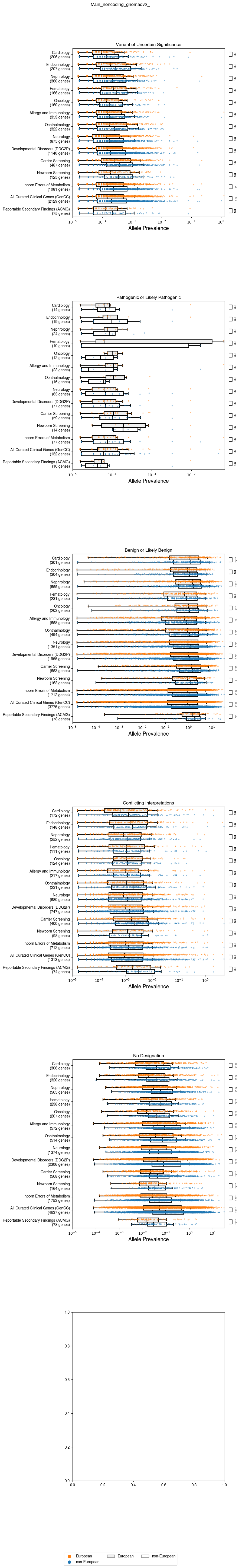

**Supplemental Figure 10b: Allele Prevalence Comparison of Noncoding CI variants in gnomAD v3.1.2 (non v2)**

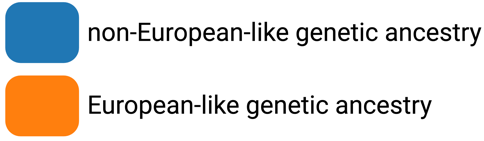

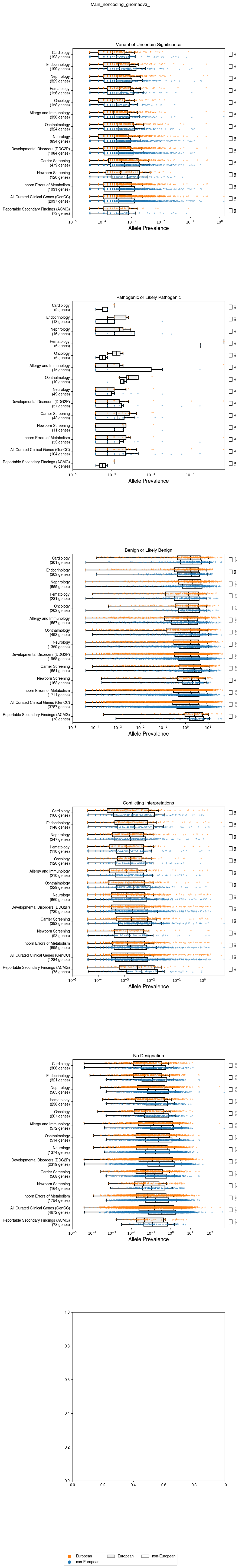

### Supplemental Figure 11: Allele Prevalence Comparison of Noncoding No Designation (ND) Variants

Box plots corresponding to ND allele prevalence (x-axis) in each gene (dot) for individuals of non-European-like (blue) versus European-like (orange) genetic ancestry for the corresponding medical specialty (y-axis) across a) gnomAD v2.1.1 and b) gnomAD v3.1.2 (non v2) for all noncoding variants. Genes with zero alleles for allele prevalence for either individuals of European-like or non-European-like genetic ancestry are omitted from the above visualization to maintain a reasonable scale for data visualization. However, genes with zero alleles for only one category of either individuals of European-like or non-European-like genetic ancestry are included in the Bonferroni-corrected, signed rank, matched pairs Wilcoxon statistical test. The Bonferroni corrected p-values associated with these comparisons are annotated according to the provided legend, with "ns" indicating not significant, * for 1.79e-04 < p ≤ 3.57e-04, ** for 8.93e-05 < p ≤ 1.79e-04, *** for 8.93e-06 < p ≤ 8.93e-05, and **** for p ≤ 8.93e-06. Also refer to Tables S5-6. Across all medical specialties and categories shown, ND variants are observed to be statistically significantly increased in individuals of non-European-like genetic ancestry compared to individuals of European-like genetic ancestry.

**Supplemental Figure 11a: Allele Prevalence Comparison of Noncoding ND variants in gnomAD v2.1.1**

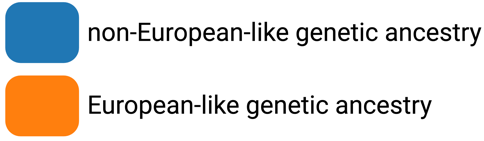

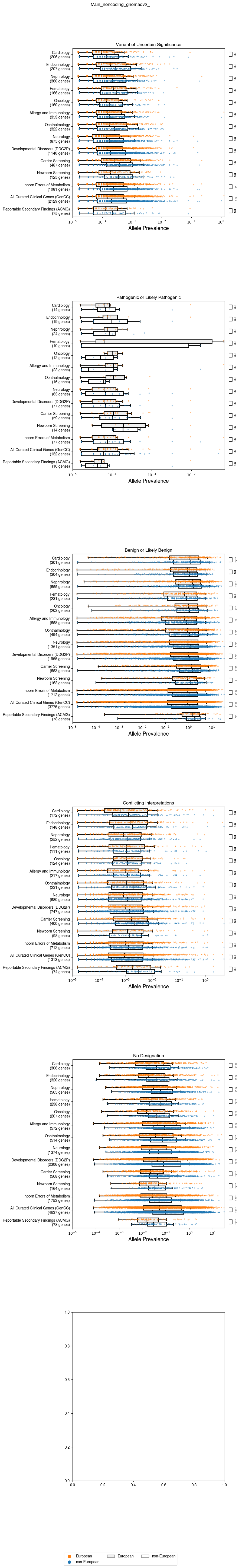

**Supplemental Figure 11b: Allele Prevalence Comparison of Noncoding ND variants in gnomAD v3.1.2 (non v2)**

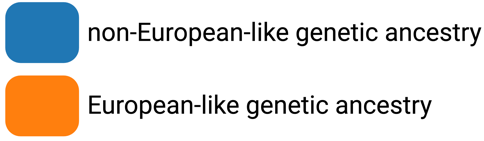

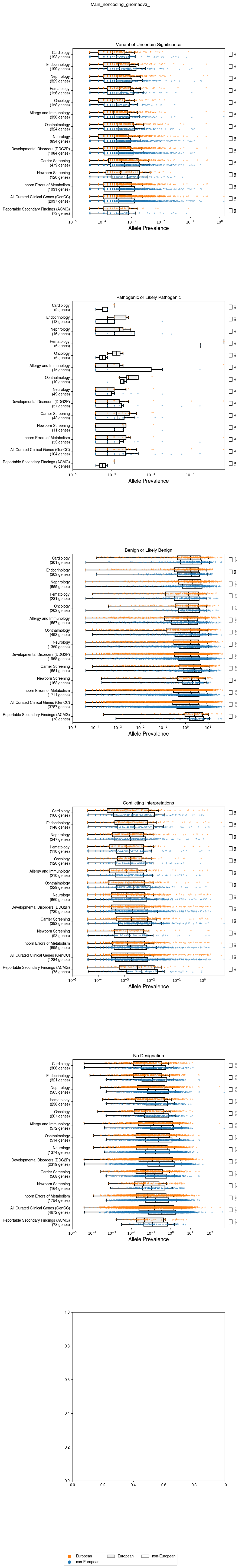

### Supplemental Figure 12: Top 20 Genes With Greatest VUS Disparity for the Reportable Secondary Findings Gene List and All Curated Clinical Genes

Bar charts plotting difference in VUS Disparity based on difference in allele prevalence (top row) or absolute difference in unique variant counts (bottom row) for a) the list of curated clinical genes (GenCC), b) the genes on the list for reportable secondary findings (ACMG), c) the genes that are high priority for saturation genome editing and d) the genes that are high priority for VAMP-seq for all three databases *All of Us* v7 (left column), gnomAD v2.1.1 (middle column), and gnomAD v3.1.2 (non v2) (right column). Many genes in the top 20 genes are in agreement across all three databases. Further, several of top ranked genes in the GenCC or ACMG lists are also near the top for the SGE and VAMP-seq lists meaning conducting the particular MAVE technique on that gene could potentially help close the VUS disparity for that gene.

**Supplemental Figure 12a: Top 20 Genes from the Reportable Secondary Findings Gene List (ACMG) with Greatest VUS Disparity Between Individuals of European-like Versus Non-European-like Genetic Ancestry**

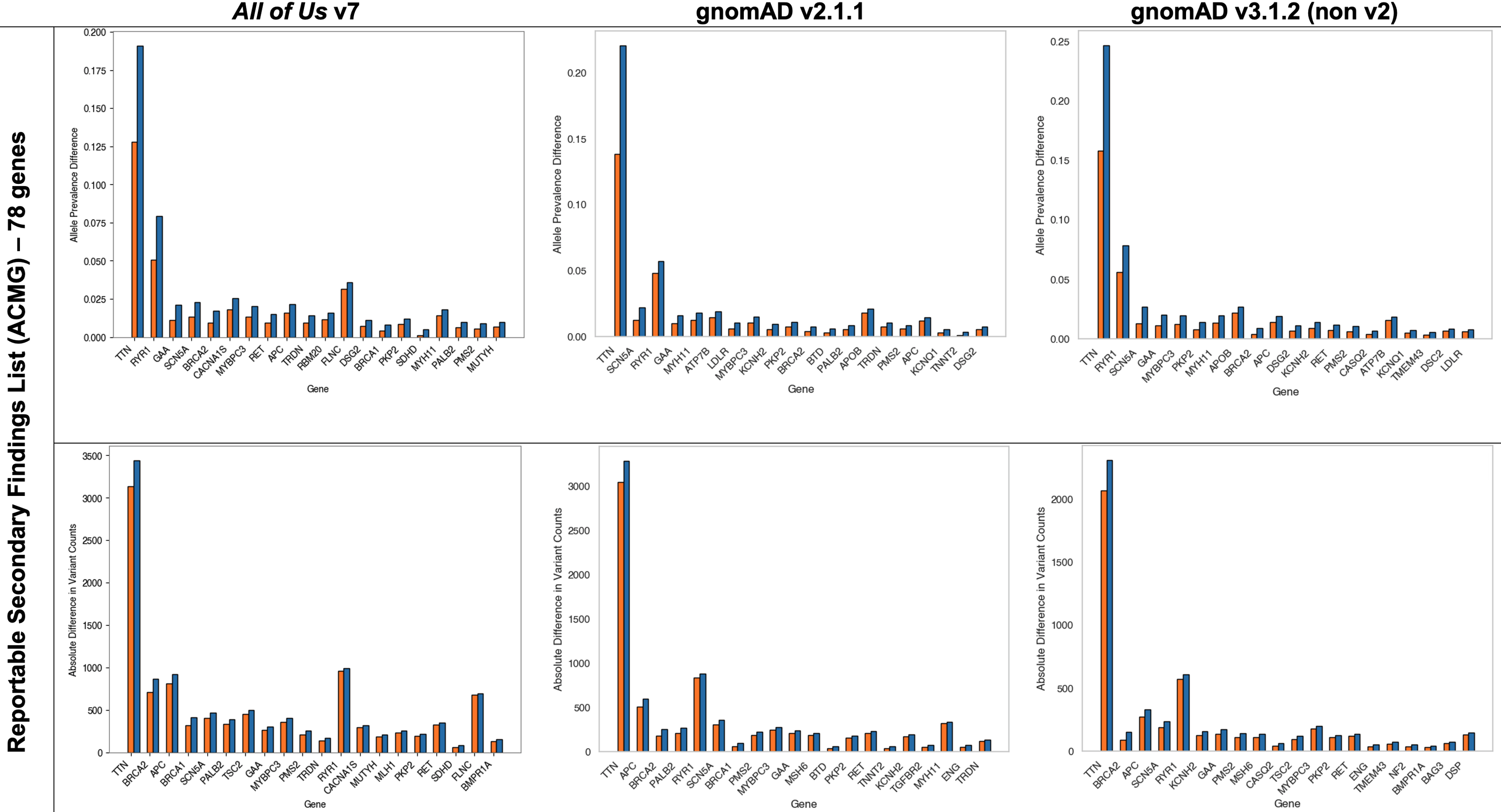

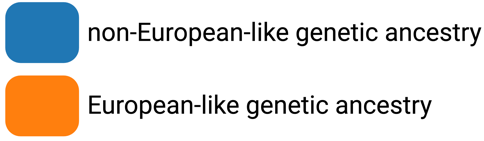

**Supplemental Figure 12b: Top 20 Genes from the List of All Curated Clinical Genes (GenCC) with Greatest VUS Disparity Between Individuals of European-like Versus Non-European-like Genetic Ancestry**

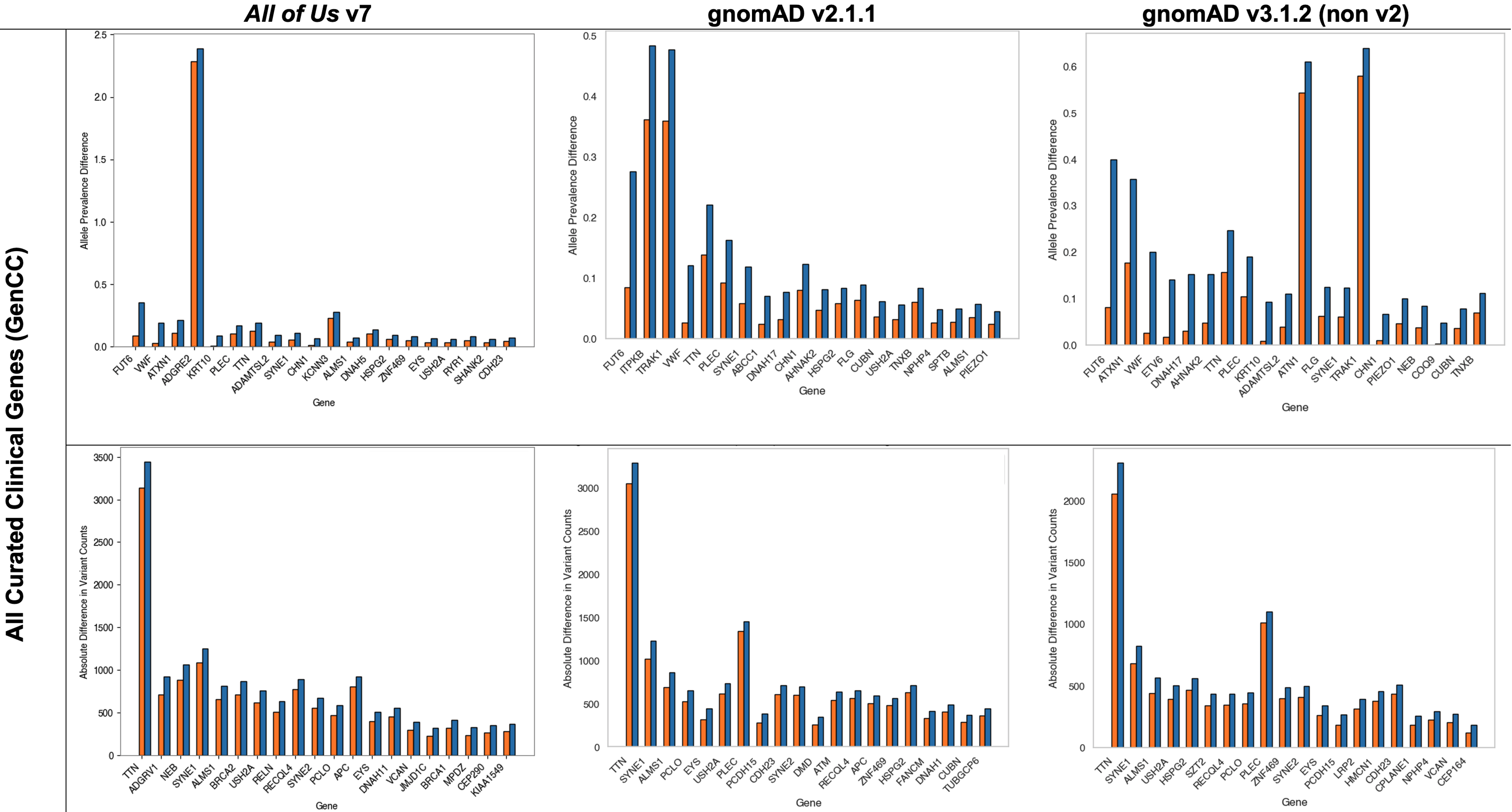

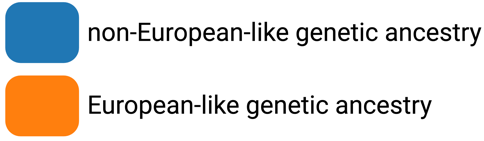

### Supplemental Figure 13: Top 20 Genes With Greatest VUS Disparity For Two Common MAVE Techniques

Bar charts plotting difference in VUS Disparity based on difference in allele prevalence (top row) or absolute difference in unique variant counts (bottom row) for a) the list of curated clinical genes (GenCC), b) the genes on the list for reportable secondary findings (ACMG), c) the genes that are high priority for saturation genome editing and d) the genes that are high priority for VAMP-seq for all three databases *All of Us* v7 (left column), gnomAD v2.1.1 (middle column), and gnomAD v3.1.2 (non v2) (right column). Many genes in the top 20 genes are in agreement across all three databases. Further, several of top ranked genes in the GenCC or ACMG lists are also near the top for the SGE and VAMP-seq lists meaning conducting the particular MAVE technique on that gene could potentially help close the VUS disparity for that gene.

**Supplemental Figure 13a: Top 20 Genes from the List of Clinical Genes (GenCC) That Are Also Targets of Saturation Genome Editing With Greatest VUS Disparity Between Individuals of European-like Versus Non-European-like Genetic Ancestry**

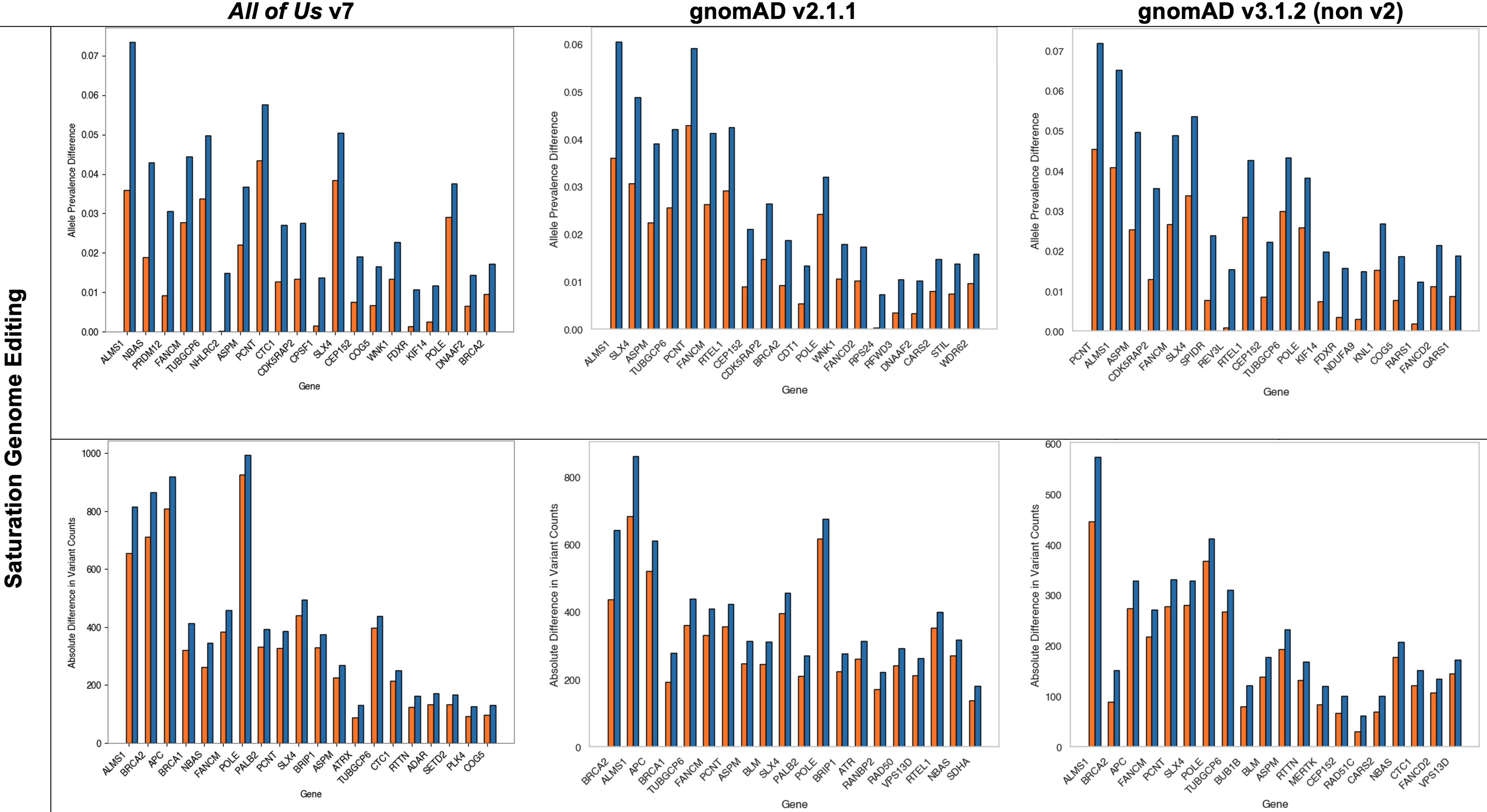

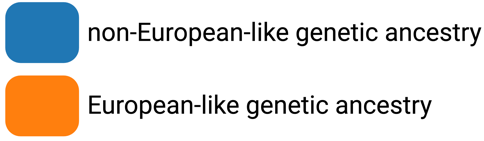

**Supplemental Figure 13b: Top 20 Genes from the List of Clinical Genes (GenCC) That Are Also Targets of VAMP-seq With Greatest VUS Disparity Between Individuals of European-like Versus Non-European-like Genetic Ancestry**

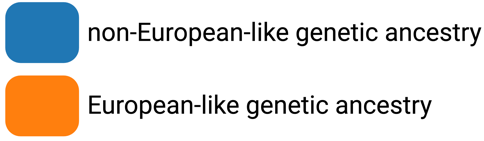

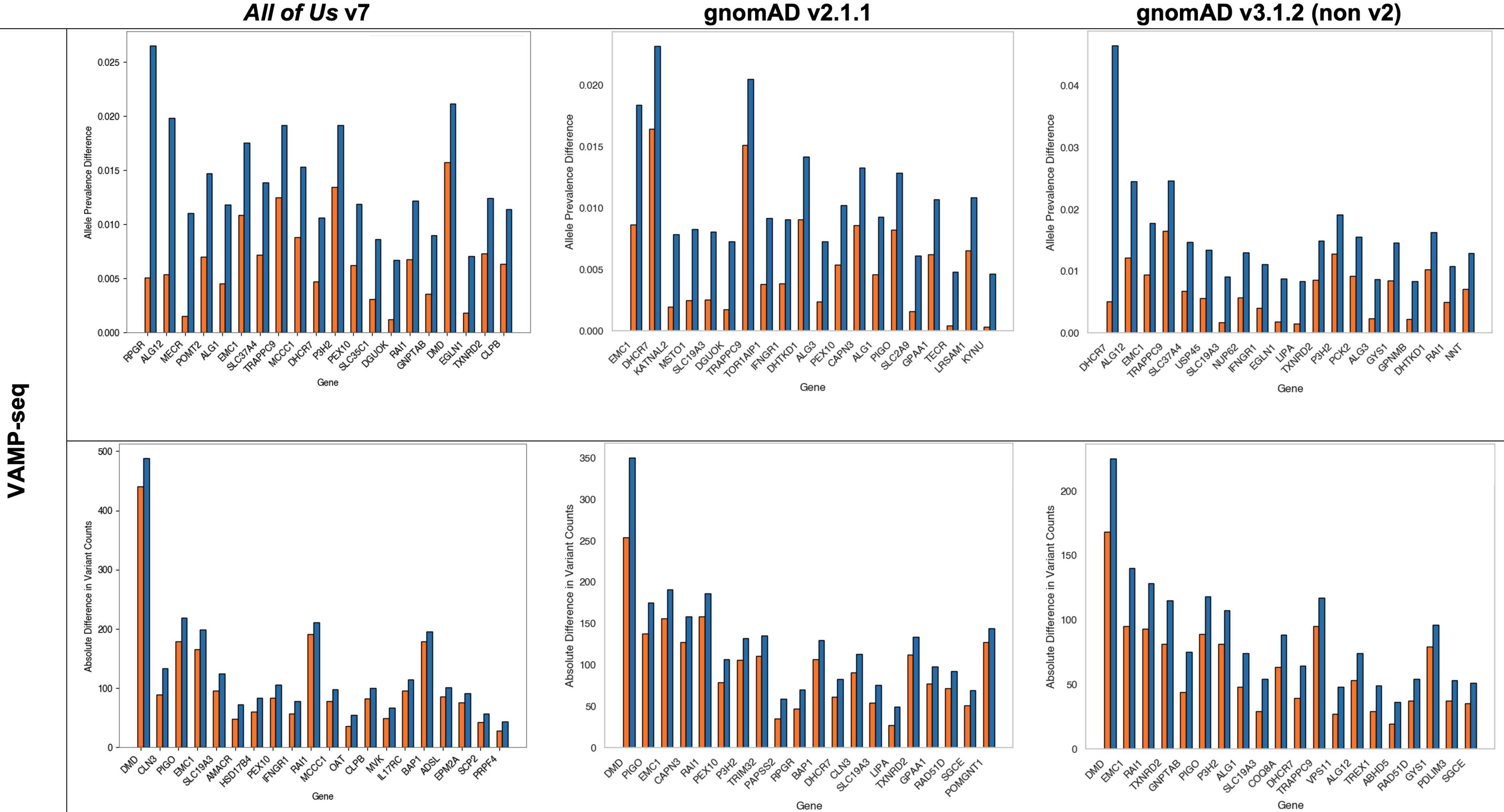

### Supplemental Figure 14: Comparative Analysis of VUS and P/LP variants in Genes for Potential MAVE Assays

Box plots corresponding to distribution of allele prevalence (x-axis) of a) VUS and b) P/LP variants for individuals of non-European-like (blue) versus European-like (orange) genetic ancestry in genes (dots) amenable to saturation genome editing (SGE), variant abundance by massively parallel sequencing (VAMP-seq), or currently being studied and registered into the MAVE Registry (y-axis) across *All of Us* v7 (top row), gnomAD v2.1.1 (middle row), and gnomAD v3.1.2 (non v2) (bottom row) for all coding variants. Genes with zero alleles for allele prevalence for either individuals of European-like or non-European-like genetic ancestry are omitted from the above visualization to maintain a reasonable scale for data visualization. However, genes with zero alleles for only one category of either individuals of European-like or non-European-like genetic ancestry are included in the Bonferroni-corrected, signed rank, matched pairs Wilcoxon statistical test. The Bonferroni corrected p-values associated with these comparisons are annotated as follows with "ns" indicating not significant, * for 1.19e-04 < p ≤ 2.38e-04, ** for 5.95e-05 < p ≤ 1.19e-04, *** for 5.95e-06 < p ≤ 5.95e-05, and **** for p ≤ 5.95e-06. Also refer to Tables S7-9. VUS are observed to be statistically significantly increased in Non-European-like populations compared to European-like populations for all three comparisons in all three databases, whereas P/LP variants are observed to be statistically significantly increased in individuals of European-like genetic ancestry compared to non-European-like for all three comparisons in all three databases. These trends align with the overall trends found while describing the disparities across medical specialties. Thus, for the genes in MAVERegistry, if MAVEs are fully realized for those genes in the coming years, those data will be able to potentially compensate for the VUS disparity in those ~100 genes. Further, if the SGE and VAMP-seq MAVEs are conducted for the 800+ examined genes, then the massive VUS reclassification from that data may help in closing the VUS disparity across medical specialties in all of clinical genetics.

**Supplemental Figure 14a: Higher Prevalence of VUS Found in Gene Lists for SGE, VAMP-seq, and current targets in the MAVE Registry in Individuals of Non-European-like Versus European-like Genetic Ancestry**

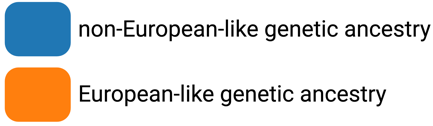

| *All of Us* v7 | 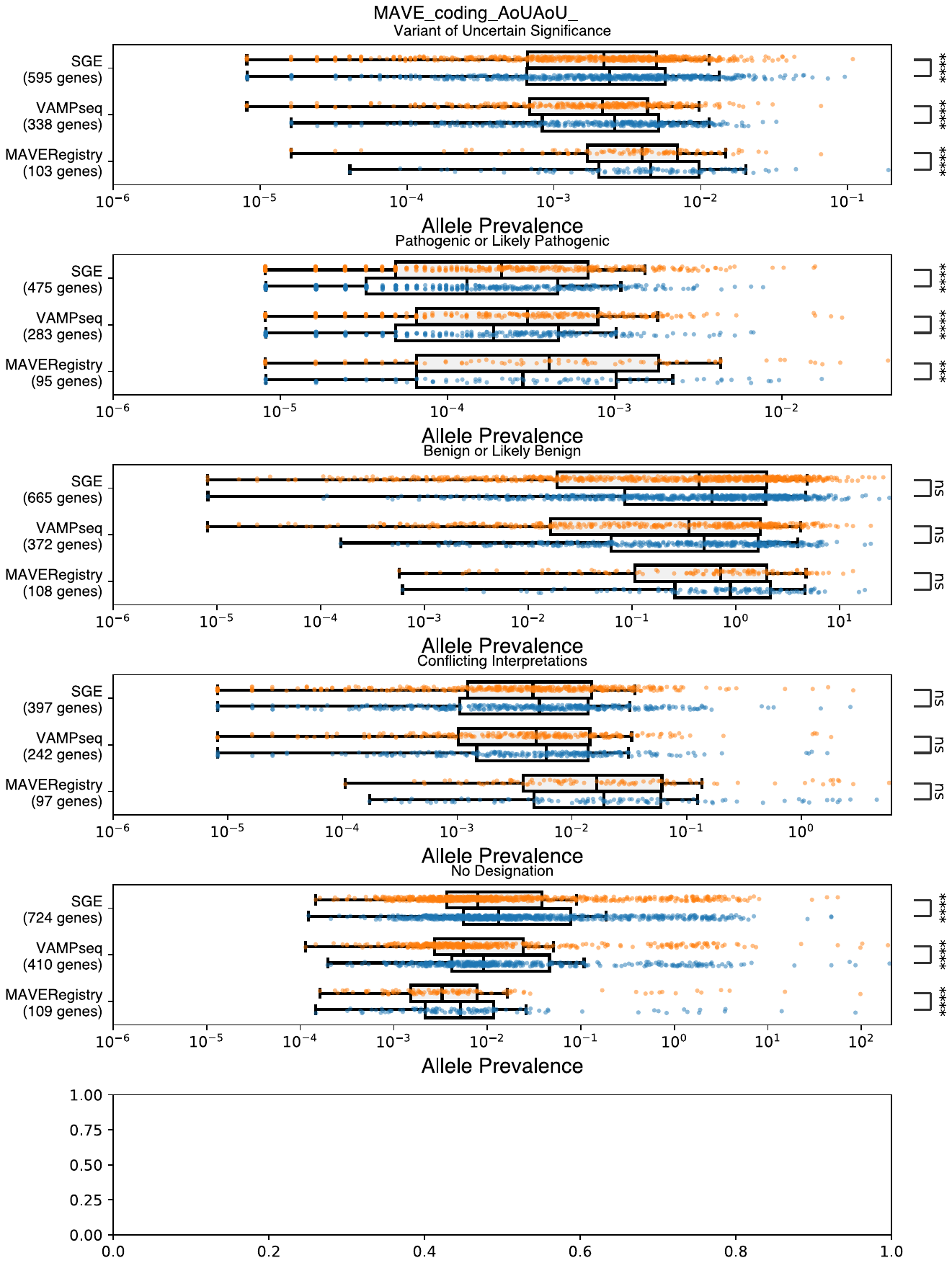 |
| --- | --- |
| gnomAD v2.1.1 | 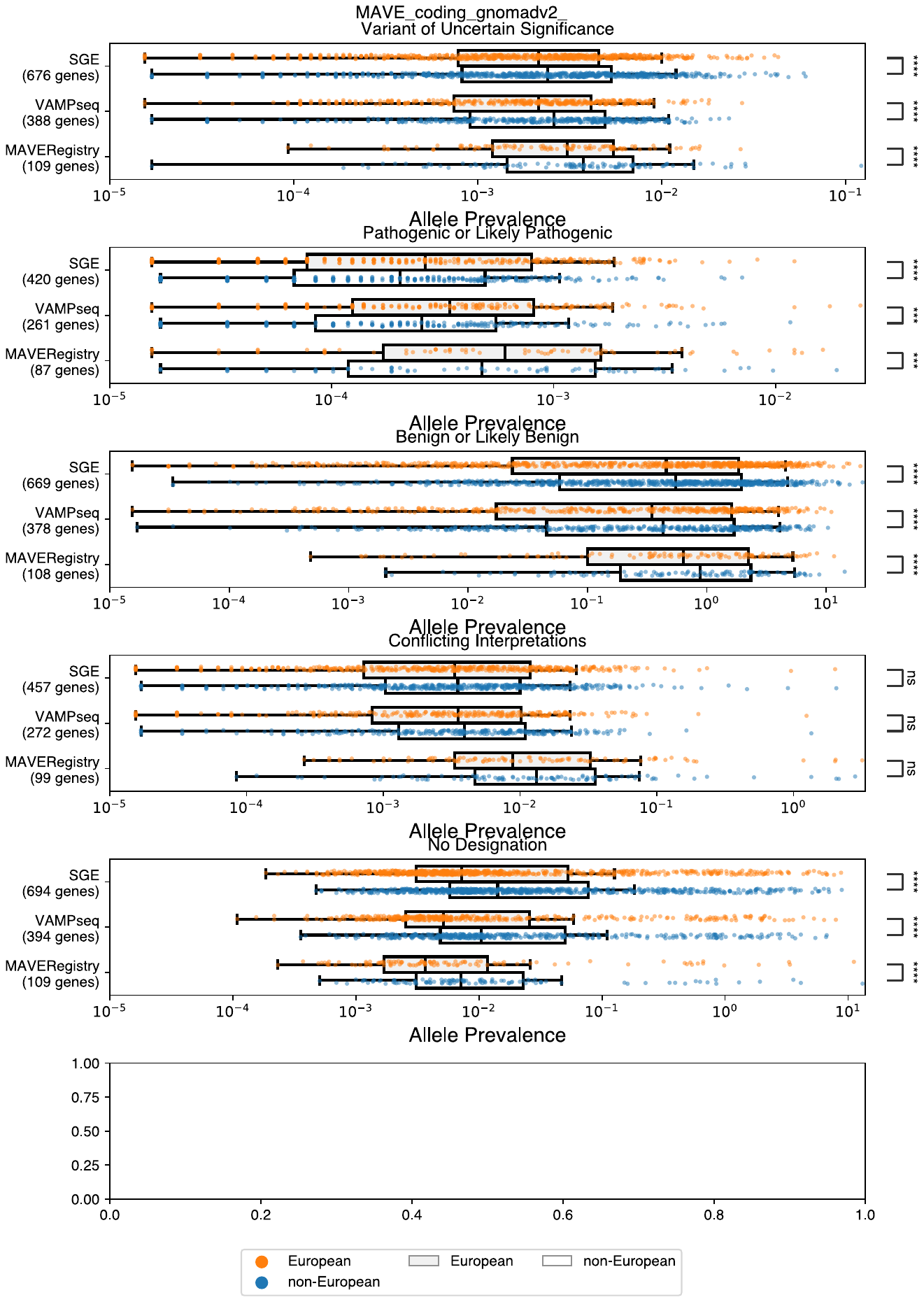 |
| gnomAD v3.1.2 (non v2) | 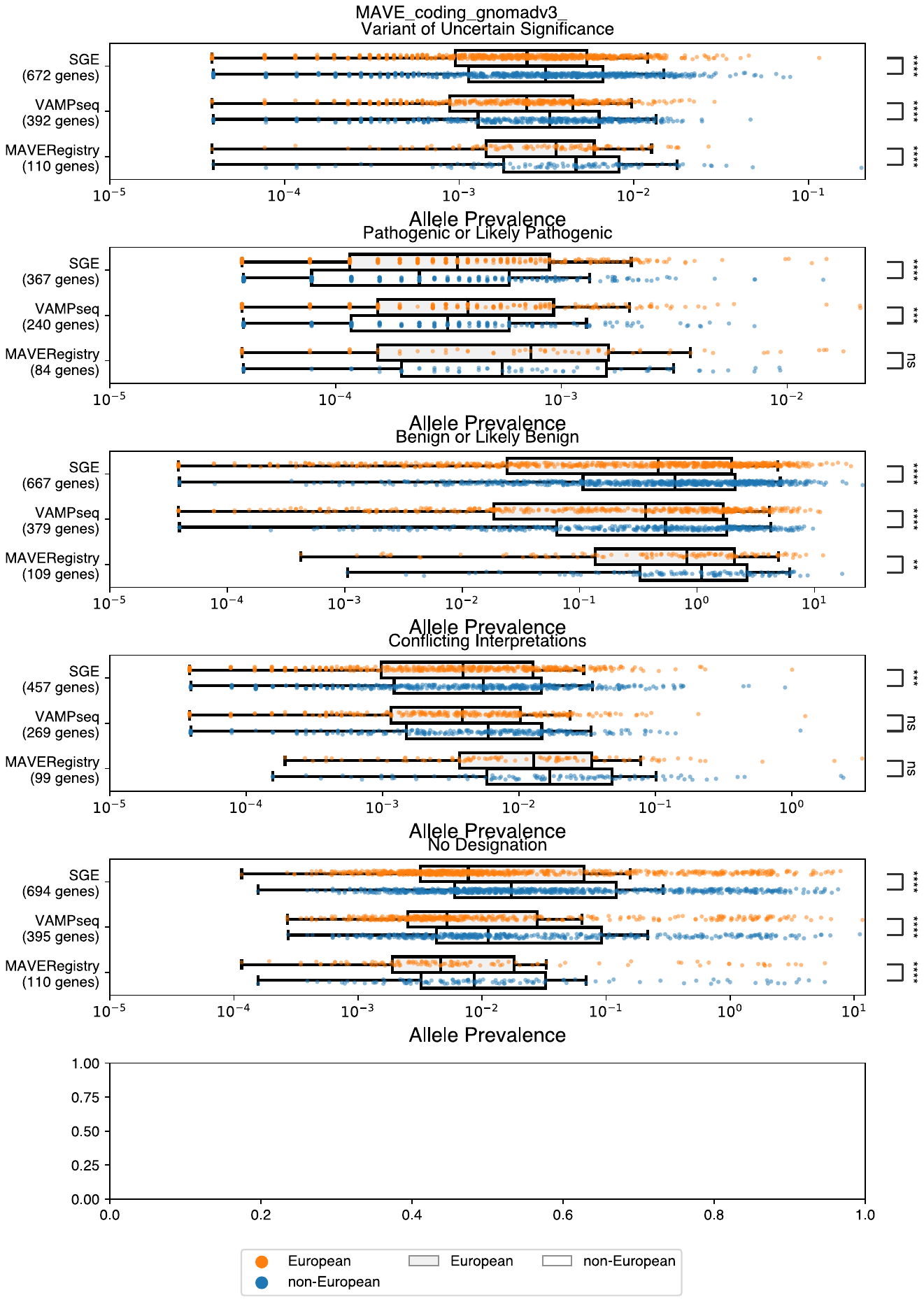 |

**Supplemental Figure 14b: Higher Prevalence of P/LP Variants Found in Gene Lists for SGE, VAMP-seq, and current targets in the MAVE Registry in Individuals of Non-European-like Versus European-like Genetic Ancestry**

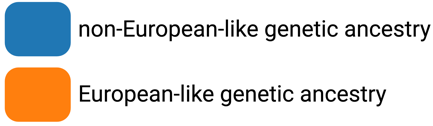

| *All of Us* v7 | 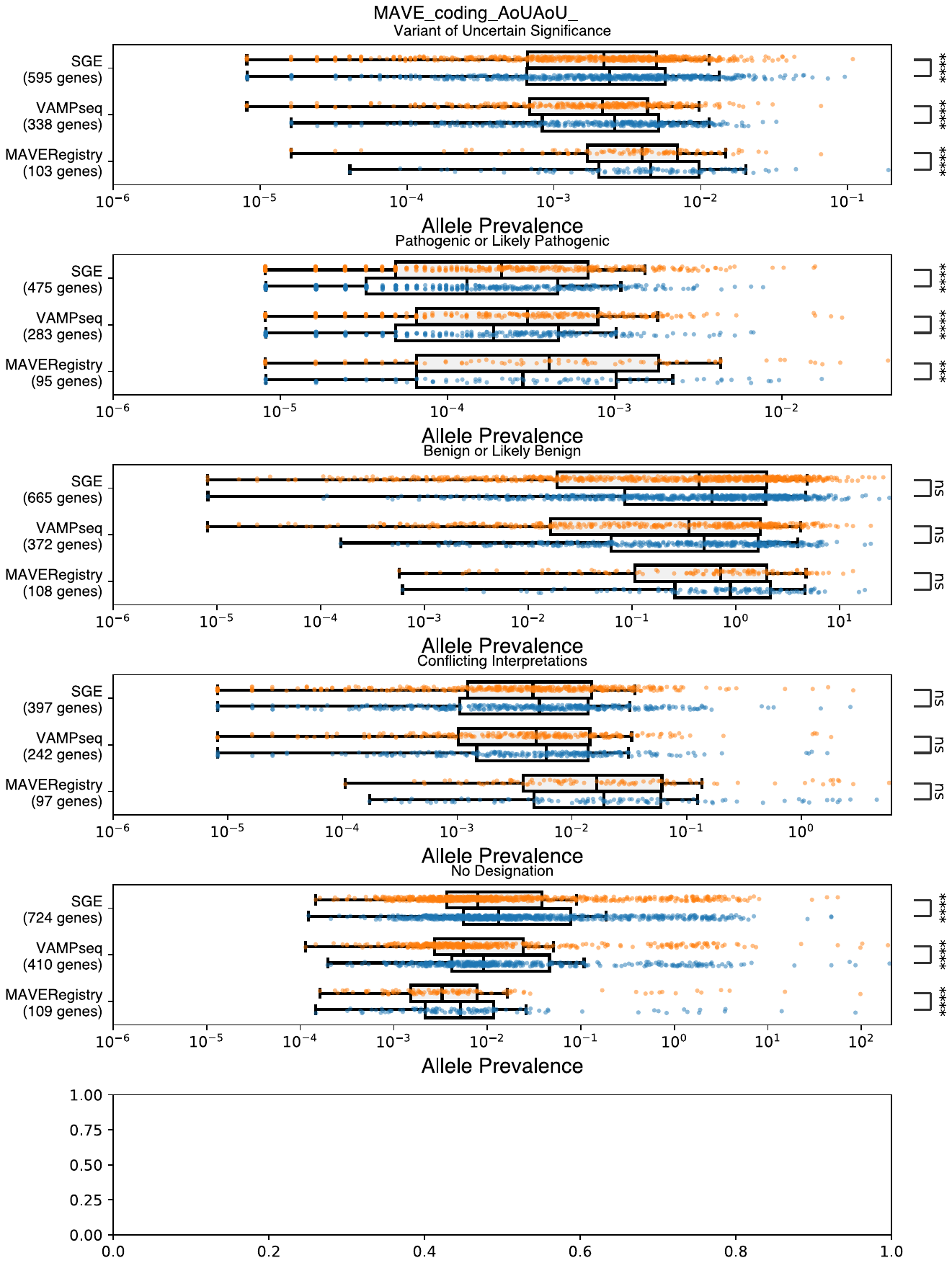 |
| --- | --- |
| gnomAD v2.1.1 | 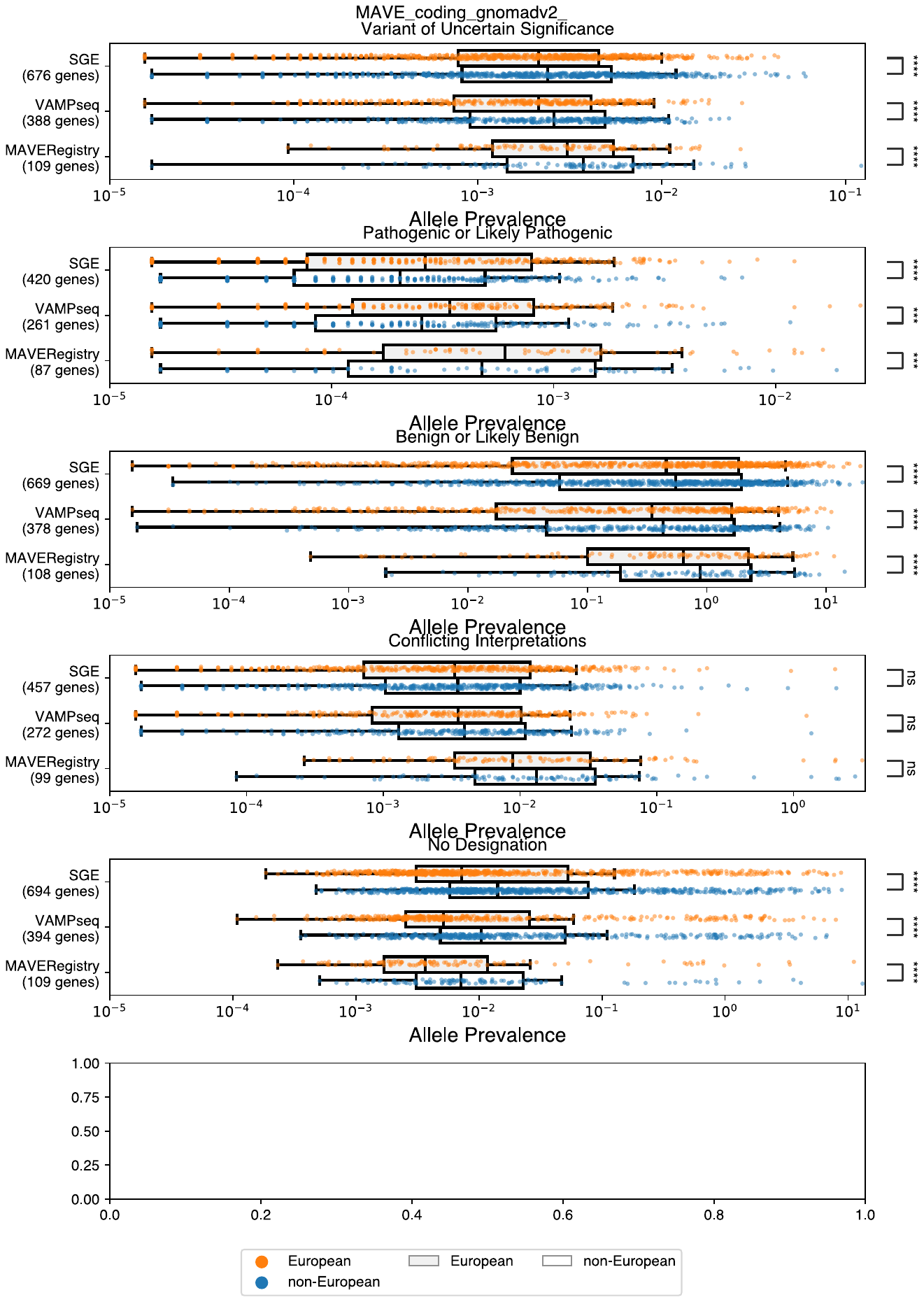 |
| gnomAD v3.1.2 (non v2) | 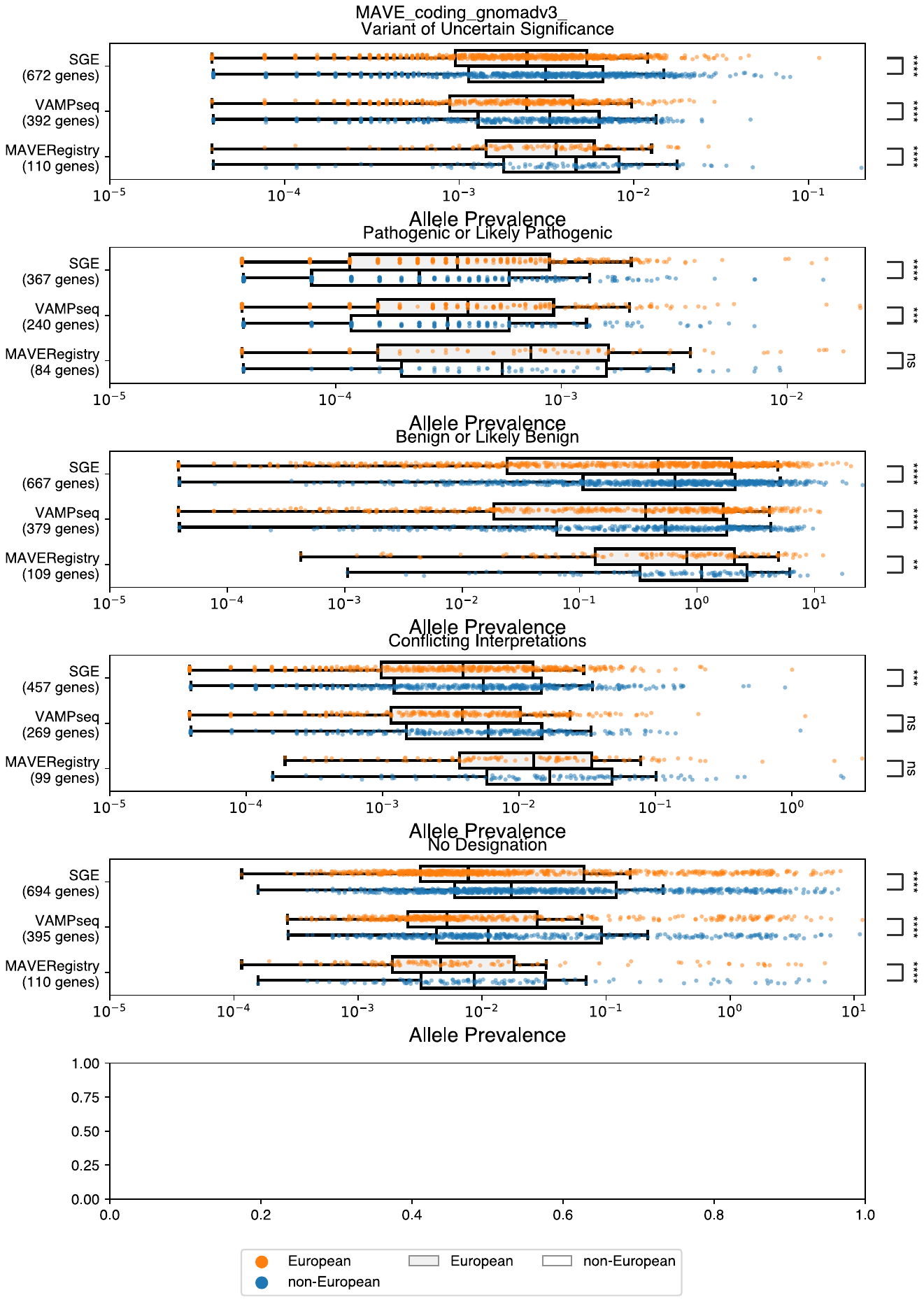 |

### Supplemental Figure 15: Variant Composition of Variants of Uncertain Significance (VUS) Across Medical Specialties

Pie charts representing the variant spectrum of VUS for all genes within the particular medical specialty in a) *All of Us* v7, b) gnomAD v2.1.1, c) gnomAD v3.1.2. The most prevalent VUS variant type, missense variants (light blue), accounts for at minimum 83% of VUS in any given specialty across all three database.

**Supplemental Figure 15a: Variant Composition of VUS Across Medical Specialties in *All of Us* v7**

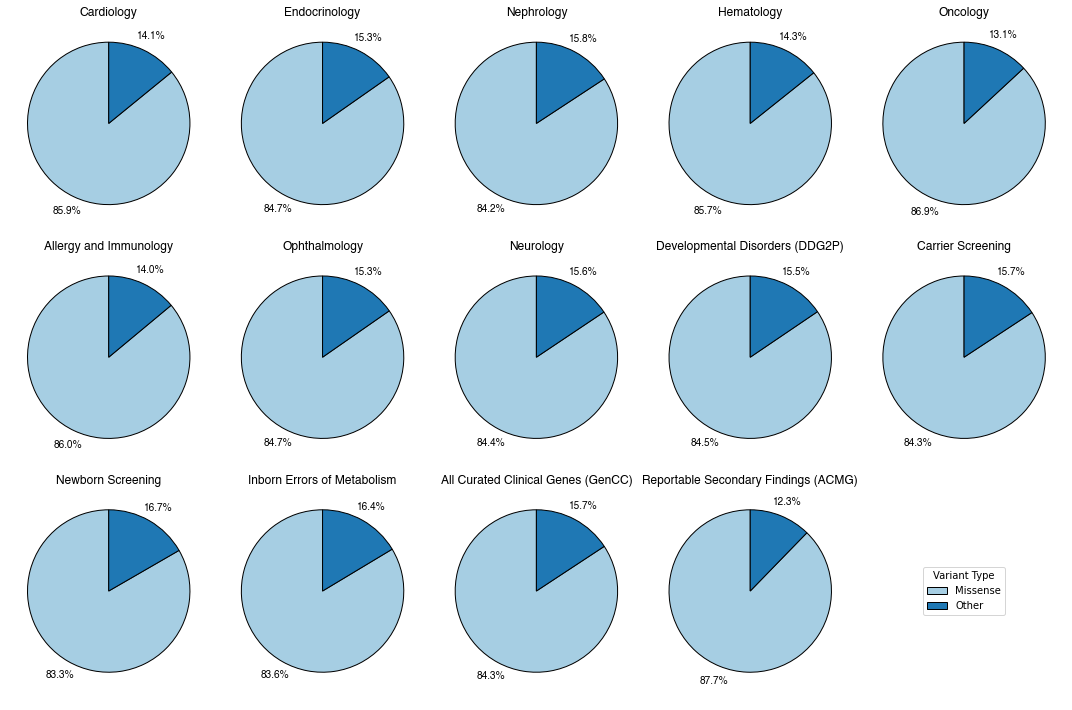

**Supplemental Figure 15b: Variant Composition of VUS Across Medical Specialties in gnomAD v2.1.1**

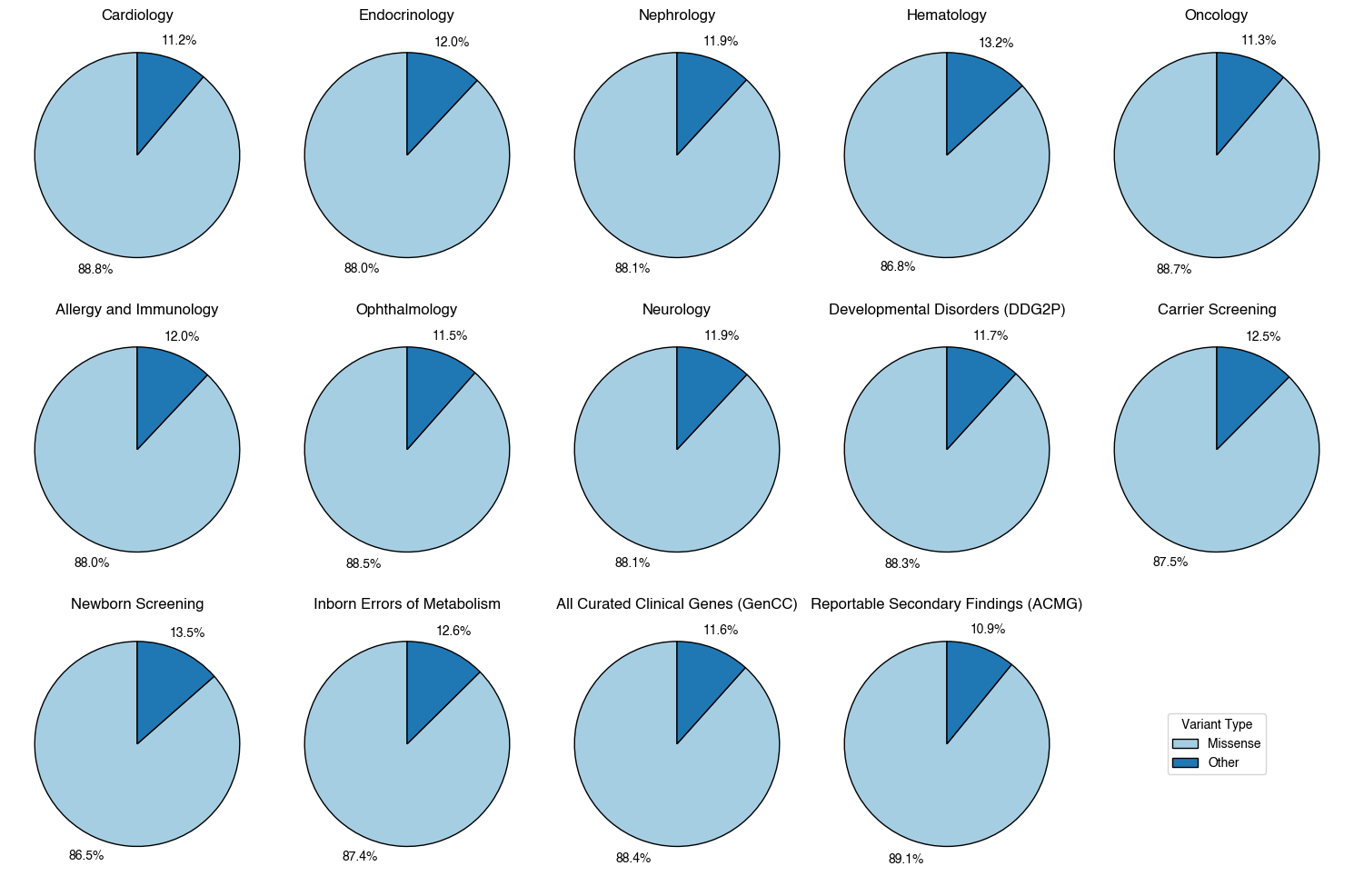

**Supplemental Figure 15c: Variant Composition of VUS Across Medical Specialties in gnomAD v3.1.2 (non v2)**

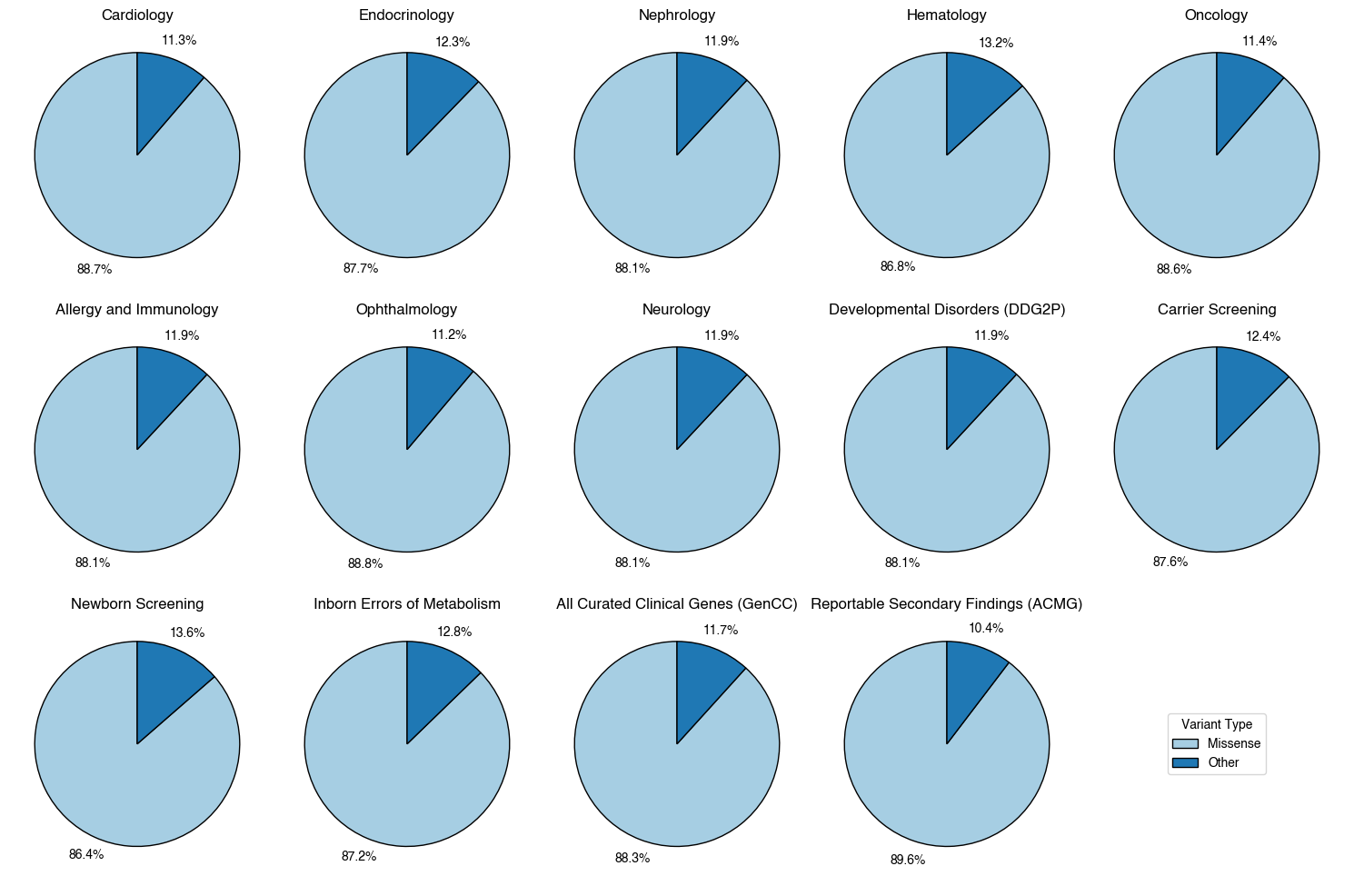

### Supplemental Figure 16: Effect Size Comparison of Missense Variants in VUS Prevalence

Effect size with 95% confidence interval (plotted and denoted on the right) shown for only the significant differences between VUS prevalence in individuals of non-European-like versus European-like genetic ancestry as measured by the rank biserial coefficient from the signed rank, matched pairs, Wilcoxon test with a Bonferroni correction. The total number of alleles from individuals of non-European-like versus European-like genetic ancestry are indicated on the left. Effect sizes in black were calculated from all coding variants while effect sizes in blue were calculated from all coding variants excluding missense variants corresponding to the medical specialty (y-axis). Thresholds as determined by Funder and Ozer (2019)^21^ for quantifying the magnitude of the effect size difference are plotted as vertical dashed line. All differences shown are statistically significant except for three (Endocrinology, Hematology, and Newborn Screening without missense variants in *All of Us* v7 although all 3 of these are significant in both gnomAD v2.1.1 and gnomAD v3.1.2 (non v2)). Also refer to Tables S10-12. Across all medical specialties, the disparity in VUS prevalence between individuals of non-European-like versus European-like genetic ancestry is not just statistically significant but very large. Further, the statistically significant disparity in VUS prevalence is still intact and large even with the exclusion of missense VUS across the medical specialties.

**Supplemental Figure 16a: Effect Size Comparison of Missense Variants in VUS Prevalence in *All of Us* v7**

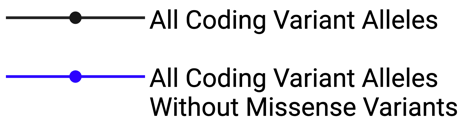

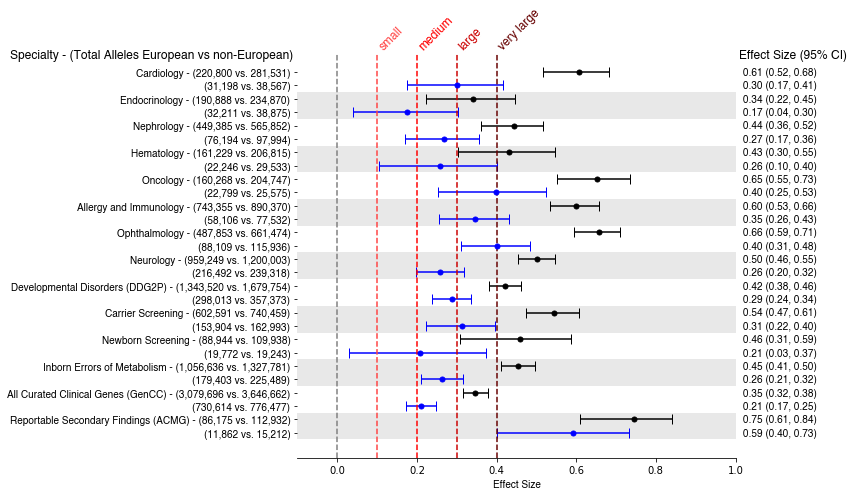

**Supplemental Figure 16b: Effect Size Comparison of Missense Variants in VUS Prevalence in gnomAD v2.1.1**

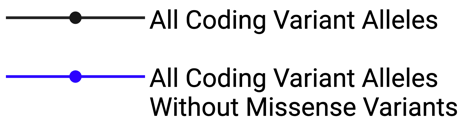

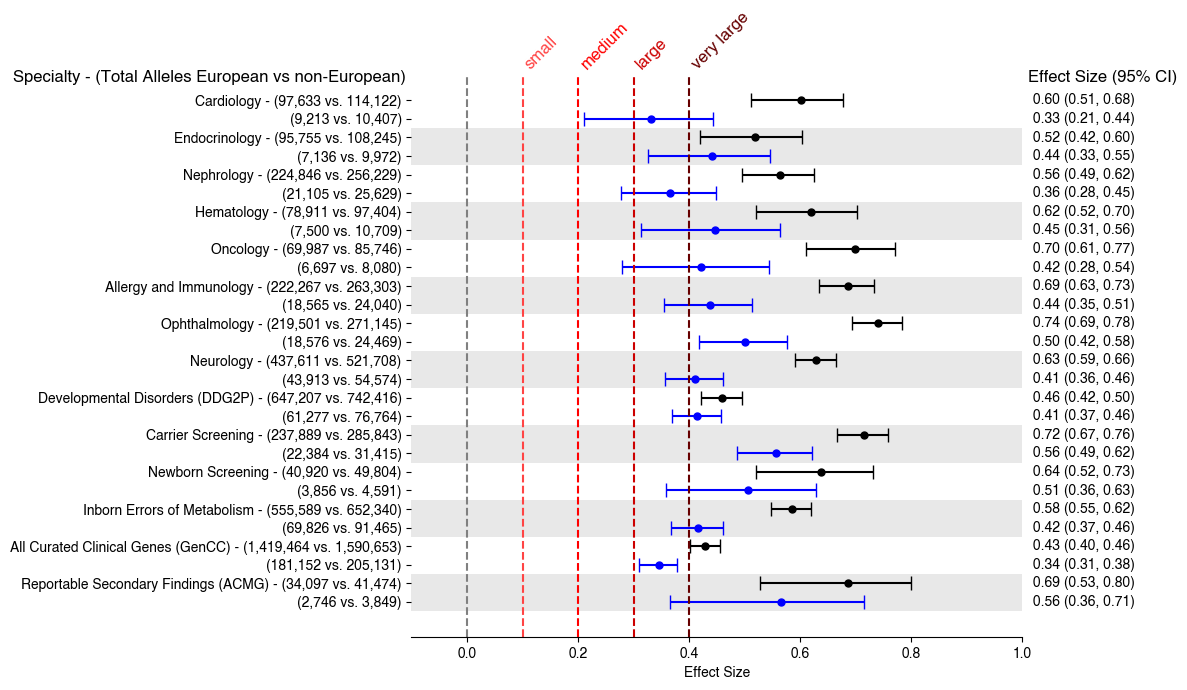

### Supplemental Figure 17: Effect Size Comparison of Missense Variants in P/LP Prevalence

Effect size with 95% confidence interval (plotted and denoted on the right) shown for only the significant differences between P/LP prevalence in individuals of non-European-like versus European-like genetic ancestry as measured by the rank biserial coefficient from the signed rank, matched pairs, Wilcoxon test with a Bonferroni correction. The total number of alleles from individuals of non-European-like versus European-like genetic ancestry are indicated on the left. Effect sizes in black were calculated from all coding variants while effect sizes in blue were calculated from all coding variants excluding missense variants corresponding to the medical specialty (y-axis). Thresholds as determined by Funder and Ozer (2019)^21^ for quantifying the magnitude of the effect size difference are plotted as vertical dashed lines. Also refer to Tables S10-12. Across several medical specialties, the disparity in P/LP prevalence between individuals of non-European-like versus European-like genetic ancestry is statistically significant for all coding variants as well as coding variants without missense variants.

**Supplemental Figure 17a: Effect Size Comparison of Pathogenic or Likely Pathogenic (P/LP) variants in *All of Us* v7**

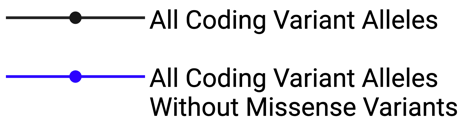

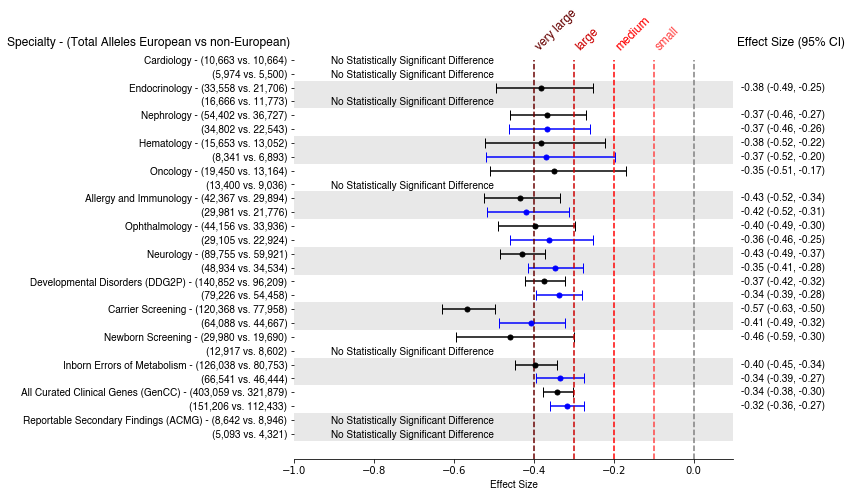

**Supplemental Figure 17b: Effect Size Comparison of P/LP variants in gnomAD v2.1.1**

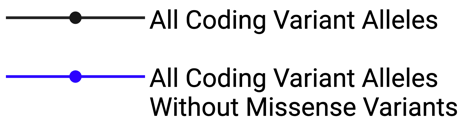

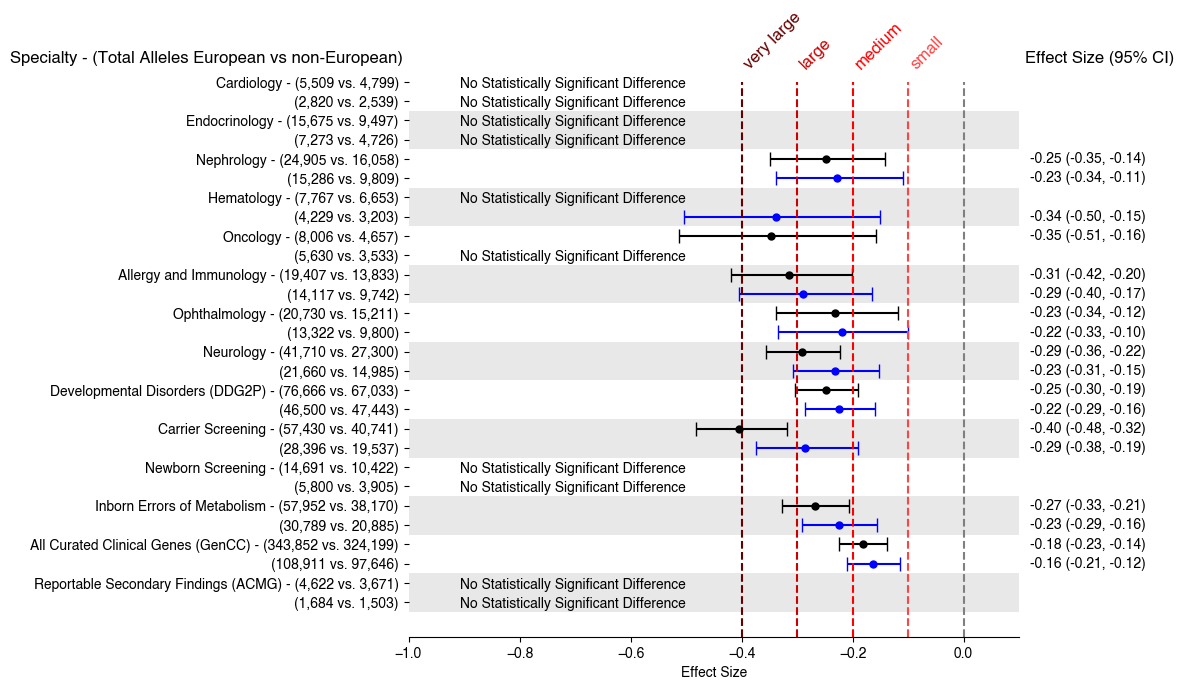

**Supplemental Figure 17c: Effect Size Comparison of P/LP variants in gnomAD v3.1.2 (non v2)**

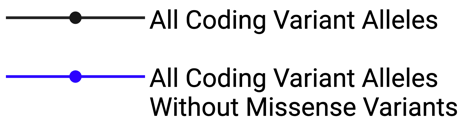

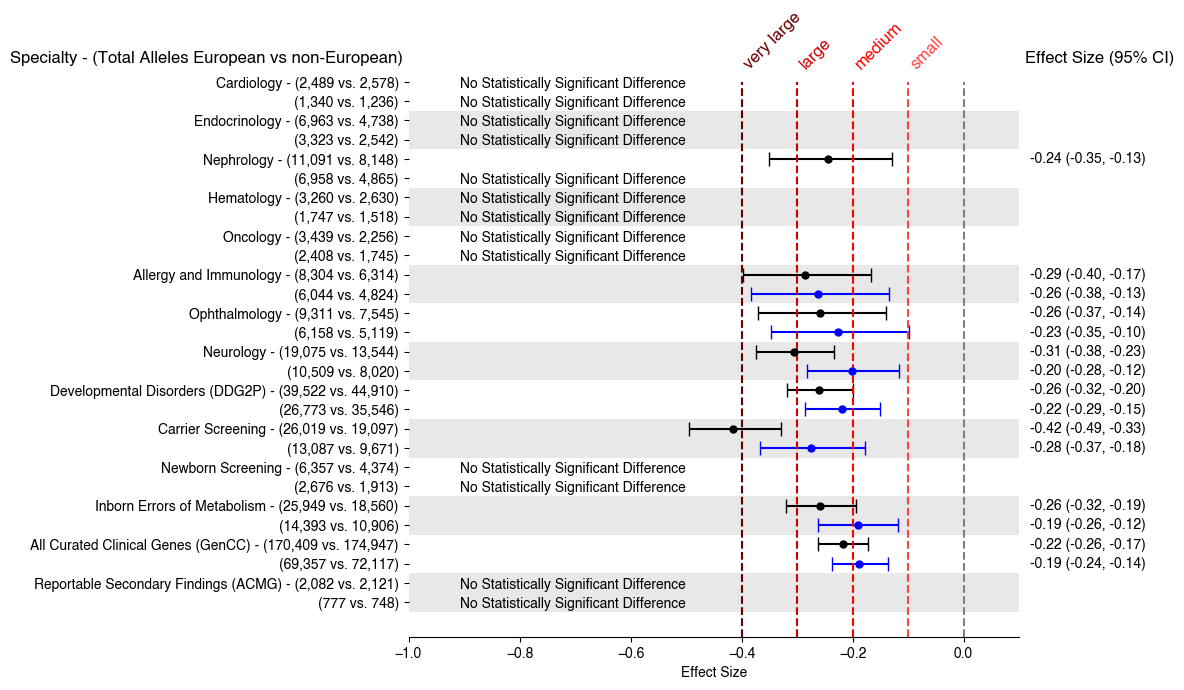

### Supplemental Figure 18: Effect Size Comparison of Benign or Likely Benign (B/LB) variants

Effect size with 95% confidence interval (plotted and denoted on the right) shown for only the significant differences between B/LB prevalence in individuals of non-European-like versus European-like genetic ancestry as measured by the rank biserial coefficient from the signed rank, matched pairs, Wilcoxon test with a Bonferroni correction (n=1000). The total number of alleles from individuals of non-European-like versus European-like genetic ancestry are indicated on the left. Effect sizes in black were calculated from all coding variants while effect sizes in blue were calculated from all coding variants excluding missense variants corresponding the medical specialty (y-axis). Thresholds as determined by Funder and Ozer (2019)^21^ for quantifying the magnitude of the effect size difference are plotted. Also refer to Tables S10-12. Overall, in the set of curated clinical genes as a whole (GenCC), the disparity in B/LB prevalence between individuals of non-European-like versus European-like genetic ancestry is not just statistically significant but intact with the exclusion of missense B/LB variants.

**Supplemental Figure 18a: Effect Size Comparison of Benign or Likely Benign (B/LB) variants in *All of Us* v7**

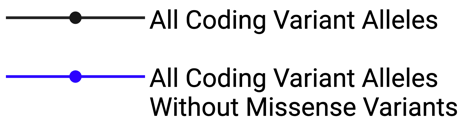

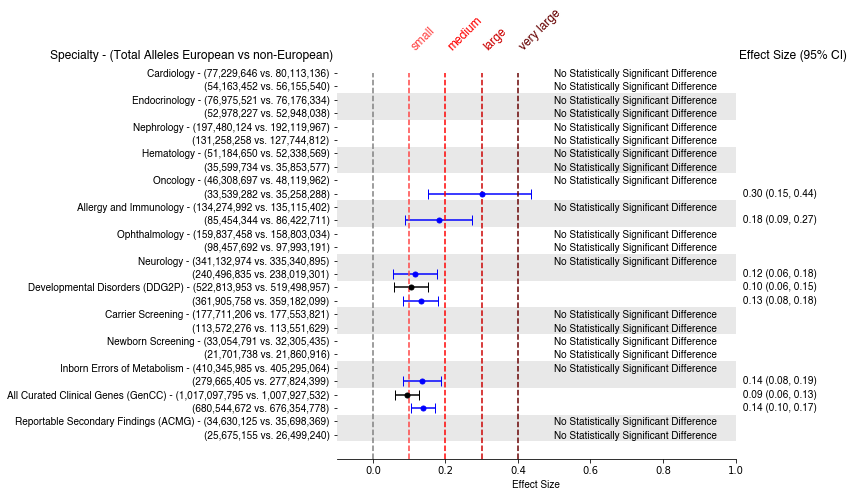

**Supplemental Figure 18b: Effect Size Comparison of B/LB variants in gnomAD v2.1.1**

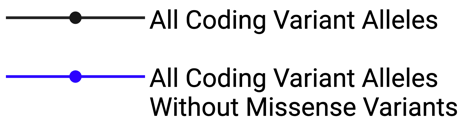

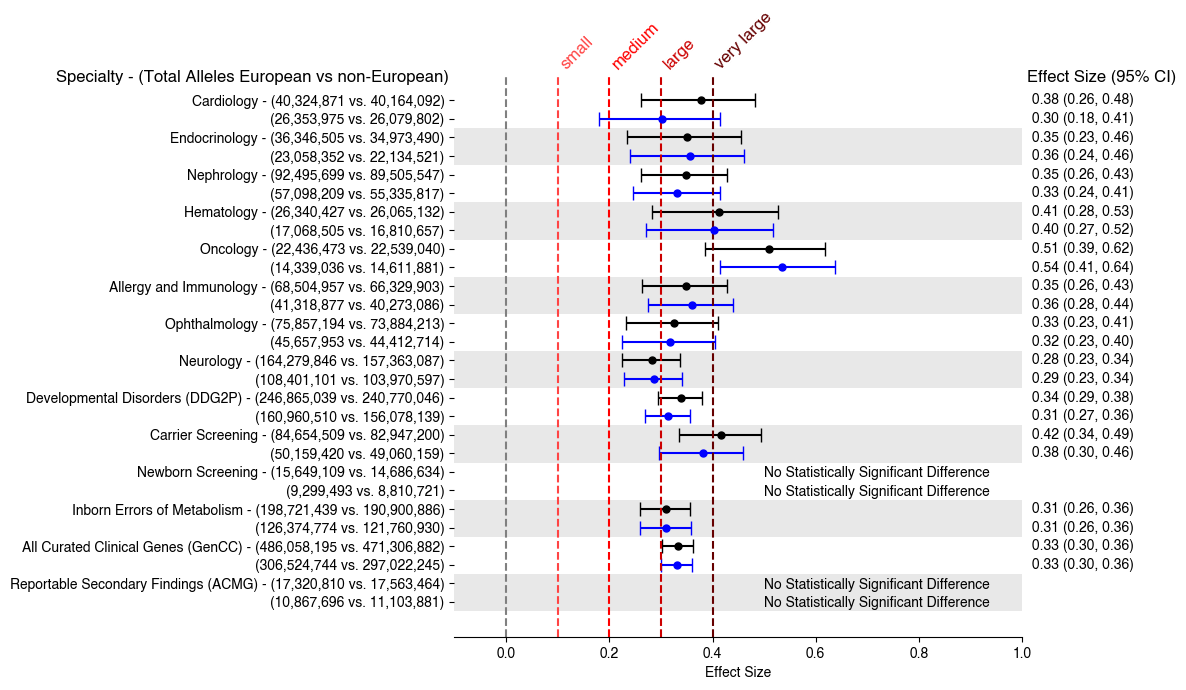

**Supplemental Figure 18c: Effect Size Comparison of B/LB variants in gnomAD v3.1.2 (non v2)**

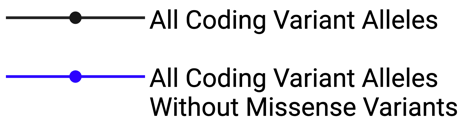

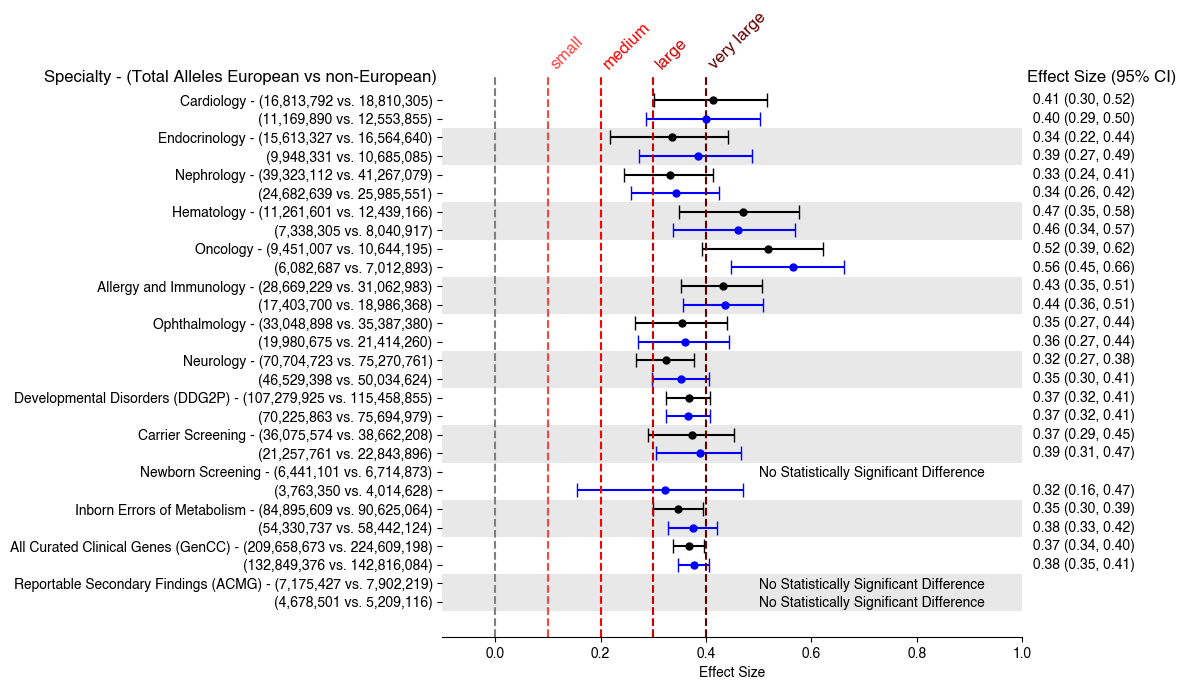

### Supplemental Figure 19: Allele Prevalence by Variant Type of Variants of Uncertain Significance (VUS)

Box plots corresponding to VUS allele prevalence (x-axis) in genes (dots) for individuals of non-European-like (blue) versus European-like (orange) genetic ancestry for the corresponding variant type (y-axis) across a) gnomAD v2.1.1 and b) gnomAD v3.1.2 (non v2) for all coding variants in the set of curated clinical genes (GenCC). The total number of alleles from individuals of non-European-like (right) versus European-like (left) genetic ancestry are indicated under each variant type in parentheses. Genes (y-axis) with zero alleles for the corresponding variant type for allele prevalence for either individuals of European-like or non-European-like genetic ancestry are omitted from the visualization to maintain a reasonable scale for data visualization. However, genes with zero alleles for only one category of either individuals of European-like or non-European-like genetic ancestry are included in the Bonferroni-corrected, signed rank, matched pairs Wilcoxon statistical test. The Bonferroni corrected p-values associated with these comparisons are annotated as follows with "ns" indicating not significant, * for 1.52e-04 < p ≤ 3.03e-04, ** for 7.58e-05 < p ≤ 1.52e-04, *** for 7.58e-06 < p ≤ 7.58e-05, and **** for p ≤ 7.58e-06. Also refer to Tables S13-15. Overall, we observe a statistically significant increase in VUS in individuals of non-European-like genetic ancestry compared to individuals of European-like genetic ancestry for missense, synonymous, splice region, and inframe variants.

**Supplemental Figure 19a: Allele Prevalence by Variant Type of VUS in *All of Us* v7**

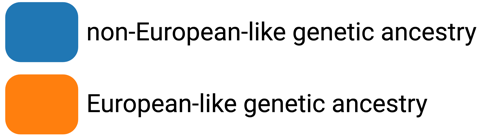

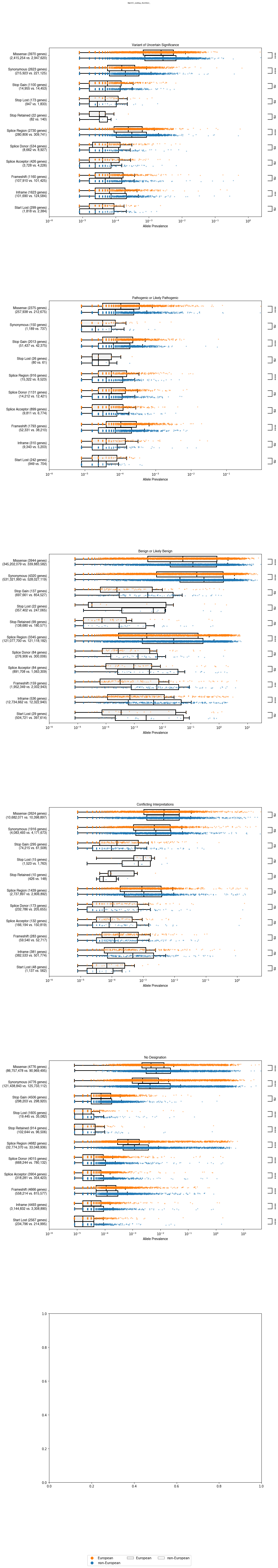

**Supplemental Figure 19b: Allele Prevalence by Variant Type of VUS in gnomAD v2.1.1**

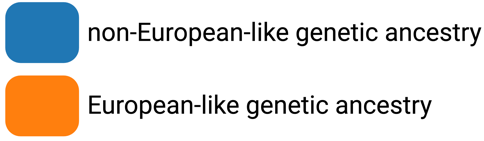

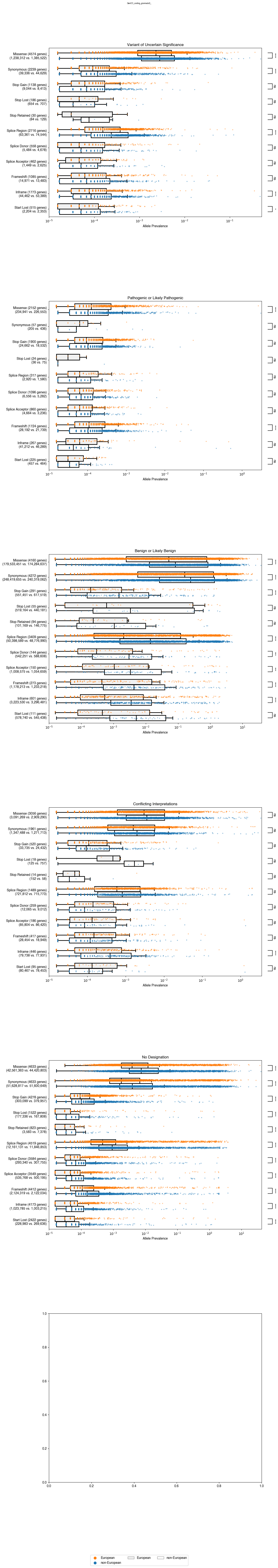

**Supplemental Figure 19c: Allele Prevalence by Variant Type of VUS in gnomAD v3.1.2 (non v2)**

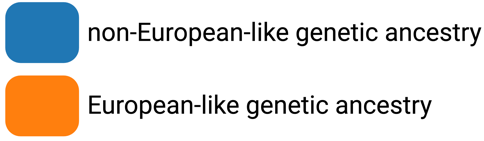

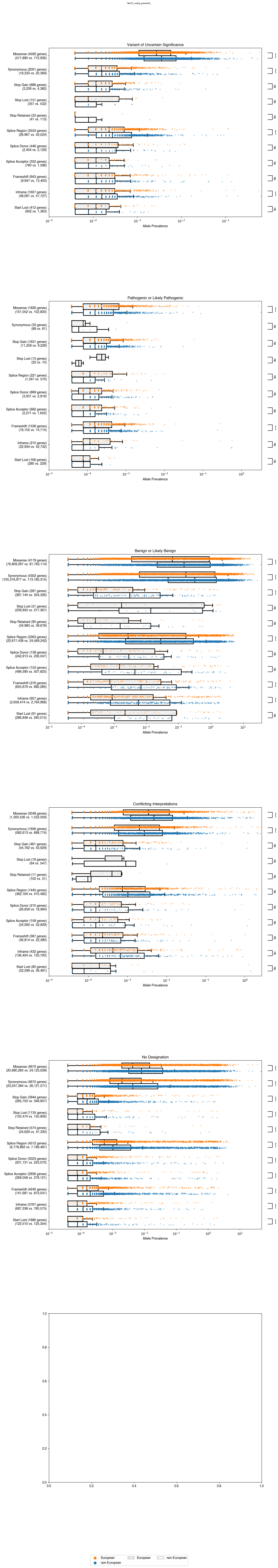

### Supplemental Figure 20: Allele Prevalence by Variant Type of Pathogenic or Likely Pathogenic (P/LP) Variants

Box plots corresponding to P/LP allele prevalence (x-axis) in genes (dots) for individuals of non-European-like (blue) versus European-like (orange) genetic ancestry for the corresponding variant type (y-axis) across a) *All of Us* v7, b) gnomAD v2.1.1, and c) gnomAD v3.1.2 (non v2) for all coding variants in the set of curated clinical genes (GenCC). The total number of alleles from individuals of non-European-like (right) versus European-like (left) genetic ancestry are indicated under each variant type in parentheses. Genes (y-axis) with zero alleles for the corresponding variant type for allele prevalence for either individuals of European-like or non-European-like genetic ancestry are omitted from the visualization to maintain a reasonable scale for data visualization. However, genes with zero alleles for only one category of either individuals of European-like or non-European-like genetic ancestry are included in the Bonferroni-corrected, signed rank, matched pairs Wilcoxon statistical test. The Bonferroni corrected p-values associated with these comparisons are annotated as follows with "ns" indicating not significant, * for 1.52e-04 < p ≤ 3.03e-04, ** for 7.58e-05 < p ≤ 1.52e-04, *** for 7.58e-06 < p ≤ 7.58e-05, and **** for p ≤ 7.58e-06. Also refer to Tables S13-15. Overall, we observe a statistically significant increase in P/LP variants in individuals of European-like genetic ancestry compared to individuals of non-European-like genetic ancestry for missense and frameshift variants.

**Supplemental Figure 20a: Allele Prevalence by Variant Type of P/LP Variants in *All of Us* v7**

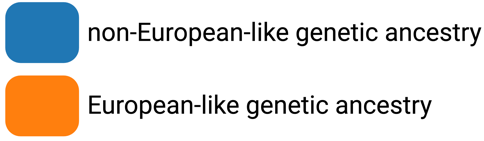

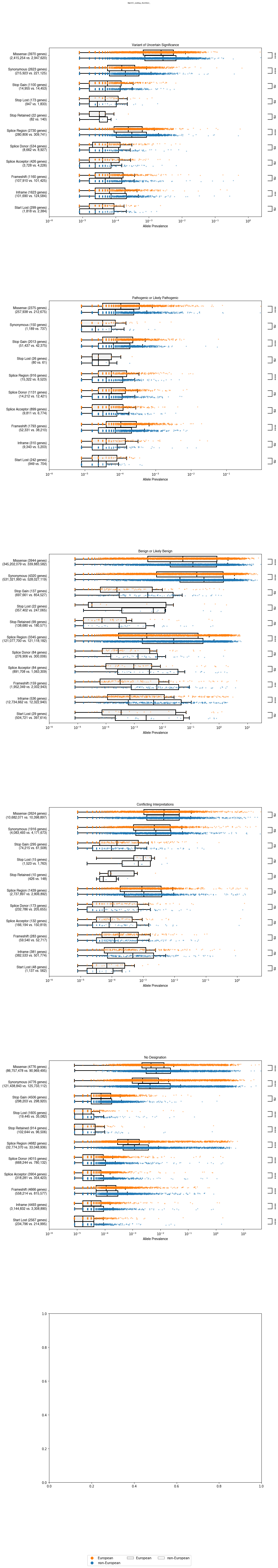

**Supplemental Figure 20b: Allele Prevalence by Variant Type of P/LP Variants in gnomAD v2.1.1**

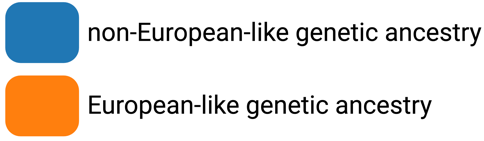

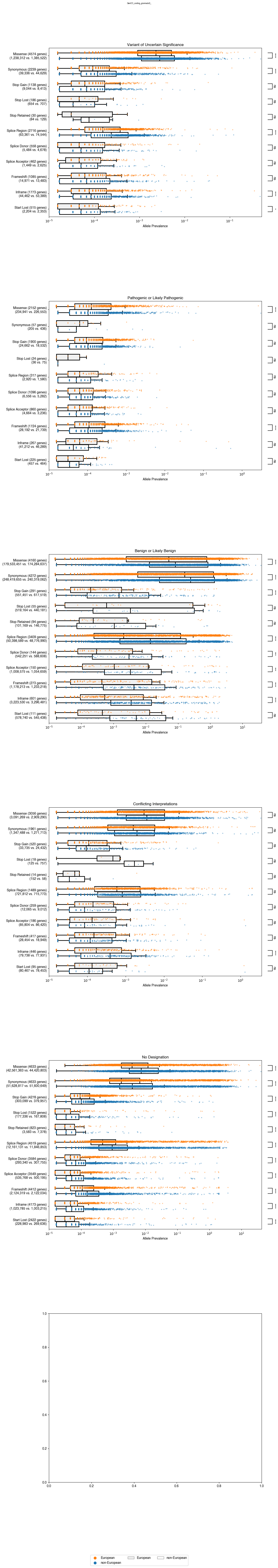

**Supplemental Figure 20c: Allele Prevalence by Variant Type of P/LP Variants in gnomAD v3.1.2 (non v2)**

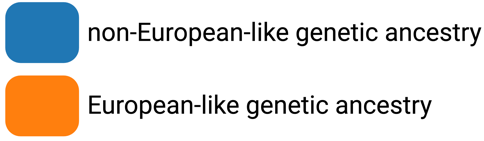

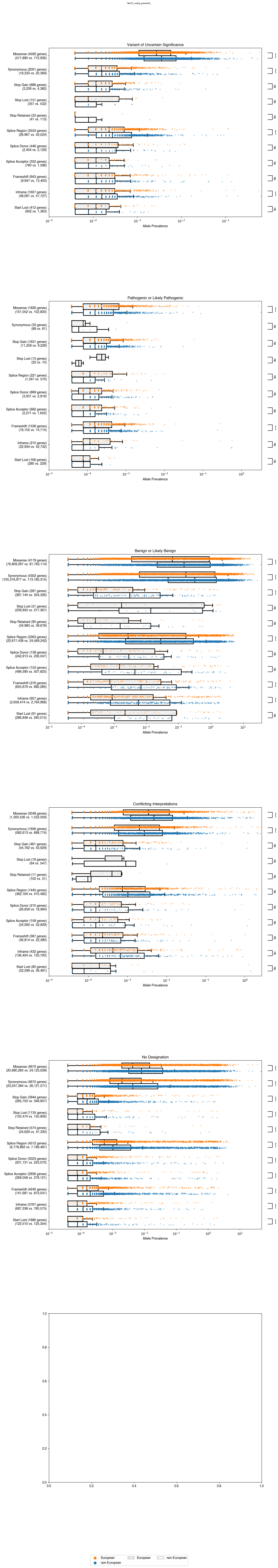

### Supplemental Figure 21: Allele Prevalence by Variant Type of Benign or Likely Benign (B/LB) Variants

Box plots corresponding to B/LB allele prevalence (x-axis) in genes (dots) for individuals of non-European-like (blue) versus European-like (orange) genetic ancestry for the corresponding variant type (y-axis) across a) *All of Us* v7, b) gnomAD v2.1.1, and c) gnomAD v3.1.2 (non v2) for all coding variants in the set of curated clinical genes (GenCC). The total number of alleles from individuals of non-European-like (right) versus European-like (left) genetic ancestry are indicated under each variant type in parentheses. Genes (y-axis) with zero alleles for the corresponding variant type for allele prevalence for either individuals of European-like or non-European-like genetic ancestry are omitted from the visualization to maintain a reasonable scale for data visualization. However, genes with zero alleles for only one category of either individuals of European-like or non-European-like genetic ancestry are included in the Bonferroni-corrected, signed rank, matched pairs Wilcoxon statistical test. The Bonferroni corrected p-values associated with these comparisons are annotated as follows with "ns" indicating not significant, * for 1.52e-04 < p ≤ 3.03e-04, ** for 7.58e-05 < p ≤ 1.52e-04, *** for 7.58e-06 < p ≤ 7.58e-05, and **** for p ≤ 7.58e-06. Also refer to Tables S13-15. Overall, we observe a statistically significant increase in B/LB allele prevalence in individuals of non-European-like genetic ancestry compared to individuals of European-like genetic ancestry for a variety of variant types.

**Supplemental Figure 21a: Allele Prevalence by Variant Type of B/LB Variants in *All of Us* v7**

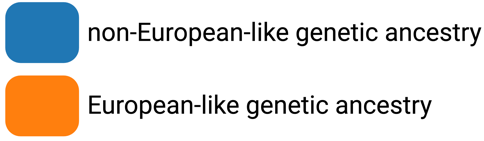

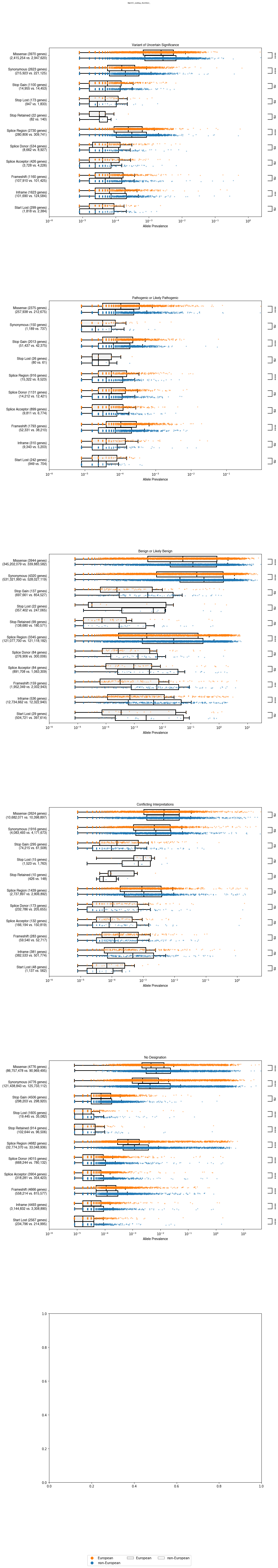

**Supplemental Figure 21b: Allele Prevalence by Variant Type of B/LB Variants in gnomAD v2.1.1**

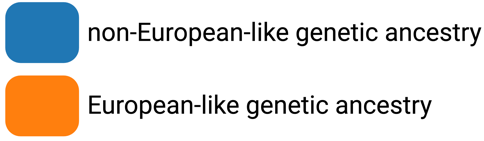

.

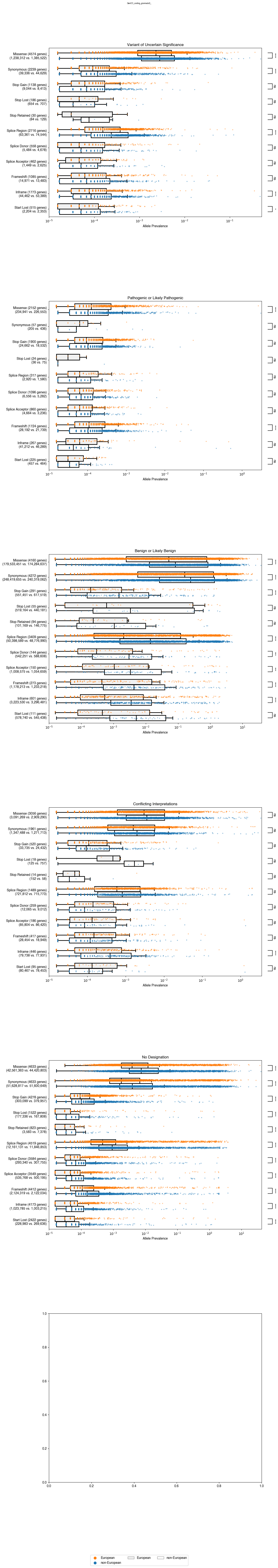

**Supplemental Figure 21c: Allele Prevalence by Variant Type of B/LB Variants in gnomAD v3.1.2 (non v2)**

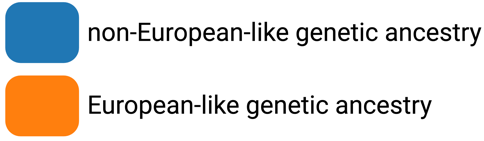

### Supplemental Figure 22: Allele Prevalence by Variant Type of Conflicting Interpretation (CI) Variants

Box plots corresponding to CI allele prevalence (x-axis) in genes (dots) for individuals of non-European-like (blue) versus European-like (orange) genetic ancestry for the corresponding variant type (y-axis) across a) *All of Us* v7, b) gnomAD v2.1.1, and c) gnomAD v3.1.2 (non v2) for all coding variants in the set of curated clinical genes (GenCC). The total number of alleles from individuals of non-European-like (right) versus European-like (left) genetic ancestry are indicated under each variant type in parentheses. Genes (y-axis) with zero alleles for the corresponding variant type for allele prevalence for either individuals of European-like or non-European-like genetic ancestry are omitted from the visualization to maintain a reasonable scale for data visualization. However, genes with zero alleles for only one category of either individuals of European-like or non-European-like genetic ancestry are included in the Bonferroni-corrected, signed rank, matched pairs Wilcoxon statistical test. The Bonferroni corrected p-values associated with these comparisons are annotated as follows with "ns" indicating not significant, * for 1.52e-04 < p ≤ 3.03e-04, ** for 7.58e-05 < p ≤ 1.52e-04, *** for 7.58e-06 < p ≤ 7.58e-05, and **** for p ≤ 7.58e-06. Also refer to Tables S13-15. Overall, while some variant types are observed to be statistically significant in individual databases, there are no clear trends across all three databases for increases in CI allele prevalence in individuals of non-European-like genetic ancestry compared to individuals of European-like genetic ancestry.

**Supplemental Figure 22a: Allele Prevalence by Variant Type of CI Variants in *All of Us* v7**

**Supplemental Figure 22b: Allele Prevalence by Variant Type of CI Variants in gnomAD v2.1.1**

**Supplemental Figure 22c: Allele Prevalence by Variant Type of CI Variants in gnomAD v3.1.2 (non v2)**

### Supplemental Figure 23: Allele Prevalence by Variant Type of No Designation (ND) Variants

Box plots corresponding to ND allele prevalence (x-axis) in genes (dots) for individuals of non-European-like (blue) versus European-like (orange) genetic ancestry for the corresponding variant type (y-axis) across a) *All of Us* v7, b) gnomAD v2.1.1, and c) gnomAD v3.1.2 (non v2) for all coding variants in the set of curated clinical genes (GenCC). The total number of alleles from individuals of non-European-like (right) versus European-like (left) genetic ancestry are indicated under each variant type in parentheses. Genes (y-axis) with zero alleles for the corresponding variant type for allele prevalence for either individuals of European-like or non-European-like genetic ancestry are omitted from the visualization to maintain a reasonable scale for data visualization. However, genes with zero alleles for only one category of either individuals of European-like or non-European-like genetic ancestry are included in the Bonferroni-corrected, signed rank, matched pairs Wilcoxon statistical test. The Bonferroni corrected p-values associated with these comparisons are annotated as follows with "ns" indicating not significant, * for 1.52e-04 < p ≤ 3.03e-04, ** for 7.58e-05 < p ≤ 1.52e-04, *** for 7.58e-06 < p ≤ 7.58e-05, and **** for p ≤ 7.58e-06. Also refer to Tables S13-15. Overall, we observe a statistically significant increase in ND allele prevalence in individuals of non-European-like genetic ancestry compared to individuals of European-like genetic ancestry for all variant types.

**Supplemental Figure 23a: Allele Prevalence by Variant Type of ND Variants in *All of Us* v7**

**Supplemental Figure 23b: Allele Prevalence by Variant Type of ND Variants in gnomAD v2.1.1**

**Supplemental Figure 23c: Allele Prevalence by Variant Type of ND Variants in gnomAD v3.1.2 (non v2)**

### Supplemental Figure 24: Allele Prevalence Comparison of Noncoding Variant Types

Box plots corresponding to allele prevalence (x-axis) for each gene (dot) for individuals of non-European-like (blue) versus European-like (orange) genetic ancestry for the corresponding medical specialty (y-axis) across a) gnomAD v2.1.1 and b) gnomAD v3.1.2 (non v2) for all noncoding variants. The total number of alleles from individuals of non-European-like (right) versus European-like (left) genetic ancestry are indicated under each variant type in parentheses. Genes with zero alleles for allele prevalence for either individuals of European-like or non-European-like genetic ancestry are omitted from the above visualization to maintain a reasonable scale for data visualization. However, genes with zero alleles for only one category of either individuals of European-like or non-European-like genetic ancestry are included in the Bonferroni-corrected, signed rank, matched pairs Wilcoxon statistical test. The Bonferroni corrected p-values associated with these comparisons are annotated according to the provided legend, with "ns" indicating not significant, * for 1.79e-04 < p ≤ 3.57e-04, ** for 8.93e-05 < p ≤ 1.79e-04, *** for 8.93e-06 < p ≤ 8.93e-05, and **** for p ≤ 8.93e-06. Also refer to Tables S16-17. Across all medical specialties and categories shown, ND and B/LB variants are observed to be statistically significantly increased in individuals of non-European-like genetic ancestry compared to individuals of European-like genetic ancestry.

**Supplemental Figure 24a: Allele Prevalence Comparison of Noncoding Variant Types in gnomAD v2.1.1**

**Supplemental Figure 24b: Allele Prevalence Comparison of Noncoding Variant Types in gnomAD v3.1.2 (non v2)**

### Supplemental Figure 25: Comparison of Counts of Unique Variants Found In Only One Genetic Ancestry Group

Grouped bar graphs corresponding to unique coding variant counts (y-axis) for a) VUS, b) P/LP, c) B/LB, d) CI and e) ND variants found either only in individuals of European-like (orange) genetic ancestry or only in individuals of non-European-like (blue) genetic ancestry across the medical specialties (x-axis) in *All of Us* v7, gnomAD v2.1.1 and gnomAD v3.1.2 (non v2). The Bonferroni corrected p-values from the chi-square test of independence associated with these comparisons are annotated along with the estimated statistical power. Also refer to Tables S2-4. Across all medical specialties and categories shown, VUS, B/LB, CI and ND variants were found at a statistically significantly higher prevalence in individuals of non-European-like genetic ancestry in all 3 databases with an estimated statistical power near 1. Conversely P/LP variants were found at a statistically significantly higher prevalence in individuals of European-like genetic ancestry in both the *All of Us* v7 and gnomAD v2.1.1 databases but not quite in gnomAD v3.1.2 (non v2). However, the low statistical power coupled with the much smaller sample of individuals sequenced in gnomAD v3.1.2 (non v2) along with the observation that there are more P/LP variants in each specialty for gnomAD v3.1.2 (non v2) just as in gnomAD v2.1.1 and *All of Us* v7 suggest that this particular statistical test is underpowered to detect any potential significant difference.

**Supplemental Figure 25a: Unique VUS Differences Between Individuals of European-like Versus Non-European-like Genetic Ancestry**

**Supplemental Figure 25b: Unique P/LP Differences Between Individuals of European-like Versus Non-European-like Genetic Ancestry**

**Supplemental Figure 25c: Unique B/LB Differences Between Individuals of European-like Versus Non-European-like Genetic Ancestry**

**Supplemental Figure 25d: Unique CI Differences Between Individuals of European-like Versus Non-European-like Genetic Ancestry**

**Supplemental Figure 25e: Unique ND Differences Between Individuals of European-like Versus Non-European-like Genetic Ancestry**

### Supplemental Figure 26: Venn Diagrams of Unique Variants Across Medical Specialties And Clinical Designation

Venn Diagrams corresponding to VUS (column 1 from left to right), P/LP (column 2), B/LB (column 3), CI (column 4) and ND (column 5) variants found either only in individuals of European-like (orange) genetic ancestry or only in individuals of non-European-like (blue) genetic ancestry across the medical specialties (y-axis) for all unique coding variant counts in a) *All of Us* v7, b) gnomAD v2.1.1 and c) gnomAD v3.1.2 (non v2) and all unique coding variants without missense variants in d) *All of Us* v7, e) gnomAD v2.1.1 and f) gnomAD v3.1.2 (non v2). The Bonferroni corrected p-values from the chi-square test of independence associated with these comparisons are annotated along with the estimated statistical power. Also refer to Tables S2-4, S10-12. Across all medical specialties and categories shown, the overlap of unique variants for VUS, P/LP, B/LB, and CI variants is statistically significantly greater than the overlap between ND variants in all 3 databases across all medical specialties in both the presence and absence of missense variants.

**Supplemental Figure 26a: Venn Diagrams of Unique Variants Across Medical Specialties for *All of Us* v7**

| Cardiology |
| --- |
| Endocrinology |
| Nephrology |
| Hematology |
| Oncology |
| Allergy and Immunology |
| Ophthalmology |
| Neurology |
| Developmental Disorders (DDG2P) |
| Carrier Screening |
| Newborn Screening |
| Inborn Errors of Metabolism |
| All Curated Clinical Genes (GenCC) |
| Reportable Secondary Findings (ACMG) |

**Supplemental Figure 26b: Venn Diagrams of Unique Variants Across Medical Specialties for gnomAD v2.1.1**

| Cardiology |
| --- |
| Endocrinology |
| Nephrology |
| Hematology |
| Oncology |
| Allergy and Immunology |
| Ophthalmology |
| Neurology |
| Developmental Disorders (DDG2P) |
| Carrier Screening |
| Newborn Screening |
| Inborn Errors of Metabolism |
| All Curated Clinical Genes (GenCC) |
| Reportable Secondary Findings (ACMG) |

**Supplemental Figure 26c: Venn Diagrams of Unique Variants Across Medical Specialties for gnomAD v3.1.2 (non v2)**

| Cardiology |
| --- |
| Endocrinology |
| Nephrology |
| Hematology |
| Oncology |
| Allergy and Immunology |
| Ophthalmology |
| Neurology |
| Developmental Disorders (DDG2P) |
| Carrier Screening |
| Newborn Screening |
| Inborn Errors of Metabolism |
| All Curated Clinical Genes (GenCC) |
| Reportable Secondary Findings (ACMG) |

**Supplemental Figure 26d: Venn Diagrams of Unique Variants Without Missense Variants Across Medical Specialties for *All of Us* v7**

| Cardiology |
| --- |
| Endocrinology |
| Nephrology |
| Hematology |
| Oncology |
| Allergy and Immunology |
| Ophthalmology |
| Neurology |
| Developmental Disorders (DDG2P) |
| Carrier Screening |
| Newborn Screening |
| Inborn Errors of Metabolism |
| All Curated Clinical Genes (GenCC) |
| Reportable Secondary Findings (ACMG) |

**Supplemental Figure 26e: Venn Diagrams of Unique Variants Without Missense Variants Across Medical Specialties for gnomAD v2.1.1**

| Cardiology |
| --- |
| Endocrinology |
| Nephrology |
| Hematology |
| Oncology |
| Allergy and Immunology |
| Ophthalmology |
| Neurology |
| Developmental Disorders (DDG2P) |
| Carrier Screening |
| Newborn Screening |
| Inborn Errors of Metabolism |
| All Curated Clinical Genes (GenCC) |
| Reportable Secondary Findings (ACMG) |

**Supplemental Figure 26f: Venn Diagrams of Unique Variants Without Missense Across Medical Specialties for gnomAD v3.1.2 (non v2)**

| Cardiology |
| --- |
| Endocrinology |
| Nephrology |
| Hematology |
| Oncology |
| Allergy and Immunology |
| Ophthalmology |
| Neurology |
| Developmental Disorders (DDG2P) |
| Carrier Screening |
| Newborn Screening |
| Inborn Errors of Metabolism |
| All Curated Clinical Genes (GenCC) |
| Reportable Secondary Findings (ACMG) |

### Supplemental Figure 27: Comparison of Allele Prevalence for Each Non-European-like Genetic Ancestry Across Differing Clinical Significance Categories

Box plots corresponding to a) VUS, b) B/LB, c) CI, d) ND allele prevalence (x-axis) in each gene (dot) for each non-European-like (shades of blue) genetic ancestry versus European-like (orange) genetic ancestry for all curated clinical genes (GenCC list; y-axis) across all three population databases (*All of Us* v7, gnomAD v2.1.1 and gnomAD v3.1.2 (non v2)) for all coding variants. Genes with zero alleles for allele prevalence for either individuals of European-like or non-European-like genetic ancestry are omitted from the above visualization to maintain a reasonable scale for data visualization. However, genes with zero alleles for only one category of either individuals of European-like or non-European-like genetic ancestry are included in the Bonferroni-corrected, signed rank, matched pairs Wilcoxon statistical test. The Bonferroni corrected p-values associated with these comparisons are annotated as follows with "ns" indicating not significant, * for 3.33e-04 < p ≤ 6.67e-04, ** for 1.67e-04 < p ≤ 3.33e-04, *** for 1.67e-05 < p ≤ 1.67e-04, and **** for p ≤ 1.67e-05. A higher prevalence of VUS were found in individuals of African/African American and Latino/Admixed American genetic ancestry. A higher prevalence of B/LB variants were found in individuals of African/African American Genetic Ancestry. A higher prevalence of CI variants were found in individuals of Latino/Admixed American, East Asian, and South Asian genetic ancestry. A higher prevalence of ND variants were found in individuals of African/African American, Latino/Admixed American, East Asian, and South Asian genetic ancestry.

**Supplemental Figure 27a: Higher Prevalence of P/LP Found in Individuals of European-like Genetic Ancestry**

| gnomAD v2.1.1 |
| --- |
| gnomAD v3.1.2 (non v2) |

**Supplemental Figure 27b: Higher Prevalence of VUS Found in Individuals of African/African American and Latino/Admixed American Genetic Ancestry**

| *All of Us* v7 |
| --- |
| gnomAD v2.1.1 |
| gnomAD v3.1.2 (non v2) |

**Supplemental Figure 27c: Higher Prevalence of B/LB Variants Found in Individuals of African/African American Genetic Ancestry**

| *All of Us* v7 |
| --- |
| gnomAD v2.1.1 |
| gnomAD v3.1.2 (non v2) |

**Supplemental Figure 27d: Higher Prevalence of CI Variants Found in Individuals of Latino/Admixed American, East Asian, and South Asian Genetic Ancestry**

| *All of Us* v7 |
| --- |
| gnomAD v2.1.1 |
| gnomAD v3.1.2 (non v2) |

**Supplemental Figure 27e: Higher Prevalence of ND Variants Found in Individuals of African/African American, Latino/Admixed American, East Asian, and South Asian Genetic Ancestry**

| *All of Us* v7 |
| --- |
| gnomAD v2.1.1 |
| gnomAD v3.1.2 (non v2) |

| Supplemental Figure 28: Evidence Code Usage for Variant Reclassification | | | | | | |
| --- | --- | --- | --- | --- | --- | --- |
|  | *BRCA1* | | *TP53* | | *PTEN* | |
| **Evidence Code** | Data Unavailable for Evidence Code | VCEP Rules | Data Unavailable for Evidence Code | VCEP Rules | Data Unavailable for Evidence Code | VCEP Rules |
| PVS1 |  | x |  | x |  | x |
| PS1 |  | x |  | x |  | x |
| PS2 |  | x | x |  | x |  |
| PS3 | Clinically Calibrated MAVE Data | | | | | |
| PS4 | x |  | x |  | x |  |
| PM1 |  | x |  | x |  | x |
| PM2 |  | x |  | x |  | x |
| PM3 | x |  |  | x |  | x |
| PM4 |  | x |  | x | x |  |
| PM5 | x |  |  | x |  | x |
| PM6 |  | x | x |  | x |  |
| PP1 | x |  | x |  | x |  |
| PP2 |  | x |  | x |  | x |
| PP3 |  | x |  | x |  | x |
| PP4 | x |  | x |  |  | x |
| PP5 |  | x |  | x |  | x |
| BA1 |  | x |  | x |  | x |
| BS1 |  | x |  | x |  | x |
| BS2 | x |  | x |  | x |  |
| BS3 | Clinically Calibrated MAVE Data | | | | | |
| BS4 | x |  | x |  | x |  |
| BP1 |  | x |  | x |  | x |
| BP2 |  | x |  | x | x |  |
| BP3 |  | x |  | x |  | x |
| BP4 |  | x |  | x |  | x |
| BP5 | x |  |  | x | x |  |
| BP6 |  | x |  | x |  | x |
| BP7 |  | x |  | x | x |  |
| Combining Criteria |  | x |  | x |  | x |

### Supplemental Figure 29: Sankey Flow Figures Showing VUS Reclassification in Each Gene

Sankey flow diagrams depicting VUS reclassification (read from left to right) for individuals of European-like (top) versus non-European-like (bottom) genetic ancestry before reclassification (No MAVE) and after reclassification (With MAVE) for a) *BRCA1*, b) *TP53* and c) *PTEN*. The VUS were reclassified as either Likely Benign (LB; light blue), Benign (B; dark blue), Likely Pathogenic (LP; red), or remained as Variants of Uncertain Significance (VUS; gray). The VUS for these three genes are the total alleles summed from all three databases *All of Us* v7, gnomAD v2.1.1, and gnomAD v3.1.2 (non v2). Reclassification was conducted using an automated pipeline based on the Variant Curation Expert Panel gene specific variant interpretation guidelines for each gene. Also refer to Table S19.

**Supplemental Figure 29a: Reclassification of *BRCA1* VUS in Individuals of European-like Versus non-European-like Genetic Ancestry**

**Supplemental Figure 29b: Reclassification of *TP53* VUS in Individuals of European-like Versus non-European-like Genetic Ancestry**

**Supplemental Figure 29c: Reclassification of *PTEN* VUS in Individuals of European-like Versus non-European-like Genetic Ancestry**

### Supplemental Figure 30: Sankey Flow Figures Showing VUS Reclassification in Each Database

Sankey flow diagrams depicting VUS reclassification (read from left to right) for individuals of European-like (top) versus non-European-like (bottom) genetic ancestry before reclassification (No MAVE) and after reclassification (With MAVE) for a) *All of Us* v7, b) gnomAD v2.1.1 and c) gnomAD v3.1.2 (non v2). The VUS were reclassified as either Likely Benign (LB; light blue), Benign (B; dark blue), Likely Pathogenic (LP; red), or remained as Variants of Uncertain Significance (VUS; gray). The VUS for these three genes are the total alleles summed across all three genes *BRCA1*, *PTEN*, and *TP53*. Reclassification was conducted using an automated pipeline based on the Variant Curation Expert Panel gene specific variant interpretation guidelines for each gene. Also refer to Table S19.

**Supplemental Figure 30a: VUS Reclassification in Individuals of European-like vs. Non-European-like Genetic Ancestry in *All of Us* v7**

**Supplemental Figure 30b: VUS Reclassification in Individuals of European-like Versus non-European-like Genetic Ancestry in gnomAD v2.1.1**

**Supplemental Figure 30c: VUS Reclassification in Individuals of European-like Versus non-European-like Genetic Ancestry in gnomAD v3.1.2 (non v2)**

### Supplemental Figure 31: Evidence Code Usage for Variant Reclassification Along With Determination of Essential Code Usage

Bar graphs for each evidence code (x-axis) used in VUS reclassification across *BRCA1*, *TP53*, and *PTEN* for all three databases, *All of Us* v7, gnomAD v2.1.1, and gnomAD v3.1.2 (non v2). Blue bars represent alleles from individuals of non-European-like genetic ancestry, whereas orange bars represent alleles from individuals of European-like genetic ancestry. Shading represents essential codes, codes which if removed from the set of evidence codes used to reclassify the VUS would cause the variant to regress back to VUS. PP3, PP3_Moderate, and BP4 correspond to the computational predictor codes. PS3, PS3_Moderate, BS3, BS3_Moderate, BS3_Supporting corresponded to the MAVE evidence codes. BA1, BS1, BS1_Supporting correspond to the allele frequency codes. Also refer to Table S19.

### Supplemental Figure 32: Evidence Code Usage for Variant Reclassification Along With Determination of Essential Code Usage in Fayer et al. (2021)^13^

Bar graphs for each evidence code or evidence code category (x-axis) used in VUS reclassification across *BRCA1*, *TP53*, and *PTEN* in Fayer et al. (2021). Blue bars represent alleles from individuals of non-European-like genetic ancestry, whereas orange bars represent alleles from individuals of European-like genetic ancestry. Shading represents essential codes, codes which if removed from the set of evidence codes used to reclassify the VUS would cause the variant to regress back to VUS. PP3, PP3_Moderate, and BP4 correspond to the computational predictor codes. PS3, PS3_Moderate, BS3, BS3_Moderate, BS3_Supporting corresponded to the MAVE evidence codes. BA1, BS1, BS1_Supporting correspond to the allele frequency codes. Also refer to Table S18.

Conflicts of Interest**:** JRL has stock ownership in 23andMe, is a paid consultant for Regeneron Genetics Center, and is a coinventor on multiple U.S. and European patents related to molecular diagnostics for inherited neuropathies, eye diseases, and bacterial genomic fingerprinting. JRL serves on the Scientific Advisory Board of Baylor Genetics. EV, JRL, and RAG declare that Baylor Genetics is a Baylor College of Medicine affiliate that derives revenue from genetic testing. BCM and Miraca Holdings have formed a joint venture with shared ownership and governance of Baylor Genetics which performs clinical microarray analysis and other genomic studies (exome sequencing and whole genome sequencing) for patient and family care. EV is a co-founder of Codified Genomics, a provider of genetic interpretation.

Code and Data Availability**:** All the code for replicating all the figures and tables in the main text and supplement is readily accessible at <https://github.com/MoezDawood/ReducingVariantClassificationInequities.git>. Additionally, all input data derived from both gnomAD v2.1.1 and gnomAD v3.1.2 (non v2) is linked at the GitHub above. For the *All of Us* analysis, both the input data and associated code are accessible through the *All of Us* workbench and will be promptly shared with requesters with approved workbench access. The code used for analysis of the *All of Us* data is the same as in the above GitHub with minor modifications made that are specific to the *All of Us* Researcher Workbench. All the input data used from *All of Us* is publicly available at the *All of Us* Public Data Browser (v7; <https://databrowser.researchallofus.org/variants>). Complete rankings across all three databases of all clinically curated genes by VUS/PorLP/BorLB/CI/ND allele prevalence difference are also linked to the GitHub. All variant reclassifications will be submitted to ClinVar.

Keywords**:** MAVE, Multiplexed Assay of Variant Effects, Variants of Uncertain Significance, VUS, Pathogenic, Benign, genetic ancestry, equity, inequity, *All of Us*, gnomAD, missense
